# Sex-specific trajectories of molecular cardiometabolic trait concentrations through childhood, adolescence and young adulthood: a cohort study

**DOI:** 10.1101/2021.07.28.21261181

**Authors:** Linda M. O’Keeffe, Kate Tilling, Joshua A. Bell, Matthew A. Lee, Deborah A. Lawlor, George Davey Smith, Patricia M. Kearney

**Author notes:** **Corresponding author:** Dr Linda M. O’Keeffe, School of Public Health, 4^th^ Floor Western Gateway Building, University College Cork, Ireland **Email:**.

## Abstract

**Background:** Causal risk factors and predictive biomarkers for cardiometabolic diseases are increasingly being identified from comprehensive metabolomic profiling in epidemiological studies. The changes which typically occur in molecular cardiometabolic traits across early life are not well characterised.

**Methods:** We quantified sex-specific trajectories of 148 metabolic trait concentrations including various lipoprotein subclasses from age 7y to 25y. Data were from offspring of the Avon Longitudinal Study of Parents and Children birth cohort study. Outcomes included concentrations of 148 traits quantified using nuclear magnetic resonance spectroscopy measured at 7y, 15y, 18y and 25y. Sex-specific trajectories of each trait concentration were modelled using linear spline multilevel models with robust standard errors.

**Findings:** 7,065-7,626 participants (11,702-14,797 repeated measures) were included in analyses. Females had higher very-low-density lipoprotein (VLDL) particle concentrations at 7y. VLDL particle concentrations decreased from 7y to 25y with larger decreases in females, leading to lower VLDL particle concentrations at 25y in females. For example, females had 0.25 SD (95% Confidence Interval (CI), 0.20, 0.31) higher very small VLDL particle concentration at 7y; mean levels decreased by 0.06 SDs (95% CI, −0.01, 0.13) in males and 0.85 SDs (95% CI, 0.79, 0.90) in females from 7y to 25y leading to 0.42 SD (95% CI, 0.35, 0.48) lower very small VLDL particle concentrations in females at 25y. Females also had higher low-density lipoprotein (LDL) particle concentrations at 7y; these increased from 7y to 25y in both sexes and increases were larger among males. By age 25y, LDL particle concentrations remained higher in females but the sex difference was smaller than in early childhood. Females had lower high-density lipoprotein (HDL) particle concentrations at 7y. HDL particle concentrations increased from 7y to 25y with larger increases among females leading to higher HDL particle concentrations in females at 25y.

**Interpretation:** Childhood and adolescence are important periods for the emergence of sex differences in atherogenic lipids and predictive biomarkers for cardiometabolic disease, mostly to the detriment of males.

**Funding:** Wellcome Trust, Medical Research Council UK, Health Research Board, Ireland

**Research in context:** *Evidence before this study:* Causal risk factors and novel predictive biomarkers for cardiometabolic diseases are increasingly being identified from the recent application of comprehensive metabolomic profiling in epidemiological studies but the change which typically occur in these traits across childhood, adolescence and early adulthood is not well u understood.

*Added value of this study:* In this prospective cohort study with repeat assessments of 148 molecular cardiometabolic traits from comprehensive metabolomic profiling at age 7y, 15y, 18y and 25y, we demonstrate that marked change in levels of causal risk factors and novel predictive biomarkers for cardiometabolic diseases occur from childhood to early adulthood. In addition, our findings suggest that childhood and adolescence are an important life stage for the development of sex differences in atherogenic lipids and predictive biomarkers for cardiometabolic disease, mostly to the detriment of males.

*Implications of all the available evidence:* Findings suggest that cardiometabolic disease prevention targeting childhood and adolescence should be prioritised for both lifelong cardiometabolic disease prevention and sex differences in cardiometabolic risk to the disadvantage of males across the life course.

## Introduction

Cardiometabolic diseases are a leading cause of death globally (1). Cardiometabolic risk factors such as adiposity, blood pressure and circulating lipids, as well as underlying subclinical artery disease likely originate in early life, potentially beginning in childhood and tracking through adolescence into adulthood (2–4). Understanding how risk factors begin, change, and track from childhood to adulthood is important for informing aetiological understanding of cardiometabolic diseases and identifying groups in need of targeted prevention.

To date, longitudinal studies have characterised how conventional cardiometabolic risk factors change over time from childhood to adulthood, including studies of change in adiposity (5–7), blood pressure (5–8), and circulating lipids (triglycerides, high-density lipoprotein (HDL) and non-HDL (5, 6, 9)), as well as glucose and insulin (5, 7, 10). In a prior study of the Avon Longitudinal Study of Parents and Children (ALSPAC), we demonstrated distinct patterns of change in conventional cardiometabolic risk factors through the first decades of life, with change for some risk factors coinciding with the sensitive period of puberty (11, 12). Moreover, we also demonstrated notable sex differences in circulating lipids that began at birth including higher HDL and non-HDL among females which widened further by age 18y (5, 7). While these studies provide important insights into early life course development and change in a small number of cardiometabolic risk factors, few studies to date have characterised change in molecular cardiometabolic traits from metabolomics platforms which are now being used for more granular and high-resolution cardiovascular phenotyping to better understand aetiology (13–18). These platforms include directly measured apolipoprotein-B containing lipoprotein subclasses that cause atherosclerotic plaques and lead to coronary heart disease (CHD), which previous studies could only indirectly estimate using conventional approaches (19). As yet, understanding of how these traits typically begin and change from childhood through to adulthood using population-based/non-clinical samples is lacking, despite potential to provide refined understanding of cardiometabolic disease aetiology. In addition, characterising the specific development of these traits is important due to striking sex differences in cardiometabolic disease risk across the life course which remain poorly understood. The study of sex differences in cardiometabolic health is now widely recognised as an area of unlocked potential for informing improved aetiological understanding and mobilising more effective future prevention opportunities(20).

Using data from ALSPAC, a large contemporary birth cohort study from South west England, we examined sex-specific trajectories of 148 concentrations of molecular cardiometabolic traits, mostly lipoprotein subclasses and fatty acids, but also including glucose and an inflammatory marker, measured using targeted metabolomics on four occasions among the same individuals, from 7y to 25y.

## Methods

### Study population

The ALSPAC is a prospective birth cohort study in South west England. Pregnant women resident in one of the three Bristol-based health districts with an expected delivery date between April 1, 1991 and December 31, 1992 were invited to participate. The study has been described elsewhere in detail (21, 22). ALSPAC initially enrolled a cohort of 14,451 pregnancies, from which 13,867 live births occurred in 13,761 women. Follow-up has included parent and offspring completed questionnaires, links to routine data and clinic attendance. Research clinics were held when the offspring were approximately seven, nine, 10, 11, 13, 15, 18 and 25 years old. Ethical approval for the study was obtained from the ALSPAC Ethics and Law Committee and the Local Research Ethics Committees. The study website contains details of all the data that is available through a fully searchable data dictionary http://www.bristol.ac.uk/alspac/researchers/access/.

### Assessment of cardiometabolic traits

Proton nuclear magnetic resonance (^1^H-NMR) spectroscopy from a targeted metabolomics platform (23, 24) was performed on EDTA-plasma samples from blood samples drawn in clinics at ages 7y, 15y, 18y and 25y to quantify 148 concentrations including cholesterol, triglyceride, and other lipid content in lipoprotein subclass particles, apolipoproteins, fatty acids, glucose, and an inflammatory marker (glycoprotein acetyls). Four traits were not measured at 25y (diacylglycerol, fatty acid chain length, estimated degree of unsaturation and conjugated linoleic acid). Bloods were taken after a minimum of a 6-hour fast (stability in these trait concentrations has been shown over different fasting durations (25)). Laboratory NMR quality control (QC) and further data preparation steps are described in Supplementary Material (eMethods 1).

### Statistical analysis

We used multilevel models to examine sex-specific patterns of change in 148 trait concentrations; 144 traits had measures available from 7y to 25y while four traits had measures available from 7 to 18y (26, 27); thus for these traits change over time is only modelled to age 18y. Multilevel models estimate mean trajectories of the outcome while accounting for the non-independence (i.e. clustering) of repeated measurements within individuals and differences in the number and timing of measurements between individuals (using all available data from all eligible participants under a Missing at Random (MAR) assumption) (28, 29). For inclusion in the present analysis, participants required data on sex and at least one measure of a metabolic trait between 7y and 25y (144 traits) and 7y and 18y (four traits). Given that sex is the exposure in these analyses, we did not adjust for potential confounders because sex cannot be altered by factors that influence the metabolic traits (outcomes). For example, body mass index (BMI) is likely to influence many of the metabolic traits but it could not change participant sex and therefore could not confound the effect of sex on metabolic trait levels (although it might mediate such effects).

#### Model selection

Change in all 148 traits was estimated here using linear spline multilevel models (two levels: measurement occasion and individual). Linear splines allow knot points to be fit at different ages to derive periods in which change is approximately linear. To select the optimal linear spline model for 144 trait concentrations measured from 7y to 25y, we ran a series of models including; model 1: a model with two linear spline periods each (7y to 18y and 18y to 25y) model 2: a second model with two linear spline periods (7y to 15y and 15y to 25y) and model 3: a single slope model (a single age term which assumed constant change from 7y to 25y).

Linear spline periods were chosen to reflect ages in whole years that were closest to mean age at clinics and hence where the density of measures was greatest; note that the same process was carried out to select models for the four traits with measures only available to 18y with a model with two linear spline periods (7y to 15y and 15y to 18y) and single slope (7y to 18y) being compared. For each trait and model, we examined Akaike’s Information Criterion (AIC) as an indicator of model fit with lower AIC values indicative of better model fit. Upon selection of the best fitting model based on AIC, we examined, observed and predicted values of models to further assess model fit. Model selection was carried out in both sexes combined to select an optimal model for each trait that would be comparable between the sexes. However, all trajectories were allowed to vary by sex in our final model for each trait by including an interaction term between the linear spline periods/age and sex. eTable 1 shows a complete list of all 148 outcomes (144 measured up to 25y and four measured to 18y) and the model details for each outcome). Following the above process of model comparisons, 68 of the 144 outcomes measured to 25y had two linear spline periods from 7-15y and 15-25y, 75 of the 144 outcomes had two linear spline periods from 7-18y and 18-25y and one of the 144 outcomes (acetoacetate) was a single slope model from 7y to 25y. The final models selected for the four outcomes measured only up to 18y had two linear spline periods from 7-15y and 15-18y.

Models were estimated with robust standard errors for both fixed effects and individual level random effects to account for skewed distributions in some traits. Unstructured variance-covariance matrices for the individual level random effects were used to estimate most trajectories; a complete list of traits, the optimal linear spline model selected and other model details including details of the variance-covariance matrix for each trait are shown in eTable 1.

For each sex, models directly estimate mean predicted level of each metabolite at 7y (the intercept) and mean predicted slopes in original units (mostly mmol/l), with slopes interpreted as change per year in each metabolite in the respective spline period/age period. Following analysis, these estimates were then used to calculate mean predicted absolute change in each trait level from 7y to 25y using the slopes given by each model. The mean predicted level of each trait at 25y was also estimated. Post-analysis, all of the above estimates were then converted to standard deviation (SD) units by dividing by the sex-combined SD of the observed metabolite at 7y, to aid comparison of results between metabolites.

All trajectories were modelled in MLwiN version 3.04 (30), called from Stata version 15 (31) using the runmlwin command (32). Data visualisation was performed using R (version 3.6.3) using the ggforestplot (0.0.2) package. Further information on our modelling strategy is in eMethods 2.

### Sensitivity analyses

We compared sex differences in metabolic traits at 7y and 25y from the multilevel models with the same differences generated from linear regression analyses at each age separately. This was done to explore the appropriateness of our modelling strategy, compared with more conventional analytic approaches. As outlined, participants included in our analyses required data on sex and at least one measure of each metabolite from 7y to 25y. Mothers of participants included in the analyses tended be of higher household social class and more educated than mothers of participants excluded due to missing data and these differences were similar between females and males (eTable 2). Thus, we performed sensitivity analyses weighted by the probability of being included in our analyses to account for the higher probability of being included due to greater social advantage (see eMethods 3 for further details). We repeated all SD unit analyses standardising with the sex-specific SD of each metabolite at 7y to examine whether our main results (standardised with sex-combined SDs) were similar.

## Results

The characteristics of participants included in analyses by sex (49% male), are shown in Table 1. A total of 148 metabolic trait concentrations were modelled. 7,626 participants (14,797 total repeated measures; 5,444 at 7y, 3048 at 15y, 3,121 at 18y and 3,184 at 25y) were included in analyses of 144 concentrations. 7,065 participants (11,702 total repeated measures; 5,395 at 7y, 3,219 at 15y and 3,088 at 18y) were included in analyses of diacylglycerol, fatty acid chain length, estimated degree of saturation and conjugated linoleic acid as these traits were not available at 25y.

**Table 1.** Characteristics of ALSPAC participants included in the analysis, by sex.

|  | **Females**<br>**n=3,909\*** | **Males**<br>**n=3,717\*** |
| --- | --- | --- |
|  | **n (%)** | **n (%)** |
| **Non-white ethnicity** | 75 (2.2) | 60 (1.8) |
| **Maternal marital status** |  |  |
| Never married | 527 (15.1) | 467 (13.7) |
| Widowed | <5 | 8 (0.2) |
| Divorced | 110 (3.2) | 118 (3.5) |
| Separated | 50 (1.4) | 36 (1.1) |
| 1<sup>st</sup> Marriage | 2581 (73.9) | 2544 (74.8) |
| Marriage 2 or 3 | 224 (6.4) | 229 (6.7) |
| **Household social class †** |  |  |
| Professional | 517 (15.8) | 532 (16.7) |
| Managerial & Technical | 1463 (44.6) | 1404 (44.1) |
| Non-Manual | 793 (24.2) | 792 (24.9) |
| Manual | 343 (10.5) | 328 (10.3) |
| Part Skilled & Unskilled | 161 (4.9) | 128 (4.0) |
| **Maternal education** |  |  |
| Less than O level | 761 (22.2) | 724 (21.6) |
| O level | 1166 (34.1) | 1195 (35.7) |
| A level | 926 (27.1) | 883 (26.4) |
| Degree or above | 569 (16.6) | 548 (16.4) |
| **Mother's Partner's highest educational qualification** |  |  |
| Less than O level | 965 (28.9) | 863 (26.5) |
| O level | 717 (21.5) | 724 (22.2) |
| A level | 916 (27.5) | 918 (28.2) |
| Degree or Above | 737 (22.1) | 750 (23.0) |
| **Maternal smoking during pregnancy** | 660 (18.9) | 657 (19.2) |
|  | **Mean (SD)** | **Mean (SD)** |
| **Gestational age (weeks)** | 3370 (512) | 3469 (578) |
| **Birth weight (g)** | 39.6 (1.8) | 39.3 (1.9) |
| **Maternal age (years)** | 28.8 (4.6) | 29.1 (4.7) |
| **Maternal pre-pregnancy BMI (kg/m<sup>2</sup>)** | 22.8 (3.6) | 22.9 (3.8) |

\*Note that denominators in this table do not sum exactly to N participants included in models due to some missing covariate data which was not required for inclusion in our analyses. † Household social class was measured as the highest of the mother's or her partner's occupational social class using data on job title and details of occupation collected about the mother and her partner from the mother's questionnaire at 32 weeks gestation. Social class was derived using the standard occupational classification (SOC) codes developed by the United Kingdom Office of Population Census and Surveys and classified as I professional, II managerial and technical, IIINM non-manual, IIIM manual, and IV&V part skilled occupations and unskilled occupations.

### VLDL concentrations

Very-low-density lipoprotein (VLDL) particle concentrations were higher in females at 7y, e.g., 0.39 SD (95% Confidence Interval (CI), 0.34, 0.44) higher for very small VLDL (Figure 1 and eTable 3). Except for large and medium concentrations among males which increased over time, most other VLDL particle concentrations decreased from 7y to 25y in both sexes (eFigure 1 and eTable 4) and females had larger decreases compared with males. At 25y, females had lower levels of most VLDL particle concentrations, except for very small VLDL particle concentrations which remained 0.08 SD (95% CI, 0.01, 0.14) higher in females, although the difference had reduced in magnitude. Patterns were broadly similar for lipid content in VLDL particles over time.

**Figure 1.**
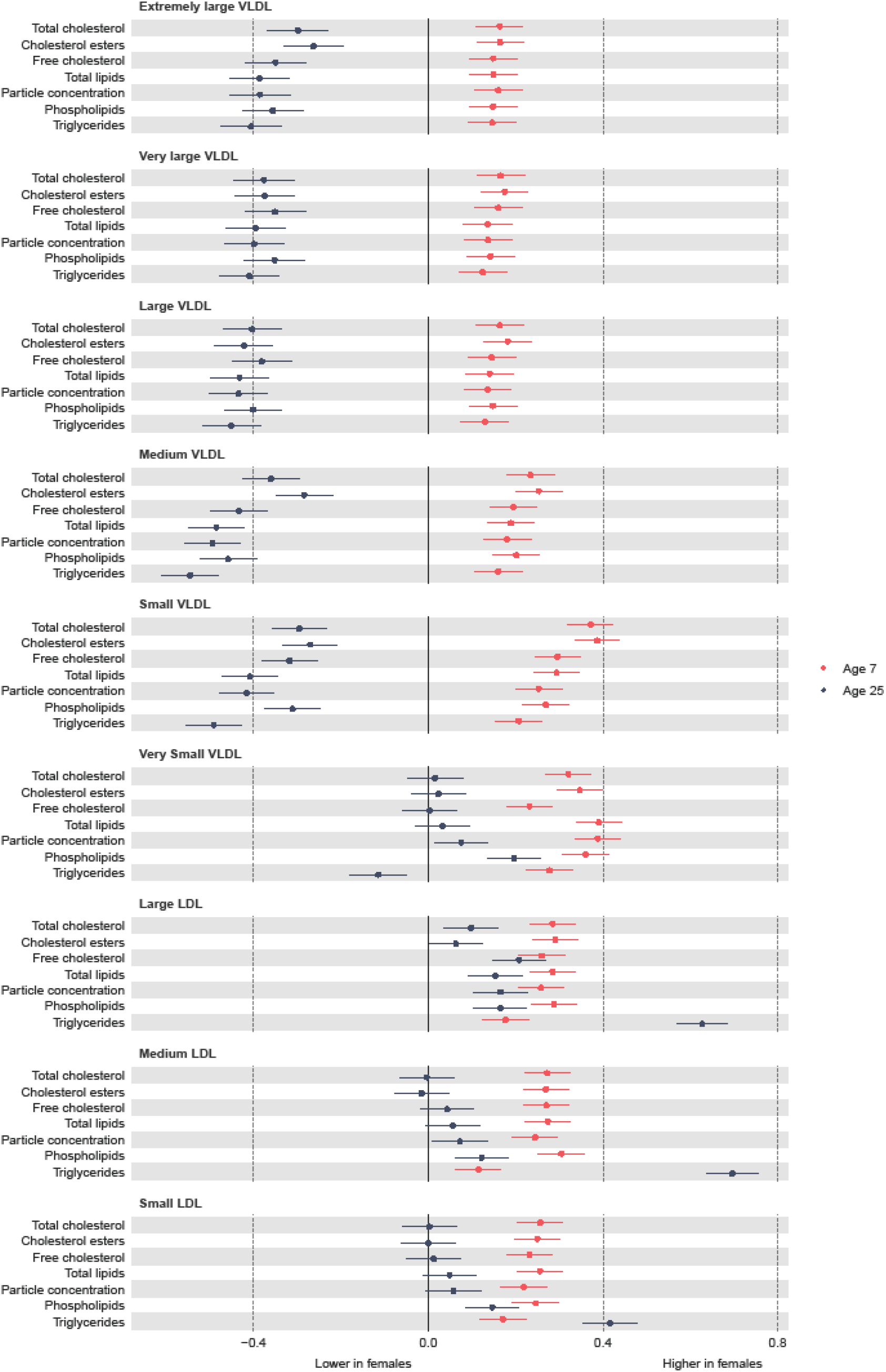
Mean sex difference in VLDL and LDL lipoprotein concentrations in SD units at 7y and 25y, estimated from multilevel models. **Legend:** LDL, low-density lipoprotein; VLDL, very-low-density lipoprotein.

### LDL concentrations

Most low-density lipoprotein (LDL) particle concentrations were higher in females at 7y, e.g., 0.26 SD (95% CI, 0.21, 0.31) higher for large LDL (Figure 1 and eTable 3). LDL particle concentrations increased from 7y to 25y (eFigure 1 and eTable 4). At 25y, higher levels of LDL particle concentrations persisted in females, e.g., 0.17 SD (95% CI, 0.1, 0.23) higher for large LDL but the difference had reduced in magnitude due to smaller increases in females compared with males. These patterns were similar for lipid content in LDL particles, except for triglycerides in LDL; females had higher levels of triglycerides in LDL at 7y, and this difference widened at 25y in contrast to a reduction in the difference for other lipid particles in LDL. This reduction appeared to be driven by smaller decreases and larger increases in triglyceride in LDL particles over time in females.

### HDL concentrations

Most high-density lipoprotein (HDL) particle concentrations were lower in females at age 7y, e.g., −0.1 SD (95% CI, −0.15, −0.05) lower for very large HDL (Figure 2 and eTable 3). Very large HDL particle concentrations decreased from 7y to 25y in both sexes and females had larger decreases than males (eFigure 2 and eTable 4). Large HDL particle concentrations increased from 7y to 25y in females but decreased in males. Most medium and small HDL particle concentrations increased from 7y to 25y in both sexes with larger increases in females. At 25y, females had higher levels of all HDL particle concentrations, e.g., 0.82 SD (95% CI, 0.77, 0.87) higher for very large HDL. Patterns were similar for different lipid contents in HDL particles.

**Figure 2.**
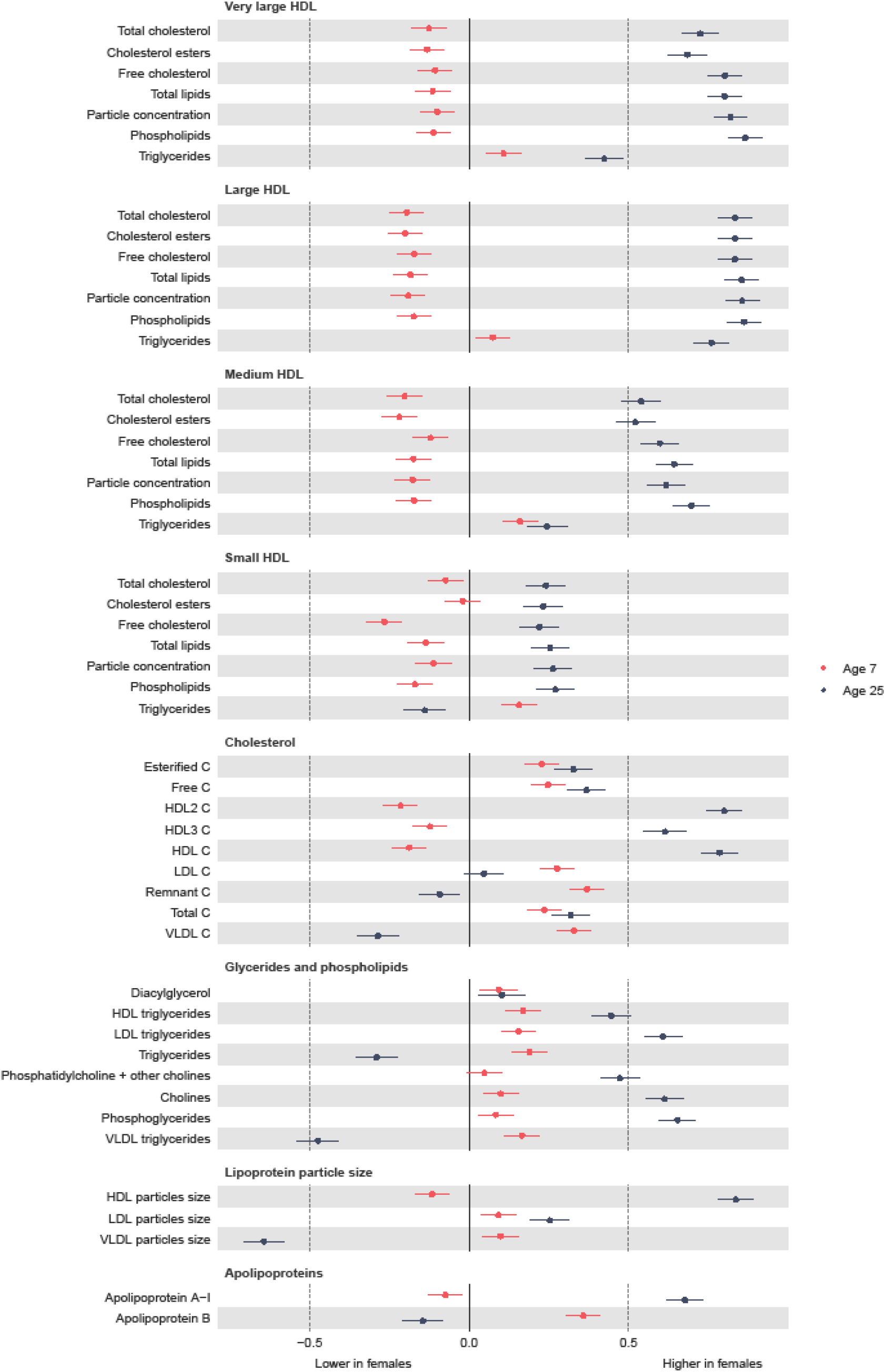
Mean sex difference in lipid in SD units at 7y and 25y, estimated from multilevel models. **Legend:** HDL, high-density lipoprotein; LDL, low-density lipoprotein; VLDL, very-low-density lipoprotein. Note that diacylglycerol is only measured up to 18y.

### Cholesterol

HDL cholesterol concentrations were lower in females at 7y (Figure 2 and eTable 3). All other cholesterol concentrations were higher in females at 7y. Most cholesterol concentrations decreased from 7y to 25y except for HDL and HDL2 in females and LDL in males, which increased over time (eFigure 2 and eTable 4). Females had smaller decreases in esterified, free, HDL3 and total cholesterol concentrations and larger decreases in VLDL and remnant cholesterol concentrations. At 25y, females had higher esterified, free, total and HDL cholesterol concentrations and lower VLDL and remnant cholesterol concentrations though levels of LDL cholesterol concentrations were similar between the sexes.

### Glycerides and phospholipids

All glyceride and phospholipid concentrations were higher in females at 7y (Figure 2 and eTable 3). Concentrations decreased over time in both sexes except for HDL triglyceride concentrations in females which increased over time and VLDL triglycerides which did not change between 7 and 25y in males (eFigure 2 and eTable 4). Females had smaller decreases for most traits, except for total triglyceride concentrations which had a larger decrease. At 25y, females had higher levels of most glyceride and phospholipid concentrations except total and VLDL triglycerides which were lower in females.

### Particle size and polipoproteins

LDL and VLDL particle size were larger in females at 7y but HDL particle size was smaller (Figure 2 and eTable 3). HDL and LDL particle size decreased in both sexes from 7y to 25y and this decrease was smaller in females (eFigure 2 and eTable 4). In contrast, VLDL particle size decreased in females and increased in males between 7y and 25y. At 25y, LDL and HDL particle size were larger in females while VLDL particle size was smaller. Apolipoprotein B was higher in females at 7y and decreased from 7y to 25y in both sexes but females had larger decreases. Apolipoprotein B was lower in females at 25y. Apolipoprotein A-1 was lower in females at 7y and increased over time in females but decreased over time in males. Apolipoprotein A-1 was higher in females at 25y.

### Other non-lipid traits

Glycoprotein acetyls were higher in females at 7y (Figure 3 and eTable 3) and decreased in females and increased in males from 7y to 25y (eFigure 3 and eTable 4), such that concentrations remained higher in females at 25y, with a smaller difference than that observed at age 7y. Citrate and lactate were higher in females at 7y and lower (citrate) in females or similar between the sexes (lactate) at 25y. Glucose was lower in females at 7y and this difference widened at 25y, driven by larger decreases in females from 7y to 25y compared with males. All amino acids were higher in females at 7y or similar between the sexes. Most amino acid concentrations decreased over time, except for alanine and phenylalanine in both sexes and branched chain amino acids in males which increased over time. At 25y, all amino acids were lower in females. Except for docosahexaenoic acid and degree of unsaturation, all fatty acids were higher in females at 7y and this difference was similar at 25y due to similar changes from 7y to 25y in females and males (all decreased except for fatty acid chain length which increased over time). Docosahexaenoic acid was higher in females at 7y and this difference widened over time. Degree of unsaturation was lower in females at 7y and higher in females at 18y (note, one of four traits only measured to 18y). Albumin and creatinine were higher in females at 7y and lower in females 25y due to smaller increases in females from 7y to 25y. Ketone bodies such as acetoacetate were higher in females at 7y; this difference was similar at 25y for beta-hydroxybutyrate but acetoacetate and acetate were lower in females at 25y.

**Figure 3.**
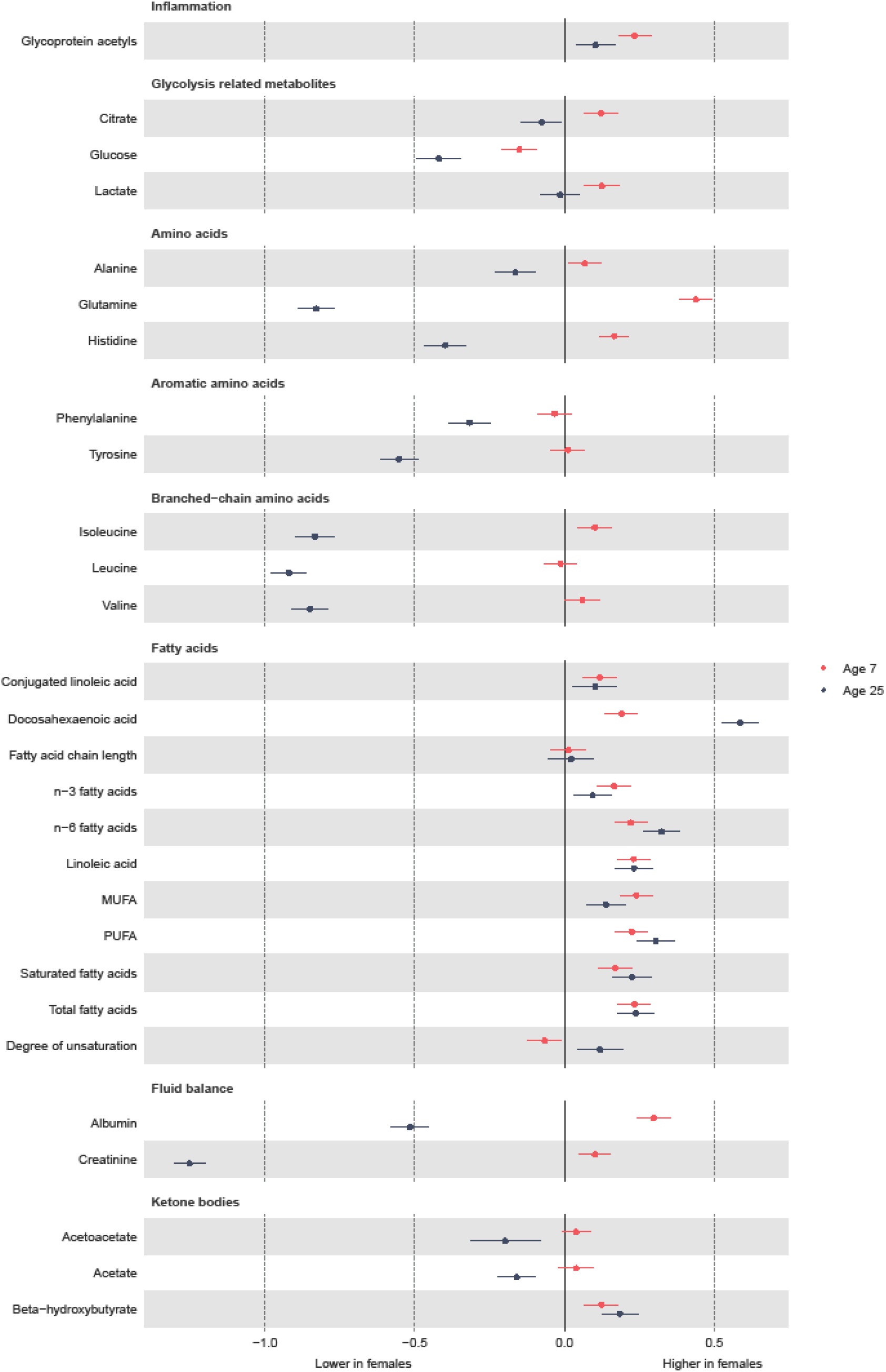
Mean sex difference in other trait concentrations in SD units at 7y and 25y, estimated from multilevel models. **Legend:** MUFA, monounsaturated fatty acids; PUFA, polyunsaturated fatty acids. Note that conjugated linoleic acid, fatty acid chain length and estimated degree of unsaturation are only measured up to 18y.

## Sensitivity and additional analyses

Mean rates of change in original units in each linear spline period are shown in eTable 5. Sex differences in each trait at 7y and 25y estimated from multilevel models were very similar to those obtained from linear regression (eTable 6). Our findings were also very similar in analyses weighted by the probability of being included in analyses (eFigures 4-6). Results were also similar when repeated standardising by the sex-specific mean and SD at 7y (eFigures 7-9).

## Discussion

In this prospective UK birth cohort study, we examined sex-specific trajectories of 148 molecular traits from a targeted metabolomics platform, each measured repeatedly from childhood to early adulthood. Overall, our findings demonstrate substantial changes in molecular cardiometabolic traits from early childhood to early adulthood. However, the magnitude of change for many atherogenic (apolipoprotein B containing) VLDL traits and non-causal HDL traits was more adverse among males, leading to higher levels of VLDL traits and lower levels of non-causal HDL traits at 25y in males. Similarly, despite higher levels of many apolipoprotein B containing LDL traits in early childhood in females, change in these between 7y and 25y was more adverse in males, leading to similar levels of LDL traits between females and males at 25y or only modestly higher levels in females compared with the larger differences observed in early childhood. Our findings suggest that childhood and adolescence are important periods for the emergence of sex differences in atherogenic lipids and predictive biomarkers for cardiometabolic diseases, mostly to the detriment of males.

CHD risk tends to appear higher in males until mid-life, after which risk becomes more similar between females and males (33). Our findings demonstrating marked changes in traits from age 7y to 25y suggest that the early life course may be important for establishing early adult sex differences in levels of VLDL and LDL particles and biomarkers for cardiometabolic disease such as glucose and HDL. In another study in the parents of participants in this cohort using more basic modelling, absolute levels of a number of traits studied here were shown to change little from 25y to 50y (34). In our study, key atherogenic non-HDL lipids such as small VLDL particle concentration were 0.25 SDs lower in males at 7y but 0.40 SDs higher in males at 25y; in the aforementioned study, these traits were approximately 0.5 SDs higher among their fathers compared with their mothers at 50y, suggesting little change in these traits from 25y onwards. Similarly, large LDL particle concentrations were 0.20 SD higher in females in our study at 25y and these were similarly higher in their mothers compared with fathers at 50y. However, in the parent cohort directly measured apolipoprotein B concentration was higher among mothers compared with fathers at 50y suggesting that the higher levels of this observed at 25y among males in our study may not track to 50y, though differences in statin use seen in other UK cohorts may explain some sex differences in these in midlife(17). Although further work is required to study the tracking of early adulthood levels of traits and sex differences in traits into mid-life in our cohort, our findings suggest that childhood and adolescence may be an important period for the emergence of atherogenic lipid levels including smaller VLDL and all LDL subclasses which contribute to the formation of atherosclerotic plaques (19).

The unique sex-specific development of VLDL and LDL lipid profiles across adolescence resulting in a tendency toward higher VLDL lipids among males and higher LDL lipids in females may have implications for more tailored and effective targeting of lipid lowering therapies in females and males. At present, statins are the first line treatment for reduction of apolipoprotein B particles from the blood via upregulation of LDL receptors and removal of LDL from plasma (19). However, the measurement of other apolipoprotein B containing particles now offers opportunities to develop and guide novel lipid lowering therapies which target other particles such as VLDL; such lipid lowering therapies may offer more options for sex-specific targeting of therapies that takes into account the unique sex-specific profile of apolipoprotein B containing particles in females and males in adulthood. However, replication of the sex-specific trajectories found here, including in cohorts with a greater number of repeated measures and longer follow-up are required.

While few studies to date have quantified change in molecular cardiometabolic traits from childhood to early adulthood, our results are comparable with longitudinal analyses of conventional cardiometabolic risk factors. In the Bogalusa Heart Study (N=4,321) which included White and Black participants age 5 to 26 years (35), females had higher LDL cholesterol from 5 to 10 years; however, a ‘male-female’ cross-over arose for LDL cholesterol during adolescence (35). Though LDL cholesterol in that study was measured using the Friedwald equation and thus more prone to measurement error due to inability to exclude IDL particles from measurement, the general pattern of higher levels in females in early childhood and movement toward a cross-over in the sex difference was observed. In addition, males in that study also had higher HDL cholesterol from age five to 10 years; similar to our study, HDL cholesterol decreased over time in both sexes but to a greater degree in males leading to a `male-female’ cross-over in HDL cholesterol at age ∼ 13 or 14, resulting in higher HDL cholesterol in females after this age which persisted into early adulthood. The `male-female’ cross-over from higher levels of HDL cholesterol in males in childhood to higher levels in females from adolescence/early adulthood was also observed in the Minneapolis Cohort Study (6) and Project Heartbeat! (8). Similar to our study, the Minneapolis Cohort Study demonstrated no strong sex differences in LDL cholesterol at 18y while in Project Heartbeat!, a sex difference to the disadvantage of females emerged between childhood and age 18y which contrasts the narrowing of the sex difference that we observed switching from higher levels of LDL cholesterol at 7y in females to similar levels between females and males at 25y. The results in both studies for triglycerides, however, were generally compatible with ours, demonstrating no strong sex differences in early childhood and the emergence of a higher levels of triglycerides in males around age 14 which widened and persisted until the end of follow-up at age 18/19 years.

### Strengths and limitations

There are several strengths to our study including the use of 148 molecular cardiometabolic trait concentrations from a targeted metabolomics platform measured on four occasions from childhood to early adulthood to characterise early life course trajectories. We used multilevel models which take account of clustering of repeated measures within individuals and the correlation between measures over time. Multilevel models also allow inclusion of all participants with at least one measure of a risk factor, thereby minimising selection bias compared with complete case approaches which require all participants to have all measures at each occasion for inclusion in analyses. While repeated NMR measures data are valuable, the number of repeated measures available is relatively sparse for the purposes of trajectory modelling, with, for instance, a period of 8 years between the 7y and 15y. As a result, our modelling approach assumed that cardiometabolic risk factors changed linearly between measurement occasions and we were not able to explore other non-linear patterns of change. While our modelling approach did minimise bias driven by use of complete case approaches for our outcome, bias may still be introduced due to exclusion of participants who did not attend any NMR clinic and do not have at least one measure of the outcome. Participants included in our analyses were more advantaged than those excluded from analyses. However, our sensitivity analyses weighted by the probability of inclusion in analyses did not differ from our main analyses suggesting our results are unlikely to be strongly driven by selection into our analyses (out of the original cohort). Finally, participants were predominantly of White ethnicity and more socially advantaged and thus, our results may not be generalizable to other populations.

## Conclusion

Childhood and adolescence are important periods for the emergence of sex differences in causal atherogenic lipids and predictive biomarkers for cardiometabolic disease, mostly to the detriment of males. Replication in larger independent studies with more repeated measures is required.

## Data Availability

Data are available upon submission and approval of a proposal to the ALSPAC Executive. Further information is available at http://www.bristol.ac.uk/alspac/researchers/our-data/.

## Author contributions

LMOK had the idea for the study, performed all analyses and wrote the manuscript up for publication. ML performed initial data preparation steps for additional quality control and provided critical revisions to the manuscript. All other authors provided critical revisions to the manuscript. LMOK will act as a guarantor for the work.

## Acknowledgements

We are extremely grateful to all the families who took part in this study, the midwives for their help in recruiting them, and the whole ALSPAC team, which includes interviewers, computer and laboratory technicians, clerical workers, research scientists, volunteers, managers, receptionists and nurses.

