## Supplementary Material for "Sex-specific trajectories of molecular cardiometabolic trait concentrations through childhood, adolescence and young adulthood: a cohort study"

**eMethods 1 Details of quality control**

*Laboratory*

For each sample, the NMR spectra were analysed for absolute metabolite quantification (molar concentration) in automated fashion. A ridge regression model was applied for quantification of each metabolite in order to overcome the problems of heavily overlapping spectral data. Quantification of lipoprotein lipid data was performed by calibrating against high performance liquid chromatography methods, and then individually cross-validated against NMR-independent lipid data. Low-molecular-weight metabolites, as well as lipid extract measures, were quantified as mmol/l based on regression modelling calibrated against a set of manually fitted metabolite measures. The calibration data was quantified based on iterative line-shape fitting analysis using PERCH NMR software (PERCH Solutions Ltd., Kuopio, Finland). Absolute quantification could not be directly established for the lipid extract measures due to experimental variation in the lipid extraction protocol. Therefore, serum extract metabolites have been scaled via the total cholesterol as quantified from the native serum LIPO spectrum. We have previously shown strong correlation between the NMR and clinical chemistry measures that are available from both methods.

*Data preparation*

Prior to statistical analysis, preparation of metabolomics data was performed for each occasion separately using the R package metaboprep (https://github.com/MRCIEU/metaboprep) (version 0.0.1)[1]. QC was performed excluding the derived metabolomics measures from missingness and clustering. Briefly, individuals, and then metabolites, with high missingness (>=80%) were removed. Missingness was then re-calculated for individuals and metabolites, with removal based on >=20% missingness.

Individuals were then removed based on total sum abundance, considering outliers as > 5 standard deviations away from the mean. Outliers are identified as being > 5 standard deviations away from the mean of principal component 1 and 2 and were excluded.

| eTable1: Metabolic trait subclass, name, units and multilevel model details for each | | | | | |
|---|---|---|---|---|---|
| **Molecular class** | **Lipid, lipoprotein or metabolite name** | **Units** | **Knot** | **Level 2 variance** | **Level 1 variance** |
| **Extremely large VLDL** | Concentration of chylomicrons and extremely large VLDL particles | mol/l | 15 | Intercept & splines; unstructured | Intercept |
| | Total lipids in chylomicrons and extremely large VLDL | mmol/l | 15 | Intercept & splines; matrix a = (1, 1, 1, 1, 1, 0) | Intercept |
| | Phospholipids in chylomicrons and extremely large VLDL | mmol/l | 15 | Intercept & splines; unstructured | Intercept |
| | Total cholesterol in chylomicrons and extremely large VLDL | mmol/l | 18 | Intercept & splines; matrix a = (1, 1, 1, 1, 1, 0) | Intercept |
| | Cholesterol esters in chylomicrons and extremely large VLDL | mmol/l | 18 | Intercept & splines; matrix a = (1, 1, 1, 1, 1, 0) | Intercept |
| | Free cholesterol in chylomicrons and extremely large VLDL | mmol/l | 15 | Intercept & splines; matrix a = (1, 1, 1, 1, 1, 0) | Intercept |
| | Triglycerides in chylomicrons and extremely large VLDL | mmol/l | 15 | Intercept & splines; matrix a = (1, 1, 1, 1, 1, 0) | Intercept |
| **Very large VLDL** | Concentration of very large VLDL particles | mol/l | 15 | Intercept & splines; unstructured | Intercept |
| | Total lipids in very large VLDL | mmol/l | 15 | Intercept & splines; unstructured | Intercept |
| | Phospholipids in very large VLDL | mmol/l | 15 | Intercept & splines; unstructured | Intercept |
| | Total cholesterol in very large VLDL | mmol/l | 15 | Intercept & splines; matrix a = (1, 1, 1, 1, 1, 0) | Intercept |
| | Cholesterol esters in very large VLDL | mmol/l | 18 | Intercept & splines; unstructured | Intercept |
| | Free cholesterol in very large VLDL | mmol/l | 15 | Intercept & splines; matrix a = (1, 1, 1, 1, 1, 0) | Intercept |
| | Triglycerides in very large VLDL | mmol/l | 15 | Intercept & splines; unstructured | Intercept |
| **Large VLDL** | Concentration of large VLDL particles | mol/l | 15 | Intercept & splines; unstructured | Intercept |
| | Total lipids in large VLDL | mmol/l | 15 | Intercept & splines; unstructured | Intercept |
| | Phospholipids in large VLDL | mmol/l | 15 | Intercept & splines; unstructured | Intercept |
| | Total cholesterol in large VLDL | mmol/l | 15 | Intercept & splines; unstructured | Intercept |
| | Cholesterol esters in large VLDL | mmol/l | 15 | Intercept & splines; unstructured | Intercept |
| | Free cholesterol in large VLDL | mmol/l | 15 | Intercept & splines; unstructured | Intercept |
| | Triglycerides in large VLDL | mmol/l | 15 | Intercept & splines; unstructured | Intercept |
| **Medium VLDL** | Concentration of large VLDL particles | mol/l | 15 | Intercept & splines; unstructured | Intercept |
| | Total lipids in small VLDL | mmol/l | 15 | Intercept & splines; unstructured | Intercept |
| | Phospholipids in small VLDL | mmol/l | 15 | Intercept & splines; unstructured | Intercept |
| | Total cholesterol in small VLDL | mmol/l | 15 | Intercept & splines; unstructured | Intercept |
| | Cholesterol esters in small VLDL | mmol/l | 15 | Intercept & splines; unstructured | Intercept |
| | Free cholesterol in small VLDL | mmol/l | 15 | Intercept & splines; unstructured | Intercept |
| | Triglycerides in small VLDL | mmol/l | 15 | Intercept & splines; unstructured | Intercept |
| **Small VLDL** | Concentration of small VLDL particles | mol/l | 15 | Intercept & splines; unstructured | Intercept |
| | Total lipids in small VLDL | mmol/l | 18 | Intercept & splines; unstructured | Intercept |
| | Phospholipids in small VLDL | mmol/l | 15 | Intercept & splines; matrix a = (1, 1, 0, 1, 1, 1) | Intercept |
| | Total cholesterol in small VLDL | mmol/l | 18 | Intercept & splines; unstructured | Intercept |
| | Cholesterol esters in small VLDL | mmol/l | 18 | Intercept & splines; unstructured | Intercept |
| | Free cholesterol in small VLDL | mmol/l | 18 | Intercept & splines; unstructured | Intercept |
| | Triglycerides in small VLDL | mmol/l | 15 | Intercept & splines; unstructured | Intercept |
| **Very small VLDL** | Concentration of very small VLDL particles | mmol/l | 15 | Intercept & splines; matrix a = (1, 1, 0, 1, 1, 1) | Intercept |

| | | | | | |
|---|---|---|---|---|---|
| | Total lipids in very small VLDL | mmol/l | 18 | Intercept & splines; unstructured | Intercept |
| | Phospholipids in very small VLDL | mmol/l | 15 | Intercept & splines; matrix a = (1, 1, 0, 1, 1, 1) | Intercept |
| | Total cholesterol in very small VLDL | mmol/l | 18 | Intercept & splines; unstructured | Intercept |
| | Cholesterol esters in very small VLDL | mmol/l | 18 | Intercept & splines; unstructured | Intercept |
| | Free cholesterol in very small VLDL | mmol/l | 18 | Intercept & splines; unstructured | Intercept |
| | Triglycerides in very small VLDL | mmol/l | 18 | Intercept & splines; unstructured | Intercept |
| **IDl** | Concentration of IDL particles | mol/l | 18 | Intercept & splines; unstructured | Intercept |
| | Total lipids in IDL | mmol/l | 18 | Intercept & splines; unstructured | Intercept |
| | Phospholipids in IDL | mmol/l | 18 | Intercept & splines; unstructured | Intercept |
| | Total cholesterol in IDL | mmol/l | 18 | Intercept & splines; unstructured | Intercept |
| | Cholesterol esters in IDL | mmol/l | 18 | Intercept & splines; unstructured | Intercept |
| | Free cholesterol in IDL | mmol/l | 18 | Intercept & splines; unstructured | Intercept |
| | Triglycerides in IDL | mmol/l | 18 | Intercept & splines; unstructured | Intercept |
| **Large LDL** | Concentration of large LDL particles | mol/l | 18 | Intercept & splines; unstructured | Intercept |
| | Total lipids in large LDL | mmol/l | 18 | Intercept & splines; unstructured | Intercept |
| | Phospholipids in large LDL | mmol/l | 18 | Intercept & splines; unstructured | Intercept |
| | Total cholesterol in large LDL | mmol/l | 18 | Intercept & splines; unstructured | Intercept |
| | Cholesterol esters in large LDL | mmol/l | 18 | Intercept & splines; unstructured | Intercept |
| | Free cholesterol in large LDL | mmol/l | 18 | Intercept & splines; unstructured | Intercept |
| | Triglycerides in large LDL | mmol/l | 18 | Intercept & splines; unstructured | Intercept |
| **Medium LDL** | Concentration of medium LDL particles | mol/l | 18 | Intercept & splines; unstructured | Intercept |
| | Total lipids in medium LDL | mmol/l | 18 | Intercept & splines; unstructured | Intercept |
| | Phospholipids in medium LDL | mmol/l | 15 | Intercept & splines; matrix a = (1, 1, 0, 1, 1, 1) | Intercept |
| | Total cholesterol in medium LDL | mmol/l | 18 | Intercept & splines; unstructured | Intercept |
| | Cholesterol esters in medium LDL | mmol/l | 18 | Intercept & splines; unstructured | Intercept |
| | Free cholesterol in medium LDL | mmol/l | 18 | Intercept & splines; unstructured | Intercept |
| | Triglycerides in medium LDL | mmol/l | 18 | Intercept & splines; matrix a = (1, 1, 1, 1, 1, 0) | Intercept |
| **Small LDL** | Concentration of small LDL particles | mol/l | 18 | Intercept & splines; unstructured | Intercept |
| | Total lipids in small LDL | mmol/l | 18 | Intercept & splines; unstructured | Intercept |
| | Phospholipids in small LDL | mmol/l | 15 | Intercept & splines; matrix a = (1, 1, 0, 1, 1, 1) | Intercept |
| | Total cholesterol in small LDL | mmol/l | 18 | Intercept & splines; unstructured | Intercept |
| | Cholesterol esters in small LDL | mmol/l | 18 | Intercept & splines; unstructured | Intercept |
| | Free cholesterol in small LDL | mmol/l | 18 | Intercept & splines; unstructured | Intercept |
| | Triglycerides in small LDL | mmol/l | 15 | Intercept & splines; matrix a = (1, 1, 1, 1, 1, 0) | Intercept |
| **Very large HDL** | Concentration of very large HDL particles | mol/l | 15 | Intercept & splines; unstructured | Intercept |
| | Total lipids in very large HDL | mmol/l | 15 | Intercept & splines; unstructured | Intercept |
| | Phospholipids in very large HDL | mmol/l | 15 | Intercept & splines; unstructured | Intercept |
| | Total cholesterol in very large HDL | mmol/l | 18 | Intercept & splines; unstructured | Intercept |

| | | | | | |
|---|---|---|---|---|---|
| | Cholesterol esters in very large HDL | mmol/l | 18 | Intercept & splines; unstructured | Intercept |
| | Free cholesterol in very large HDL | mmol/l | 15 | Intercept & splines; unstructured | Intercept |
| | Triglycerides in very large HDL | mmol/l | 15 | Intercept & splines; unstructured | Intercept |
| **Large HDL** | Concentration of large HDL particles | mol/l | 18 | Intercept & splines; unstructured | Intercept |
| | Total lipids in large HDL | mmol/l | 18 | Intercept & splines; unstructured | Intercept |
| | Phospholipids in large HDL | mmol/l | 18 | Intercept & splines; unstructured | Intercept |
| | Total cholesterol in large HDL | mmol/l | 18 | Intercept & splines; unstructured | Intercept |
| | Cholesterol esters in large HDL | mmol/l | 18 | Intercept & splines; unstructured | Intercept |
| | Free cholesterol in large HDL | mmol/l | 18 | Intercept & splines; unstructured | Intercept |
| | Triglycerides in large HDL | mmol/l | 18 | Intercept & splines; unstructured | Intercept |
| **Medium HDL** | Concentration of medium HDL particles | mol/l | 18 | Intercept & splines; unstructured | Intercept |
| | Total lipids in medium HDL | mmol/l | 15 | Intercept & splines; matrix a = (1, 1, 0, 1, 1, 1) | Intercept |
| | Phospholipids in medium HDL | mmol/l | 15 | Intercept & splines; matrix a = (1, 1, 0, 1, 1, 1) | Intercept |
| | Total cholesterol in medium HDL | mmol/l | 18 | Intercept & splines; unstructured | Intercept |
| | Cholesterol esters in medium HDL | mmol/l | 18 | Intercept & splines; unstructured | Intercept |
| | Free cholesterol in medium HDL | mmol/l | 18 | Intercept & splines; unstructured | Intercept |
| | Triglycerides in medium HDL | mmol/l | 15 | Intercept & splines; matrix a = (1, 1, 0, 1, 1, 1) | Intercept |
| **Small HDL** | Concentration of small HDL particles | mol/l | 15 | Intercept & splines; matrix a = (0, 1, 0, 1, 1, 1) | Intercept |
| | Total lipids in small HDL | mmol/l | 15 | Intercept & splines; matrix a = (1, 1, 0, 1, 1, 0) | Intercept |
| | Phospholipids in small HDL | mmol/l | 15 | Intercept & splines; matrix a = (1, 1, 0, 1, 1, 1) | Intercept |
| | Total cholesterol in small HDL | mmol/l | 15 | Intercept & splines; matrix a = (1, 1, 0, 1, 1, 1) | Intercept |
| | Cholesterol esters in small HDL | mmol/l | 15 | Intercept & splines; matrix a = (1, 1, 0, 1, 1, 1) | Intercept |
| | Free cholesterol in small HDL | mmol/l | 18 | Intercept & splines; unstructured | Intercept |
| | Triglycerides in small HDL | mmol/l | 15 | Intercept & splines; unstructured | Intercept |
| **Lipoprotein particle size** | Mean diameter for VLDL particles | nm | 15 | Intercept & splines; unstructured | Intercept |
| | Mean diameter for LDL particles | nm | 15 | Intercept & splines; matrix a = (1, 1, 0, 1, 1, 1) | Intercept |
| | Mean diameter for HDL particles | nm | 15 | Intercept & splines; unstructured | Intercept |
| **Cholesterol concentrations** | Total cholesterol | mmol/l | 18 | Intercept & splines; unstructured | Intercept |
| | Total cholesterol in VLDL | mmol/l | 18 | Intercept & splines; unstructured | Intercept |
| | Remnant cholesterol (non-HDL and non-LDL cholesterol) | mmol/l | 18 | Intercept & splines; unstructured | Intercept |
| | Total cholesterol in LDL | mmol/l | 18 | Intercept & splines; unstructured | Intercept |
| | Total cholesterol in HDL | mmol/l | 18 | Intercept & splines; unstructured | Intercept |
| | Total cholesterol in HDL2 | mmol/l | 18 | Intercept & splines; unstructured | Intercept |
| | Total cholesterol in HDL3 | mmol/l | 18 | Intercept & splines; unstructured | Intercept |
| | Esterified cholesterol | mmol/l | 15 | Intercept & splines; matrix a = (1, 1, 0, 1, 1, 1) | Intercept |
| | Free cholesterol | mmol/l | 18 | Intercept & splines; unstructured | Intercept |
| | Total triglycerides | mmol/l | 15 | Intercept & splines; unstructured | Intercept |

| | | | | | |
|---|---|---|---|---|---|
| **Glycerides and phospholipid concentrations** | Triglycerides in VLDL | mmol/l | 15 | Intercept & splines; unstructured | Intercept |
| | Triglycerides in LDL | mmol/l | 18 | Intercept & splines; matrix a = (1, 1, 1, 1, 1, 0) | Intercept |
| | Triglycerides in HDL | mmol/l | 15 | Intercept & splines; unstructured | Intercept |
| | Diacylglycerol* | mmol/l | 15 | Intercept & splines; matrix a = (1, 1, 1, 1, 1, 0) | Intercept |
| | Total phosphoglycerides (mmol/l) | mmol/l | 18 | Intercept & splines; matrix a = (1, 1, 0, 1, 1, 1) | Intercept |
| | Phosphatidylcholine and other cholines (mmol/l) | mmol/l | 18 | Intercept & splines; unstructured | Intercept |
| | Total cholines (mmol/l) | mmol/l | 15 | Intercept & splines; unstructured | Intercept |
| **Apolipoprotein concentrations** | Apolipoprotein A-1 | g/l | 18 | Intercept & splines; unstructured | Intercept |
| | Apolipoprotein B | g/l | 15 | Intercept & splines; unstructured | Intercept |
| **Fatty acid concentrations** | Total fatty acids | mmol/l | 15 | Intercept & splines; unstructured | Intercept |
| | Fatty acid length* | | 15 | Intercept & splines; unstructured | Intercept |
| | Estimated degree of saturation* | | 15 | Intercept & splines; unstructured | Intercept |
| | 22:6, docosahexaenoic acid | mmol/l | 15 | Intercept & splines; matrix a = (1, 1, 0, 1, 1, 1) | Intercept |
| | 18:2 linoleic acid | mmol/l | 15 | Intercept & splines; unstructured | Intercept |
| | Conjugated linoleic acid* | mmol/l | 15 | Intercept & splines; unstructured | Intercept |
| | Omega-3 fatty acids | mmol/l | 15 | Intercept & splines; matrix a = (1, 1, 0, 1, 1, 1) | Intercept |
| | Omega-6 fatty acids | mmol/l | 15 | Intercept & splines; unstructured | Intercept |
| | Polyunsaturated fatty acids | mmol/l | 15 | Intercept & splines; unstructured | Intercept |
| | Monounsaturated fatty acids; 16:1, 18:1 | mmol/l | 18 | Intercept & splines; unstructured | Intercept |
| | Saturated fatty acids | mmol/l | 15 | Intercept & splines; unstructured | Intercept |
| **Glycolysis related metabolite** | Glucose | mmol/l | 15 | Intercept & splines; matrix a = (1, 1, 1, 1, 1, 0) | Intercept |
| | Lactate | mmol/l | 15 | Intercept & splines; unstructured | Intercept |
| | Citrate | mmol/l | 18 | Intercept & splines; unstructured | Intercept |
| **Amino acid concentrations** | Alanine | mmol/l | 18 | Intercept & splines; unstructured | Intercept |
| | Glutamine | mmol/l | 18 | Intercept & splines; unstructured | Intercept |
| | Histidine | mmol/l | 18 | Intercept & splines; matrix a = (1, 1, 1, 1, 1, 0) | Intercept |
| branched | Isoleucine | mmol/l | 18 | Intercept & splines; matrix a = (1, 1, 1, 1, 1, 0) | Intercept |
| branched | Leucine | mmol/l | 18 | Intercept & splines; unstructured | Intercept |
| branched | Valine | mmol/l | 15 | Intercept & splines; unstructured | Intercept |
| aromatic | Phenylalanine | mmol/l | 18 | Intercept & splines; unstructured | Intercept |
| aromatic | Tyrosine | mmol/l | 15 | Intercept & splines; matrix a = (1, 1, 1, 1, 1, 0) | Intercept |
| **Ketone body concentrations** | Acetate | mmol/l | 15 | Intercept & splines; matrix a = (1, 1, 0, 1, 1, 1) | Intercept |
| | Acetoacetate | mmol/l | Linear | Intercept & slope; unstructured | Intercept |
| | 3-hydroxybutyrate | mmol/l | 18 | Intercept & splines; matrix a = (0, 1, 1, 1, 1, 1) | Intercept |
| **Fluid balance marker** | Creatinine | mmol/l | 18 | Intercept & splines; unstructured | Intercept |
| | Albumin | mmol/l | 18 | Intercept & splines; matrix a = (1, 1, 0, 1, 1, 1) | Intercept |
| **Inflammation marker** | Glycoprotein acetyls, mainly a1-acid glycoprotein | mmol/l | 18 | Intercept & splines; unstructured | Intercept |

*These metabolites were not measured at 25y; all models include data only up to aged 18y. HDL: high-density lipoprotein; IDL: intermediate-density lipoprotein; LDL: low-density lipoprotein; VLDL: very-low-density lipoprotein.

**eMethods 2**

Age (in years) was centred at the first available measure (7y). All models included individual level random effects for the intercept and each linear spline period selected. For 35 of 148 outcomes modelled (eTable 1) the covariance of the individual level random effects (level 2) were set to zero for some parameters to improve model convergence. Models allowing occasion level measurement error to vary with age (level 1 random effects for the slopes) were also explored for each risk factor. However, due to difficulties with convergence given sparsity of measures, our models included only a random effect for the intercept at level 1. All models included an interaction term for sex to allow trajectories to vary by sex. Thus the model for each outcome took the form of $metabolite_{ij} = \beta_0 + \beta_1 + u_{0j} + (\beta_2 + u_{1j})s_{ij1} + (\beta_3 + u_{2j})s_{ij2} + (\beta_4 + u_{1j})s_{ij1} + (\beta_5 + u_{2j})s_{ij2} + e_{ij}$ where for person j at measurement occasion i; $\beta_0$ represents the fixed effect coefficient for the average intercept in males, $\beta_1$ represents the difference between the intercept for females compared with males, $\beta_2$ and $\beta_3$ represent fixed effect coefficients for the average linear slopes of each linear spline in males, $\beta_4$ and $\beta_5$ represent the difference in the fixed effect coefficients for the average linear slopes of each linear spline in females compared with males, $u_{0j}$ to $u_{2j}$ indicate person-specific (or individual level/level 2) random effects for the intercept and slopes respectively, and $e_{ij}$ represents the occasion-specific residuals or measurement error which was allowed to vary by the intercept.

**eMethods 3**

**Characteristics of included vs. included participants**

We examined characteristics associated with not being included in our analyses due to missing data on sex or molecular traits. To do this, we compared the socio-demographic characteristics at birth of mothers and partners of participants included in the analyses compared to those excluded from the analyses. All characteristics were measured during pregnancy or at birth through questionnaires or from routine health records.

Marital status was obtained from antenatal questionnaires and classified as never married, widowed, divorced, separated, first marriage, marriage 2 or 3. Household social class was measured as the highest of the mother's or her partner's occupational social class using data on job title and details of occupation collected about the mother and her partner from the mother's questionnaire at 32 weeks gestation. Social class was derived using the standard occupational classification (SOC) codes developed by the United Kingdom Office of Population Census and Surveys and classified as I professional, II managerial and technical, IIINM non-manual, IIIM manual, and IV&V part skilled occupations and unskilled occupations. A questionnaire at 32 weeks gestation asked mothers to report their educational attainment, which was categorized as below O-Level (Ordinary Level; exams taken in different subjects usually at age 15-16 at the completion of legally required school attendance, equivalent to today's UK General Certificate of Secondary Education), O-Level only, A-Level (Advanced-Level; exams taken in different subjects usually at age 18), or university degree or above. A questionnaire at 32 weeks gestation asked partners to report their educational attainment, which was categorized as below O-Level (Ordinary Level; exams taken in different subjects usually at age 15-16 at the completion of legally required school attendance, equivalent to today's UK General Certificate of Secondary Education), O-Level only, A-Level (Advanced-Level; exams taken in different subjects usually at age 18), or university degree or above. Smoking in the first trimester of pregnancy was self-reported by mothers at 18 weeks gestation. Birthweight and gestational age were derived from clinical records. Maternal age was reported in the mother's antenatal questionnaires. Maternal pre-pregnancy weight and height were self-reported in antenatal questionnaires.

**Weighted sensitivity analysis**

We performed weighted sensitivity analyses using inverse probability weighting to address potential selection bias. The participant level weights were estimated using logistic regression using all socio-demographic characteristics listed above with the addition of sex and were subsequently incorporated into the multilevel models as level two weights which adjust for the unequal probability of selection of the participants.

| eTable 2: Characteristics of offspring included in analyses compared to those excluded due to missing sex, cardiometabolic trait data or attrition from the cohort | | | | | | |
|---|---|---|---|---|---|---|
| | **Female participants included n=3,909*** | **Female participants excluded** | **N excluded females** | **Male participants included n=3,717*** | **Male participants excluded** | **N excluded males** |
| | **n (%)** | **n (%)** | **n** | **n (%)** | **n (%)** | **n** |
| **Non-white ethnicity** | 75 (2.2) | 77 (2.99) | 2573 | 60 (1.8) | 112 (3.7) | 3050 |
| **Maternal marital status** | | | 2887 | | | 3433 |
| Never married | 527 (15.1) | 680 (23.6) | | 467 (13.7) | 853 (24.9) | |
| Widowed | <5 | 6 (0.2) | | 8 (0.2) | <5 | |
| Divorced | 110 (3.2) | 143 (5.0) | | 118 (3.5) | 188 (5.5) | |
| Separated | 50 (1.4) | 58 (2.0) | | 36 (1.1) | 70 (2.0) | |
| 1$^{st}$ Marriage | 2581 (73.9) | 1832 (63.5) | | 2544 (74.8) | 2086 (60.8) | |
| Marriage 2 or 3 | 224 (6.4) | 168 (5.8) | | 229 (6.7) | 234 (6.8) | |
| **Household social class †** | | | 2337 | | | 2758 |
| Professional | 517 (15.8) | 213 (9.1) | | 532 (16.7) | 275 (10.0) | |
| Managerial & Technical | 1463 (44.6) | 893 (38.2) | | 1404 (44.1) | 1062 (38.5) | |
| Non-Manual | 793 (24.2) | 637 (27.3) | | 792 (24.9) | 723 (26.2) | |
| Manual | 343 (10.5) | 411 (17.6) | | 328 (10.3) | 479 (17.4) | |
| Part Skilled & Unskilled | 161 (4.9) | 183 (7.8) | | 128 (4.0) | 219 (7.9) | |
| **Maternal education** | | | 2605 | | | 3088 |
| Less than O level | 761 (22.2) | 1025 (39.4) | | 724 (21.6) | 1238 (40.1) | |
| O level | 1166 (34.1) | 920 (35.3) | | 1195 (35.7) | 1036 (33.6) | |
| A level | 926 (27.1) | 435 (16.7) | | 883 (26.4) | 550 (17.8) | |
| Degree or above | 569 (16.6) | 225 (8.6) | | 548 (16.4) | 264 (8.6) | |
| **Mother's Partner's highest educational qualification** | | | 2479 | | | 2915 |
| Less than O level | 965 (28.9) | 1056 (42.6) | | 863 (26.5) | 1258 (43.2) | |
| O level | 717 (21.5) | 501 (20.2) | | 724 (22.2) | 609 (20.9) | |
| A level | 916 (27.5) | 625 (25.2) | | 918 (28.2) | 657 (22.5) | |
| Degree or Above | 737 (22.1) | 297 (12.0) | | 750 (23.0) | 391 (13.4) | |
| **Maternal smoking during pregnancy** | 660 (18.9) | 891 (30.5) | 2992 | 657 (19.2) | 1126 (32.8) | 3438 |
| | ***Mean (SD)*** | ***Mean (SD)*** | | ***Mean (SD)*** | ***Mean (SD)*** | |
| **Birthweight (g)** | 3370 (512) | 3283 (579) | 3158 | 3469 (578) | 3395 (631) | 3723 |
| **Gestational age (weeks)** | 39.6 (1.8) | 39.2 (2.9) | 3230 | 39.3 (1.9) | 39.0 (3.0) | 3812 |
| **Maternal age (years)** | 28.8 (4.6) | 26.8 (5.0) | 3198 | 29.1 (4.7) | 27.1 (5.1) | 3778 |
| **Maternal pre-pregnancy BMI (kg/m$^2$)** | 22.8 (3.6) | 23.1 (4.1) | 2448 | 22.9 (3.8) | 23.0 (3.9) | 2845 |

*Represents participants included in models of 144 concentrations with data at all four time points; exact denominators in this table will vary due to missing data for characteristics which were not required for inclusion in analyses.

**eTable 3: Mean difference in traits at 7y and 25y comparing females with males**

| | Original units | | SD units | |
|---|---|---|---|---|
| | **Difference (95% CI) in trait at 7y comparing females to males** | **Difference (95% CI) in trait at 25y comparing females to males** | **Difference (95% CI) in trait at 7y comparing females to males** | **Difference (95% CI) in trait at 25y comparing females to males** |
| Concentration of chylomicrons and extremely large VLDL particles (mol/l) | 1.68E-11 (1.11E-11,2.26E-11) | -2.63E-11 (-3.10E-11,-2.16E-11) | 0.16(0.11,0.21) | -0.39(-0.45,-0.32) |
| Total lipids in chylomicrons and extremely large VLDL (mmol/l) | 3.51E-03 (2.22E-03,4.79E-03) | -5.65E-03 (-6.65E-03,-4.64E-03) | 0.15(0.09,0.2) | -0.39(-0.46,-0.32) |
| Phospholipids in chylomicrons and extremely large VLDL (mmol/l) | 4.25E-04 (2.68E-04,5.81E-04) | -6.51E-04 (-7.77E-04,-5.25E-04) | 0.15(0.09,0.2) | -0.36(-0.43,-0.29) |
| Total cholesterol in chylomicrons and extremely large VLDL (mmol/l) | 5.91E-04 (3.97E-04,7.84E-04) | -7.50E-04 (-9.22E-04,-5.77E-04) | 0.16(0.11,0.22) | -0.3(-0.37,-0.23) |
| Cholesterol esters in chylomicrons and extremely large VLDL (mmol/l) | 2.97E-04 (2.01E-04,3.94E-04) | -3.59E-04 (-4.52E-04,-2.65E-04) | 0.16(0.11,0.22) | -0.26(-0.33,-0.19) |
| Free cholesterol in chylomicrons and extremely large VLDL (mmol/l) | 2.84E-04 (1.79E-04,3.88E-04) | -4.12E-04 (-4.94E-04,-3.31E-04) | 0.15(0.09,0.2) | -0.35(-0.42,-0.28) |
| Triglycerides in chylomicrons and extremely large VLDL (mmol/l) | 2.51E-03 (1.57E-03,3.45E-03) | -4.20E-03 (-4.90E-03,-3.49E-03) | 0.15(0.09,0.2) | -0.41(-0.48,-0.34) |
| Concentration of very large VLDL particles (mol/l) | 7.96E-11 (4.77E-11,1.11E-10) | -1.66E-10 (-1.94E-10,-1.37E-10) | 0.14(0.08,0.19) | -0.4(-0.47,-0.33) |
| Total lipids in very large VLDL (mmol/l) | 7.62E-03 (4.55E-03,1.07E-02) | -1.59E-02 (-1.86E-02,-1.32E-02) | 0.14(0.08,0.19) | -0.4(-0.46,-0.33) |
| Phospholipids in very large VLDL (mmol/l) | 1.34E-03 (8.22E-04,1.85E-03) | -2.33E-03 (-2.78E-03,-1.88E-03) | 0.14(0.09,0.2) | -0.35(-0.42,-0.28) |
| Total cholesterol in very large VLDL (mmol/l) | 1.79E-03 (1.20E-03,2.38E-03) | -2.82E-03 (-3.33E-03,-2.30E-03) | 0.17(0.11,0.22) | -0.38(-0.45,-0.31) |
| Cholesterol esters in very large VLDL (mmol/l) | 9.61E-04 (6.68E-04,1.25E-03) | -1.51E-03 (-1.79E-03,-1.24E-03) | 0.18(0.12,0.23) | -0.38(-0.44,-0.31) |
| Free cholesterol in very large VLDL (mmol/l) | 8.59E-04 (5.66E-04,1.15E-03) | -1.21E-03 (-1.45E-03,-9.74E-04) | 0.16(0.11,0.21) | -0.35(-0.42,-0.28) |
| Triglycerides in very large VLDL (mmol/l) | 4.49E-03 (2.52E-03,6.46E-03) | -1.07E-02 (-1.25E-02,-8.97E-03) | 0.12(0.07,0.18) | -0.41(-0.48,-0.34) |
| Concentration of large VLDL particles (mol/l) | 3.61E-10 (2.16E-10,5.06E-10) | -1.11E-09 (-1.28E-09,-9.44E-10) | 0.14(0.08,0.19) | -0.43(-0.5,-0.37) |
| Total lipids in large VLDL (mmol/l) | 2.25E-02 (1.38E-02,3.12E-02) | -6.39E-02 (-7.37E-02,-5.41E-02) | 0.14(0.09,0.2) | -0.43(-0.5,-0.37) |
| Phospholipids in large VLDL (mmol/l) | 4.22E-03 (2.66E-03,5.78E-03) | -1.08E-02 (-1.26E-02,-9.04E-03) | 0.15(0.09,0.2) | -0.4(-0.47,-0.34) |
| Total cholesterol in large VLDL (mmol/l) | 5.87E-03 (3.91E-03,7.83E-03) | -1.32E-02 (-1.54E-02,-1.10E-02) | 0.16(0.11,0.22) | -0.4(-0.47,-0.34) |
| Cholesterol esters in large VLDL (mmol/l) | 3.05E-03 (2.14E-03,3.96E-03) | -6.89E-03 (-7.97E-03,-5.82E-03) | 0.18(0.13,0.24) | -0.42(-0.49,-0.36) |
| Free cholesterol in large VLDL (mmol/l) | 2.83E-03 (1.76E-03,3.89E-03) | -6.30E-03 (-7.41E-03,-5.19E-03) | 0.15(0.09,0.2) | -0.38(-0.45,-0.31) |
| Triglycerides in large VLDL (mmol/l) | 1.24E-02 (7.18E-03,1.76E-02) | -3.99E-02 (-4.58E-02,-3.41E-02) | 0.13(0.08,0.18) | -0.45(-0.52,-0.38) |
| Concentration of medium VLDL particles (mol/l) | 1.05E-09 (7.37E-10,1.37E-09) | -3.34E-09 (-3.78E-09,-2.90E-09) | 0.18(0.13,0.23) | -0.49(-0.56,-0.43) |
| Total lipids in medium VLDL (mmol/l) | 3.73E-02 (2.66E-02,4.80E-02) | -1.09E-01 (-1.23E-01,-9.44E-02) | 0.19(0.14,0.24) | -0.49(-0.55,-0.42) |
| Phospholipids in medium VLDL (mmol/l) | 7.60E-03 (5.56E-03,9.65E-03) | -1.97E-02 (-2.25E-02,-1.70E-02) | 0.2(0.15,0.26) | -0.46(-0.52,-0.39) |
| Total cholesterol in medium VLDL (mmol/l) | 1.19E-02 (9.16E-03,1.46E-02) | -2.07E-02 (-2.44E-02,-1.70E-02) | 0.23(0.18,0.29) | -0.36(-0.43,-0.3) |
| Cholesterol esters in medium VLDL (mmol/l) | 7.01E-03 (5.52E-03,8.49E-03) | -8.63E-03 (-1.06E-02,-6.66E-03) | 0.25(0.2,0.31) | -0.28(-0.35,-0.22) |
| Free cholesterol in medium VLDL (mmol/l) | 4.91E-03 (3.54E-03,6.27E-03) | -1.20E-02 (-1.38E-02,-1.02E-02) | 0.2(0.14,0.25) | -0.43(-0.5,-0.37) |
| Triglycerides in medium VLDL (mmol/l) | 1.78E-02 (1.18E-02,2.39E-02) | -6.84E-02 (-7.65E-02,-6.04E-02) | 0.16(0.11,0.21) | -0.55(-0.61,-0.48) |
| Concentration of small VLDL particles (mol/l) | 1.46E-09 (1.15E-09,1.77E-09) | -3.08E-09 (-3.55E-09,-2.62E-09) | 0.25(0.2,0.31) | -0.42(-0.48,-0.35) |
| Total lipids in small VLDL (mmol/l) | 3.40E-02 (2.79E-02,4.01E-02) | -5.84E-02 (-6.74E-02,-4.94E-02) | 0.29(0.24,0.35) | -0.41(-0.47,-0.35) |
| Phospholipids in small VLDL (mmol/l) | 6.19E-03 (4.97E-03,7.42E-03) | -8.95E-03 (-1.08E-02,-7.14E-03) | 0.27(0.22,0.32) | -0.31(-0.37,-0.25) |
| Total cholesterol in small VLDL (mmol/l) | 1.58E-02 (1.36E-02,1.80E-02) | -1.56E-02 (-1.88E-02,-1.23E-02) | 0.37(0.32,0.42) | -0.3(-0.36,-0.23) |
| Cholesterol esters in small VLDL (mmol/l) | 1.12E-02 (9.71E-03,1.27E-02) | -9.38E-03 (-1.15E-02,-7.23E-03) | 0.39(0.34,0.44) | -0.27(-0.33,-0.21) |
| Free cholesterol in small VLDL (mmol/l) | 4.61E-03 (3.80E-03,5.43E-03) | -6.15E-03 (-7.37E-03,-4.93E-03) | 0.3(0.24,0.35) | -0.32(-0.38,-0.25) |

**eTable 3: Mean difference in traits at 7y and 25y comparing females with males**

| | Original units | | SD units | |
|---|---|---|---|---|
| | **Difference (95% CI) in trait at 7y comparing females to males** | **Difference (95% CI) in trait at 25y comparing females to males** | **Difference (95% CI) in trait at 7y comparing females to males** | **Difference (95% CI) in trait at 25y comparing females to males** |
| Triglycerides in small VLDL (mmol/l) | 1.21E-02 (8.97E-03,1.53E-02) | -3.32E-02 (-3.75E-02,-2.89E-02) | 0.21(0.15,0.26) | -0.49(-0.56,-0.43) |
| Concentration of very small VLDL particles (mol/l) | 1.79E-09 (1.55E-09,2.03E-09) | 5.40E-10 (1.04E-10,9.77E-10) | 0.39(0.34,0.44) | 0.08(0.01,0.14) |
| Total lipids in very small VLDL (mmol/l) | 2.84E-02 (2.47E-02,3.21E-02) | 2.98E-03 (-2.65E-03,8.60E-03) | 0.39(0.34,0.44) | 0.03(-0.03,0.09) |
| Phospholipids in very small VLDL (mmol/l) | 8.72E-03 (7.46E-03,9.98E-03) | 5.91E-03 (4.08E-03,7.74E-03) | 0.36(0.31,0.41) | 0.2(0.14,0.26) |
| Total cholesterol in very small VLDL (mmol/l) | 1.43E-02 (1.20E-02,1.66E-02) | 7.17E-04 (-2.18E-03,3.61E-03) | 0.32(0.27,0.37) | 0.02(-0.05,0.08) |
| Cholesterol esters in very small VLDL (mmol/l) | 1.06E-02 (9.07E-03,1.22E-02) | 7.42E-04 (-1.26E-03,2.74E-03) | 0.35(0.3,0.4) | 0.02(-0.04,0.09) |
| Free cholesterol in very small VLDL (mmol/l) | 3.65E-03 (2.84E-03,4.47E-03) | 4.21E-05 (-9.04E-04,9.88E-04) | 0.23(0.18,0.28) | 0(-0.06,0.07) |
| Triglycerides in very small VLDL (mmol/l) | 5.41E-03 (4.39E-03,6.43E-03) | -2.49E-03 (-3.89E-03,-1.10E-03) | 0.28(0.22,0.33) | -0.11(-0.18,-0.05) |
| Concentration of IDL particles (mol/l) | 4.19E-09 (3.47E-09,4.90E-09) | 4.78E-09 (3.52E-09,6.03E-09) | 0.3(0.25,0.36) | 0.23(0.17,0.29) |
| Total lipids in IDL (mmol/l) | 5.60E-02 (4.69E-02,6.51E-02) | 4.73E-02 (3.44E-02,6.02E-02) | 0.32(0.27,0.37) | 0.22(0.16,0.28) |
| Phospholipids in IDL (mmol/l) | 1.28E-02 (1.04E-02,1.53E-02) | 1.41E-02 (1.09E-02,1.74E-02) | 0.27(0.22,0.32) | 0.26(0.2,0.32) |
| Total cholesterol in IDL (mmol/l) | 3.79E-02 (3.18E-02,4.41E-02) | 2.44E-02 (1.55E-02,3.32E-02) | 0.32(0.27,0.37) | 0.17(0.11,0.23) |
| Cholesterol esters in IDL (mmol/l) | 2.82E-02 (2.38E-02,3.25E-02) | 1.27E-02 (6.27E-03,1.91E-02) | 0.33(0.28,0.38) | 0.12(0.06,0.18) |
| Free cholesterol in IDL (mmol/l) | 9.70E-03 (7.77E-03,1.16E-02) | 1.17E-02 (9.21E-03,1.42E-02) | 0.26(0.21,0.31) | 0.28(0.22,0.34) |
| Triglycerides in IDL (mmol/l) | 5.19E-03 (4.10E-03,6.28E-03) | 8.87E-03 (7.62E-03,1.01E-02) | 0.25(0.2,0.3) | 0.43(0.37,0.49) |
| Concentration of large LDL particles (mol/l) | 6.66E-09 (5.32E-09,8.00E-09) | 6.20E-09 (3.89E-09,8.50E-09) | 0.26(0.21,0.31) | 0.17(0.1,0.23) |
| Total lipids in large LDL (mmol/l) | 6.39E-02 (5.23E-02,7.55E-02) | 4.14E-02 (2.50E-02,5.78E-02) | 0.28(0.23,0.34) | 0.15(0.09,0.21) |
| Phospholipids in large LDL (mmol/l) | 1.32E-02 (1.08E-02,1.55E-02) | 9.27E-03 (5.85E-03,1.27E-02) | 0.29(0.24,0.34) | 0.16(0.1,0.23) |
| Total cholesterol in large LDL (mmol/l) | 4.66E-02 (3.81E-02,5.50E-02) | 1.95E-02 (7.36E-03,3.17E-02) | 0.28(0.23,0.34) | 0.1(0.04,0.16) |
| Cholesterol esters in large LDL (mmol/l) | 3.58E-02 (2.95E-02,4.22E-02) | 9.62E-03 (2.99E-04,1.89E-02) | 0.29(0.24,0.34) | 0.06(0,0.12) |
| Free cholesterol in large LDL (mmol/l) | 1.07E-02 (8.59E-03,1.29E-02) | 9.91E-03 (7.02E-03,1.28E-02) | 0.26(0.21,0.31) | 0.21(0.15,0.27) |
| Triglycerides in large LDL (mmol/l) | 4.09E-03 (2.88E-03,5.31E-03) | 1.26E-02 (1.14E-02,1.37E-02) | 0.18(0.12,0.23) | 0.63(0.57,0.69) |
| Concentration of medium LDL particles (mol/l) | 5.57E-09 (4.39E-09,6.76E-09) | 2.46E-09 (3.41E-10,4.59E-09) | 0.24(0.19,0.3) | 0.07(0.01,0.13) |
| Total lipids in medium LDL (mmol/l) | 3.81E-02 (3.09E-02,4.54E-02) | 9.72E-03 (-1.04E-03,2.05E-02) | 0.27(0.22,0.33) | 0.06(-0.01,0.12) |
| Phospholipids in medium LDL (mmol/l) | 8.01E-03 (6.64E-03,9.38E-03) | 4.33E-03 (2.19E-03,6.47E-03) | 0.3(0.25,0.36) | 0.12(0.06,0.18) |
| Total cholesterol in medium LDL (mmol/l) | 2.83E-02 (2.29E-02,3.37E-02) | -5.03E-04 (-8.72E-03,7.72E-03) | 0.27(0.22,0.32) | 0(-0.07,0.06) |
| Cholesterol esters in medium LDL (mmol/l) | 2.31E-02 (1.86E-02,2.75E-02) | -1.68E-03 (-8.26E-03,4.91E-03) | 0.27(0.22,0.32) | -0.02(-0.08,0.05) |
| Free cholesterol in medium LDL (mmol/l) | 5.22E-03 (4.22E-03,6.22E-03) | 1.17E-03 (-5.04E-04,2.85E-03) | 0.27(0.22,0.32) | 0.04(-0.02,0.1) |
| Triglycerides in medium LDL (mmol/l) | 1.77E-03 (9.68E-04,2.57E-03) | 7.42E-03 (6.80E-03,8.05E-03) | 0.11(0.06,0.17) | 0.7(0.64,0.75) |
| Concentration of small LDL particles (mol/l) | 5.42E-09 (4.13E-09,6.72E-09) | 2.37E-09 (-1.85E-10,4.93E-09) | 0.22(0.17,0.27) | 0.06(0,0.12) |
| Total lipids in small LDL (mmol/l) | 2.27E-02 (1.81E-02,2.73E-02) | 5.67E-03 (-1.54E-03,1.29E-02) | 0.26(0.2,0.31) | 0.05(-0.01,0.11) |
| Phospholipids in small LDL (mmol/l) | 4.50E-03 (3.54E-03,5.46E-03) | 4.06E-03 (2.39E-03,5.72E-03) | 0.25(0.19,0.3) | 0.15(0.09,0.21) |
| Total cholesterol in small LDL (mmol/l) | 1.67E-02 (1.34E-02,2.01E-02) | 2.15E-04 (-5.06E-03,5.49E-03) | 0.26(0.2,0.31) | 0(-0.06,0.06) |
| Cholesterol esters in small LDL (mmol/l) | 1.40E-02 (1.11E-02,1.68E-02) | -9.53E-06 (-4.15E-03,4.13E-03) | 0.25(0.2,0.3) | 0(-0.06,0.06) |

**eTable 3: Mean difference in traits at 7y and 25y comparing females with males**

| | Original units | | SD units | |
|---|---|---|---|---|
| | **Difference (95% CI) in trait at 7y comparing females to males** | **Difference (95% CI) in trait at 25y comparing females to males** | **Difference (95% CI) in trait at 7y comparing females to males** | **Difference (95% CI) in trait at 25y comparing females to males** |
| Free cholesterol in small LDL (mmol/l) | 2.77E-03 (2.15E-03,3.39E-03) | 2.32E-04 (-9.59E-04,1.42E-03) | 0.23(0.18,0.28) | 0.01(-0.05,0.07) |
| Triglycerides in small LDL (mmol/l) | 1.59E-03 (1.10E-03,2.09E-03) | 2.97E-03 (2.52E-03,3.42E-03) | 0.17(0.12,0.22) | 0.42(0.35,0.48) |
| Concentration of very large HDL particles (mol/l) | -1.16E-08 (-1.77E-08,-5.51E-09) | 1.53E-07 (1.44E-07,1.63E-07) | -0.1(-0.15,-0.05) | 0.82(0.77,0.87) |
| Total lipids in very large HDL (mmol/l) | -1.58E-02 (-2.31E-02,-8.47E-03) | 1.51E-01 (1.41E-01,1.61E-01) | -0.11(-0.17,-0.06) | 0.8(0.75,0.85) |
| Phospholipids in very large HDL (mmol/l) | -8.35E-03 (-1.23E-02,-4.43E-03) | 9.26E-02 (8.71E-02,9.81E-02) | -0.11(-0.16,-0.06) | 0.87(0.81,0.92) |
| Total cholesterol in very large HDL (mmol/l) | -8.26E-03 (-1.17E-02,-4.78E-03) | 5.78E-02 (5.33E-02,6.24E-02) | -0.13(-0.18,-0.07) | 0.72(0.67,0.78) |
| Cholesterol esters in very large HDL (mmol/l) | -6.22E-03 (-8.75E-03,-3.68E-03) | 3.80E-02 (3.47E-02,4.13E-02) | -0.13(-0.18,-0.08) | 0.68(0.62,0.74) |
| Free cholesterol in very large HDL (mmol/l) | -2.03E-03 (-3.03E-03,-1.03E-03) | 1.98E-02 (1.85E-02,2.11E-02) | -0.11(-0.16,-0.05) | 0.8(0.75,0.85) |
| Triglycerides in very large HDL (mmol/l) | 5.59E-04 (2.76E-04,8.41E-04) | 2.46E-03 (2.11E-03,2.81E-03) | 0.11(0.05,0.16) | 0.42(0.36,0.48) |
| Concentration of large HDL particles (mol/l) | -4.38E-08 (-5.58E-08,-3.18E-08) | 4.22E-07 (3.96E-07,4.48E-07) | -0.19(-0.24,-0.14) | 0.86(0.8,0.91) |
| Total lipids in large HDL (mmol/l) | -3.19E-02 (-4.10E-02,-2.28E-02) | 2.69E-01 (2.53E-01,2.86E-01) | -0.18(-0.24,-0.13) | 0.85(0.8,0.91) |
| Phospholipids in large HDL (mmol/l) | -1.33E-02 (-1.73E-02,-9.29E-03) | 1.22E-01 (1.14E-01,1.29E-01) | -0.17(-0.23,-0.12) | 0.86(0.81,0.92) |
| Total cholesterol in large HDL (mmol/l) | -1.88E-02 (-2.39E-02,-1.38E-02) | 1.39E-01 (1.30E-01,1.48E-01) | -0.2(-0.25,-0.14) | 0.83(0.78,0.89) |
| Cholesterol esters in large HDL (mmol/l) | -1.53E-02 (-1.92E-02,-1.13E-02) | 1.06E-01 (9.90E-02,1.12E-01) | -0.2(-0.25,-0.15) | 0.83(0.78,0.89) |
| Free cholesterol in large HDL (mmol/l) | -3.58E-03 (-4.67E-03,-2.50E-03) | 3.31E-02 (3.10E-02,3.52E-02) | -0.17(-0.22,-0.12) | 0.83(0.78,0.89) |
| Triglycerides in large HDL (mmol/l) | 2.59E-04 (7.13E-05,4.47E-04) | 8.79E-03 (8.16E-03,9.42E-03) | 0.07(0.02,0.13) | 0.76(0.71,0.81) |
| Concentration of medium HDL particles (mol/l) | -2.98E-08 (-3.90E-08,-2.06E-08) | 2.75E-07 (2.48E-07,3.01E-07) | -0.18(-0.23,-0.12) | 0.62(0.56,0.68) |
| Total lipids in medium HDL (mmol/l) | -1.49E-02 (-1.96E-02,-1.02E-02) | 1.25E-01 (1.14E-01,1.36E-01) | -0.17(-0.23,-0.12) | 0.64(0.59,0.7) |
| Phospholipids in medium HDL (mmol/l) | -8.09E-03 (-1.07E-02,-5.51E-03) | 6.09E-02 (5.60E-02,6.58E-02) | -0.17(-0.23,-0.12) | 0.7(0.64,0.75) |
| Total cholesterol in medium HDL (mmol/l) | -8.22E-03 (-1.04E-02,-6.02E-03) | 5.73E-02 (5.10E-02,6.36E-02) | -0.2(-0.26,-0.15) | 0.54(0.48,0.6) |
| Cholesterol esters in medium HDL (mmol/l) | -7.33E-03 (-9.14E-03,-5.52E-03) | 4.33E-02 (3.84E-02,4.83E-02) | -0.22(-0.27,-0.16) | 0.52(0.46,0.58) |
| Free cholesterol in medium HDL (mmol/l) | -9.05E-04 (-1.31E-03,-4.99E-04) | 1.39E-02 (1.26E-02,1.53E-02) | -0.12(-0.18,-0.07) | 0.6(0.54,0.66) |
| Triglycerides in medium HDL (mmol/l) | 1.26E-03 (8.34E-04,1.70E-03) | 2.73E-03 (2.03E-03,3.43E-03) | 0.16(0.11,0.21) | 0.24(0.18,0.31) |
| Concentration of small HDL particles (mol/l) | -2.83E-08 (-4.23E-08,-1.43E-08) | 1.87E-07 (1.46E-07,2.29E-07) | -0.11(-0.17,-0.06) | 0.26(0.2,0.32) |
| Total lipids in small HDL (mmol/l) | -9.61E-03 (-1.36E-02,-5.65E-03) | 4.13E-02 (3.16E-02,5.10E-02) | -0.14(-0.19,-0.08) | 0.25(0.19,0.31) |
| Phospholipids in small HDL (mmol/l) | -7.62E-03 (-1.01E-02,-5.15E-03) | 2.02E-02 (1.57E-02,2.46E-02) | -0.17(-0.22,-0.11) | 0.27(0.21,0.33) |
| Total cholesterol in small HDL (mmol/l) | -3.28E-03 (-5.76E-03,-8.05E-04) | 2.45E-02 (1.84E-02,3.05E-02) | -0.07(-0.13,-0.02) | 0.24(0.18,0.3) |
| Cholesterol esters in small HDL (mmol/l) | -8.07E-04 (-3.02E-03,1.41E-03) | 2.08E-02 (1.54E-02,2.61E-02) | -0.02(-0.08,0.04) | 0.23(0.17,0.29) |
| Free cholesterol in small HDL (mmol/l) | -2.55E-03 (-3.07E-03,-2.02E-03) | 3.45E-03 (2.50E-03,4.41E-03) | -0.27(-0.32,-0.21) | 0.22(0.16,0.28) |
| Triglycerides in small HDL (mmol/l) | 1.34E-03 (8.78E-04,1.81E-03) | -1.30E-03 (-1.90E-03,-6.97E-04) | 0.16(0.1,0.21) | -0.14(-0.2,-0.07) |
| Mean diameter for VLDL particles (mm) | 1.28E-01 (5.69E-02,2.00E-01) | -7.44E-01 (-8.16E-01,-6.72E-01) | 0.1(0.04,0.15) | -0.64(-0.71,-0.58) |
| Mean diameter for LDL particles (mm) | 9.55E-03 (3.94E-03,1.52E-02) | 4.54E-02 (3.45E-02,5.63E-02) | 0.09(0.04,0.15) | 0.25(0.19,0.31) |
| Mean diameter for HDL particles (mm) | -1.77E-02 (-2.57E-02,-9.76E-03) | 2.05E-01 (1.92E-01,2.18E-01) | -0.12(-0.17,-0.06) | 0.84(0.78,0.89) |
| Serum total cholesterol (mmol/l) | 1.41E-01 (1.10E-01,1.72E-01) | 2.57E-01 (2.08E-01,3.05E-01) | 0.24(0.18,0.29) | 0.32(0.26,0.38) |

**eTable 3: Mean difference in traits at 7y and 25y comparing females with males**

| | Original units | | SD units | |
|---|---|---|---|---|
| | Difference (95% CI) in trait at 7y comparing females to males | Difference (95% CI) in trait at 25y comparing females to males | Difference (95% CI) in trait at 7y comparing females to males | Difference (95% CI) in trait at 25y comparing females to males |
| Total cholesterol in VLDL (mmol/l) | 5.08E-02 (4.27E-02,5.88E-02) | -5.20E-02 (-6.35E-02,-4.04E-02) | 0.33(0.28,0.38) | -0.29(-0.35,-0.22) |
| Remnant cholesterol (non-HDL, non-LDL -cholesterol) (mmol/l) | 8.86E-02 (7.64E-02,1.01E-01) | -2.78E-02 (-4.65E-02,-9.20E-03) | 0.37(0.32,0.42) | -0.09(-0.15,-0.03) |
| Total cholesterol in LDL (mmol/l) | 9.17E-02 (7.45E-02,1.09E-01) | 1.92E-02 (-6.21E-03,4.47E-02) | 0.28(0.22,0.33) | 0.05(-0.01,0.11) |
| Total cholesterol in HDL (mmol/l) | -3.90E-02 (-5.00E-02,-2.81E-02) | 2.67E-01 (2.48E-01,2.85E-01) | -0.19(-0.24,-0.14) | 0.78(0.73,0.84) |
| Total cholesterol in HDL2 (mmol/l) | -2.97E-02 (-3.70E-02,-2.25E-02) | 2.48E-01 (2.31E-01,2.65E-01) | -0.21(-0.27,-0.16) | 0.8(0.75,0.85) |
| Total cholesterol in HDL3 (mmol/l) | -9.15E-03 (-1.31E-02,-5.19E-03) | 2.17E-02 (1.93E-02,2.41E-02) | -0.12(-0.18,-0.07) | 0.61(0.55,0.68) |
| Esterified cholesterol (mmol/l) | 9.74E-02 (7.49E-02,1.20E-01) | 1.88E-01 (1.54E-01,2.22E-01) | 0.23(0.18,0.28) | 0.33(0.27,0.39) |
| Free cholesterol (mmol/l) | 4.48E-02 (3.54E-02,5.43E-02) | 8.63E-02 (7.24E-02,1.00E-01) | 0.25(0.19,0.3) | 0.37(0.31,0.43) |
| Serum total triglycerides (mmol/l) | 7.20E-02 (5.14E-02,9.26E-02) | -1.11E-01 (-1.36E-01,-8.56E-02) | 0.19(0.14,0.24) | -0.29(-0.36,-0.22) |
| Triglycerides in VLDL (mmol/l) | 5.49E-02 (3.70E-02,7.29E-02) | -1.59E-01 (-1.81E-01,-1.37E-01) | 0.17(0.11,0.22) | -0.47(-0.54,-0.41) |
| Triglycerides in LDL (mmol/l) | 7.31E-03 (4.85E-03,9.78E-03) | 2.27E-02 (2.05E-02,2.49E-02) | 0.16(0.1,0.21) | 0.61(0.55,0.67) |
| Triglycerides in HDL (mmol/l) | 3.49E-03 (2.36E-03,4.62E-03) | 1.26E-02 (1.09E-02,1.44E-02) | 0.17(0.11,0.22) | 0.45(0.38,0.51) |
| Diacylglycerol (mmol/l)* | 1.00E-03 (3.85E-04,1.62E-03) | 9.52E-04 (2.71E-04,1.63E-03) | 0.09(0.04,0.15) | 0.1(0.03,0.18) |
| Total phosphoglycerides (mmol/l) | 2.27E-02 (7.82E-03,3.75E-02) | 2.15E-01 (1.97E-01,2.34E-01) | 0.08(0.03,0.14) | 0.65(0.6,0.71) |
| Phosphatidylcholine and other cholines (mmol/l) | 1.31E-02 (-1.70E-03,2.78E-02) | 1.65E-01 (1.44E-01,1.86E-01) | 0.05(-0.01,0.1) | 0.47(0.41,0.53) |
| Total cholines (mmol/l) | 2.95E-02 (1.34E-02,4.56E-02) | 2.40E-01 (2.17E-01,2.62E-01) | 0.1(0.05,0.15) | 0.61(0.56,0.67) |
| Apolipoprotein A-I (g/l) | -8.28E-03 (-1.42E-02,-2.36E-03) | 1.40E-01 (1.28E-01,1.51E-01) | -0.07(-0.13,-0.02) | 0.68(0.62,0.73) |
| Apolipoprotein B (g/l) | 4.42E-02 (3.77E-02,5.06E-02) | -2.25E-02 (-3.21E-02,-1.29E-02) | 0.36(0.31,0.41) | -0.15(-0.21,-0.08) |
| Total fatty acids (mmol/l) | 3.72E-01 (2.86E-01,4.58E-01) | 4.61E-01 (3.41E-01,5.81E-01) | 0.23(0.18,0.29) | 0.24(0.18,0.3) |
| Fatty acid length* | 4.51E-03 (-1.56E-02,2.46E-02) | 7.86E-03 (-1.94E-02,3.51E-02) | 0.01(-0.04,0.07) | 0.02(-0.05,0.1) |
| Estimated degree of unsaturation* | -3.97E-03 (-7.38E-03,-5.72E-04) | 7.57E-03 (2.75E-03,1.24E-02) | -0.07(-0.12,-0.01) | 0.12(0.04,0.19) |
| 22:6, docosahexaenoic acid (mmol/l) | 5.08E-03 (3.64E-03,6.53E-03) | 1.91E-02 (1.72E-02,2.11E-02) | 0.19(0.14,0.24) | 0.59(0.53,0.65) |
| 18:2, linoleic acid (mmol/l) | 1.02E-01 (7.83E-02,1.26E-01) | 1.25E-01 (9.22E-02,1.58E-01) | 0.23(0.18,0.28) | 0.23(0.17,0.29) |
| Conjugated linoleic acid (mmol/l)* | 2.09E-03 (1.08E-03,3.09E-03) | 1.42E-03 (3.96E-04,2.45E-03) | 0.12(0.06,0.17) | 0.1(0.03,0.18) |
| Omega-3 fatty acids (mmol/l) | 1.13E-02 (7.59E-03,1.50E-02) | 7.34E-03 (2.41E-03,1.23E-02) | 0.17(0.11,0.22) | 0.09(0.03,0.16) |
| Omega-6 fatty acids (mmol/l) | 1.09E-01 (8.28E-02,1.35E-01) | 1.98E-01 (1.61E-01,2.35E-01) | 0.22(0.17,0.27) | 0.32(0.26,0.38) |
| Polyunsaturated fatty acids (mmol/l) | 1.20E-01 (9.18E-02,1.49E-01) | 2.06E-01 (1.65E-01,2.46E-01) | 0.22(0.17,0.28) | 0.3(0.24,0.37) |
| Monounsaturated fatty acids; 16:1, 18:1 (mmol/l) | 1.34E-01 (1.04E-01,1.64E-01) | 8.93E-02 (4.82E-02,1.30E-01) | 0.24(0.19,0.29) | 0.14(0.07,0.2) |
| Saturated fatty acids (mmol/l) | 1.14E-01 (7.69E-02,1.50E-01) | 1.59E-01 (1.15E-01,2.04E-01) | 0.17(0.11,0.22) | 0.22(0.16,0.29) |
| Glucose (mmol/l) | -7.74E-02 (-1.06E-01,-4.84E-02) | -1.92E-01 (-2.25E-01,-1.59E-01) | -0.15(-0.21,-0.09) | -0.42(-0.49,-0.35) |
| Lactate (mmol/l) | 5.99E-02 (3.24E-02,8.73E-02) | -7.08E-03 (-3.62E-02,2.20E-02) | 0.12(0.07,0.18) | -0.02(-0.08,0.05) |
| Citrate (mmol/l) | 3.05E-03 (1.67E-03,4.42E-03) | -1.88E-03 (-3.51E-03,-2.46E-04) | 0.12(0.07,0.18) | -0.08(-0.14,-0.01) |
| Alanine (mmol/l) | 4.33E-03 (7.96E-04,7.87E-03) | -9.16E-03 (-1.29E-02,-5.47E-03) | 0.07(0.01,0.12) | -0.16(-0.23,-0.1) |
| Glutamine (mmol/l) | 2.54E-02 (2.23E-02,2.86E-02) | -6.36E-02 (-6.83E-02,-5.89E-02) | 0.44(0.39,0.49) | -0.83(-0.89,-0.77) |

**eTable 3: Mean difference in traits at 7y and 25y comparing females with males**

| | Original units | | SD units | |
|---|---|---|---|---|
| | Difference (95% CI) in trait at 7y comparing females to males | Difference (95% CI) in trait at 25y comparing females to males | Difference (95% CI) in trait at 7y comparing females to males | Difference (95% CI) in trait at 25y comparing females to males |
| Histidine (mmol/l) | 2.38E-03 (1.71E-03,3.05E-03) | -3.20E-03 (-3.75E-03,-2.66E-03) | 0.17(0.12,0.21) | -0.4(-0.47,-0.33) |
| Isoleucine (mmol/l) | 1.81E-03 (8.22E-04,2.81E-03) | -9.90E-03 (-1.07E-02,-9.15E-03) | 0.1(0.05,0.16) | -0.83(-0.9,-0.77) |
| Leucine (mmol/l) | -2.03E-04 (-1.03E-03,6.23E-04) | -1.22E-02 (-1.30E-02,-1.14E-02) | -0.01(-0.07,0.04) | -0.92(-0.98,-0.86) |
| Valine (mmol/l) | 2.09E-03 (1.20E-04,4.06E-03) | -2.49E-02 (-2.67E-02,-2.32E-02) | 0.06(0,0.12) | -0.85(-0.91,-0.79) |
| Phenylalanine (mmol/l) | -2.86E-04 (-7.56E-04,1.84E-04) | -2.00E-03 (-2.42E-03,-1.58E-03) | -0.03(-0.09,0.02) | -0.32(-0.38,-0.25) |
| Tyrosine (mmol/l) | 1.67E-04 (-7.27E-04,1.06E-03) | -5.41E-03 (-6.02E-03,-4.80E-03) | 0.01(-0.05,0.07) | -0.55(-0.62,-0.49) |
| Acetate (mmol/l) | 9.42E-04 (-4.36E-04,2.32E-03) | -5.65E-03 (-7.92E-03,-3.38E-03) | 0.04(-0.02,0.1) | -0.16(-0.22,-0.1) |
| Acetoacetate (mmol/l) | 1.82E-03 (-3.99E-04,4.05E-03) | -2.05E-03 (-3.25E-03,-8.54E-04) | 0.04(-0.01,0.09) | -0.2(-0.31,-0.08) |
| 3-hydroxybutyrate (mmol/l) | 1.21E-02 (6.54E-03,1.76E-02) | 1.61E-02 (1.09E-02,2.14E-02) | 0.12(0.07,0.18) | 0.18(0.12,0.25) |
| Creatinine (mmol/l) | 6.22E-04 (3.06E-04,9.38E-04) | -1.17E-02 (-1.22E-02,-1.12E-02) | 0.1(0.05,0.15) | -10.25(-10.3,-10.2) |
| Albumin (mmol/l) | 1.08E-03 (8.79E-04,1.27E-03) | -2.89E-03 (-3.24E-03,-2.54E-03) | 0.3(0.24,0.35) | -0.52(-0.58,-0.45) |
| Glycoprotein acetyls, mainly a1-acid glycoprotein (mmol/l) | 3.23E-02 (2.48E-02,3.98E-02) | 1.75E-02 (6.60E-03,2.85E-02) | 0.23(0.18,0.29) | 0.1(0.04,0.17) |

*These metabolites (diacylglycerol, fatty acid chain length, estimated degree of saturation and conjugated linoleic acid) were not measured at 25y; all models include data only up to aged 18y and values in this table for these traits are at 7y and 18y respectively. HDL: high-density lipoprotein; IDL: intermediate-density lipoprotein; LDL: low-density lipoprotein; VLDL: very-low-density lipoprotein.

| eTable 4: Mean absolute difference in traits between 7y and 25y in males and females | | | | |
|---|---|---|---|---|
| | Original units | | SD units | |
| | Mean absolute difference between 7y and 25y (95% CI) | | Mean absolute difference between 7y and 25y (95% CI) | |
| | Males | Females | Males | Females |
| | Beta | Beta | | |
| Concentration of chylomicrons and extremely large VLDL particles (mol/l) | -2.95E-11 (-3.48E-11,-2.41E-11) | -7.26E-11 (-7.71E-11,-6.81E-11) | -0.28(-0.33,-0.23) | -0.69(-0.73,-0.65) |
| Total lipids in chylomicrons and extremely large VLDL (mmol/l) | -1.02E-02 (-1.13E-02,-8.99E-03) | -1.93E-02 (-2.03E-02,-1.83E-02) | -0.43(-0.48,-0.38) | -0.82(-0.86,-0.78) |
| Phospholipids in chylomicrons and extremely large VLDL (mmol/l) | -1.37E-03 (-1.52E-03,-1.23E-03) | -2.45E-03 (-2.57E-03,-2.32E-03) | -0.48(-0.53,-0.43) | -0.85(-0.9,-0.81) |
| Total cholesterol in chylomicrons and extremely large VLDL (mmol/l) | -1.53E-03 (-1.72E-03,-1.34E-03) | -2.87E-03 (-3.02E-03,-2.71E-03) | -0.42(-0.47,-0.37) | -0.79(-0.84,-0.75) |
| Cholesterol esters in chylomicrons and extremely large VLDL (mmol/l) | -6.10E-04 (-7.10E-04,-5.11E-04) | -1.27E-03 (-1.35E-03,-1.19E-03) | -0.34(-0.39,-0.28) | -0.7(-0.74,-0.66) |
| Free cholesterol in chylomicrons and extremely large VLDL (mmol/l) | -9.13E-04 (-1.01E-03,-8.18E-04) | -1.61E-03 (-1.69E-03,-1.53E-03) | -0.48(-0.53,-0.43) | -0.84(-0.89,-0.8) |
| Triglycerides in chylomicrons and extremely large VLDL (mmol/l) | -7.28E-03 (-8.13E-03,-6.44E-03) | -1.40E-02 (-1.47E-02,-1.33E-02) | -0.43(-0.47,-0.38) | -0.82(-0.86,-0.78) |
| Concentration of very large VLDL particles (mol/l) | -1.35E-10 (-1.66E-10,-1.04E-10) | -3.80E-10 (-4.05E-10,-3.54E-10) | -0.23(-0.28,-0.18) | -0.65(-0.7,-0.61) |
| Total lipids in very large VLDL (mmol/l) | -1.53E-02 (-1.82E-02,-1.23E-02) | -3.88E-02 (-4.12E-02,-3.63E-02) | -0.27(-0.33,-0.22) | -0.69(-0.73,-0.65) |
| Phospholipids in very large VLDL (mmol/l) | -2.62E-03 (-3.12E-03,-2.12E-03) | -6.29E-03 (-6.70E-03,-5.88E-03) | -0.28(-0.33,-0.23) | -0.67(-0.71,-0.62) |
| Total cholesterol in very large VLDL (mmol/l) | -4.30E-03 (-4.87E-03,-3.73E-03) | -8.90E-03 (-9.37E-03,-8.44E-03) | -0.4(-0.45,-0.35) | -0.82(-0.87,-0.78) |
| Cholesterol esters in very large VLDL (mmol/l) | -2.05E-03 (-2.35E-03,-1.75E-03) | -4.52E-03 (-4.76E-03,-4.29E-03) | -0.37(-0.43,-0.32) | -0.82(-0.87,-0.78) |
| Free cholesterol in very large VLDL (mmol/l) | -2.30E-03 (-2.57E-03,-2.02E-03) | -4.37E-03 (-4.60E-03,-4.14E-03) | -0.43(-0.48,-0.38) | -0.81(-0.86,-0.77) |
| Triglycerides in very large VLDL (mmol/l) | -8.33E-03 (-1.03E-02,-6.40E-03) | -2.36E-02 (-2.52E-02,-2.20E-02) | -0.23(-0.28,-0.18) | -0.65(-0.7,-0.61) |
| Concentration of large VLDL particles (mol/l) | 1.90E-10 (2.41E-11,3.57E-10) | -1.28E-09 (-1.41E-09,-1.16E-09) | 0.07(0.01,0.13) | -0.48(-0.53,-0.43) |
| Total lipids in large VLDL (mmol/l) | 7.76E-04 (-8.96E-03,1.05E-02) | -8.56E-02 (-9.32E-02,-7.81E-02) | 0(-0.06,0.07) | -0.54(-0.58,-0.49) |
| Phospholipids in large VLDL (mmol/l) | 1.36E-03 (-4.06E-04,3.12E-03) | -1.37E-02 (-1.51E-02,-1.23E-02) | 0.05(-0.01,0.11) | -0.48(-0.53,-0.43) |
| Total cholesterol in large VLDL (mmol/l) | -3.42E-03 (-5.59E-03,-1.25E-03) | -2.25E-02 (-2.42E-02,-2.08E-02) | -0.1(-0.16,-0.03) | -0.63(-0.67,-0.58) |
| Cholesterol esters in large VLDL (mmol/l) | 4.91E-05 (-9.97E-04,1.09E-03) | -9.89E-03 (-1.07E-02,-9.10E-03) | 0(-0.06,0.07) | -0.59(-0.64,-0.54) |
| Free cholesterol in large VLDL (mmol/l) | -3.47E-03 (-4.60E-03,-2.33E-03) | -1.26E-02 (-1.35E-02,-1.17E-02) | -0.18(-0.24,-0.12) | -0.65(-0.69,-0.6) |
| Triglycerides in large VLDL (mmol/l) | 2.82E-03 (-3.00E-03,8.64E-03) | -4.95E-02 (-5.40E-02,-4.50E-02) | 0.03(-0.03,0.09) | -0.52(-0.57,-0.47) |
| Concentration of medium VLDL particles (mol/l) | 2.06E-09 (1.66E-09,2.47E-09) | -2.33E-09 (-2.63E-09,-2.03E-09) | 0.35(0.28,0.42) | -0.4(-0.45,-0.35) |
| Total lipids in medium VLDL (mmol/l) | 4.81E-02 (3.46E-02,6.15E-02) | -9.81E-02 (-1.08E-01,-8.82E-02) | 0.24(0.18,0.31) | -0.5(-0.55,-0.45) |
| Phospholipids in medium VLDL (mmol/l) | 8.05E-03 (5.47E-03,1.06E-02) | -1.93E-02 (-2.12E-02,-1.74E-02) | 0.21(0.14,0.28) | -0.51(-0.56,-0.46) |
| Total cholesterol in medium VLDL (mmol/l) | 7.18E-03 (3.77E-03,1.06E-02) | -2.54E-02 (-2.80E-02,-2.28E-02) | 0.14(0.07,0.21) | -0.5(-0.55,-0.45) |
| Cholesterol esters in medium VLDL (mmol/l) | 6.21E-03 (4.41E-03,8.01E-03) | -9.43E-03 (-1.08E-02,-8.05E-03) | 0.22(0.16,0.29) | -0.34(-0.39,-0.29) |
| Free cholesterol in medium VLDL (mmol/l) | 9.50E-04 (-7.32E-04,2.63E-03) | -1.60E-02 (-1.73E-02,-1.47E-02) | 0.04(-0.03,0.1) | -0.64(-0.69,-0.59) |
| Triglycerides in medium VLDL (mmol/l) | 3.28E-02 (2.53E-02,4.04E-02) | -5.35E-02 (-5.90E-02,-4.79E-02) | 0.29(0.23,0.36) | -0.48(-0.53,-0.43) |
| Concentration of small VLDL particles (mol/l) | -3.56E-10 (-7.61E-10,4.85E-11) | -4.90E-09 (-5.21E-09,-4.59E-09) | -0.06(-0.13,0.01) | -0.85(-0.9,-0.79) |
| Total lipids in small VLDL (mmol/l) | -5.76E-02 (-6.55E-02,-4.98E-02) | -1.50E-01 (-1.56E-01,-1.44E-01) | -0.5(-0.57,-0.43) | -10.3(-10.35,-10.24) |
| Phospholipids in small VLDL (mmol/l) | -1.27E-02 (-1.42E-02,-1.12E-02) | -2.78E-02 (-2.91E-02,-2.66E-02) | -0.55(-0.62,-0.49) | -10.21(-10.27,-10.16) |
| Total cholesterol in small VLDL (mmol/l) | -4.64E-02 (-4.91E-02,-4.37E-02) | -7.78E-02 (-8.00E-02,-7.56E-02) | -10.09(-10.15,-10.03) | -10.83(-10.88,-10.77) |
| Cholesterol esters in small VLDL (mmol/l) | -3.29E-02 (-3.47E-02,-3.11E-02) | -5.35E-02 (-5.50E-02,-5.20E-02) | -10.14(-10.2,-10.08) | -10.85(-10.9,-10.8) |
| Free cholesterol in small VLDL (mmol/l) | -1.35E-02 (-1.46E-02,-1.25E-02) | -2.43E-02 (-2.51E-02,-2.35E-02) | -0.87(-0.94,-0.8) | -10.56(-10.61,-10.51) |
| Triglycerides in small VLDL (mmol/l) | 1.85E-03 (-2.08E-03,5.78E-03) | -4.35E-02 (-4.65E-02,-4.05E-02) | 0.03(-0.04,0.1) | -0.74(-0.79,-0.69) |
| Concentration of very small VLDL particles (mol/l) | -2.00E-09 (-2.33E-09,-1.67E-09) | -3.25E-09 (-3.54E-09,-2.95E-09) | -0.43(-0.51,-0.36) | -0.7(-0.77,-0.64) |
| Total lipids in very small VLDL (mmol/l) | -1.05E-01 (-1.09E-01,-1.00E-01) | -1.30E-01 (-1.34E-01,-1.26E-01) | -10.44(-10.51,-10.38) | -10.79(-10.85,-10.74) |
| Phospholipids in very small VLDL (mmol/l) | -1.20E-02 (-1.34E-02,-1.06E-02) | -1.48E-02 (-1.61E-02,-1.35E-02) | -0.49(-0.55,-0.44) | -0.61(-0.67,-0.56) |
| Total cholesterol in very small VLDL (mmol/l) | -7.94E-02 (-8.18E-02,-7.69E-02) | -9.30E-02 (-9.52E-02,-9.08E-02) | -10.77(-10.83,-10.72) | -20.08(-20.13,-20.03) |
| Cholesterol esters in very small VLDL (mmol/l) | -5.88E-02 (-6.05E-02,-5.71E-02) | -6.87E-02 (-7.02E-02,-6.72E-02) | -10.92(-10.97,-10.86) | -20.24(-20.29,-20.19) |
| Free cholesterol in very small VLDL (mmol/l) | -2.06E-02 (-2.14E-02,-1.97E-02) | -2.42E-02 (-2.50E-02,-2.34E-02) | -10.31(-10.36,-10.25) | -10.54(-10.58,-10.49) |
| Triglycerides in very small VLDL (mmol/l) | -1.44E-02 (-1.56E-02,-1.32E-02) | -2.23E-02 (-2.34E-02,-2.12E-02) | -0.74(-0.8,-0.68) | -10.14(-10.2,-10.09) |
| Concentration of IDL particles (mol/l) | 2.41E-09 (1.46E-09,3.36E-09) | 3.00E-09 (2.13E-09,3.87E-09) | 0.18(0.11,0.24) | 0.22(0.16,0.28) |
| Total lipids in IDL (mmol/l) | -1.22E-01 (-1.32E-01,-1.12E-01) | -1.31E-01 (-1.40E-01,-1.22E-01) | -0.69(-0.75,-0.64) | -0.74(-0.8,-0.69) |
| Phospholipids in IDL (mmol/l) | -2.06E-02 (-2.32E-02,-1.80E-02) | -1.93E-02 (-2.17E-02,-1.68E-02) | -0.43(-0.49,-0.38) | -0.41(-0.46,-0.35) |

| eTable 4: Mean absolute difference in traits between 7y and 25y in males and females | | | | |
|---|---|---|---|---|
| | Original units | | SD units | |
| | Mean absolute difference between 7y and 25y (95% CI) | | Mean absolute difference between 7y and 25y (95% CI) | |
| | Males | Females | Males | Females |
| Total cholesterol in IDL (mmol/l) | -8.18E-02 (-8.87E-02,-7.50E-02) | -9.53E-02 (-1.02E-01,-8.91E-02) | -0.68(-0.74,-0.63) | -0.8(-0.85,-0.74) |
| Cholesterol esters in IDL (mmol/l) | -6.02E-02 (-6.52E-02,-5.52E-02) | -7.57E-02 (-8.02E-02,-7.13E-02) | -0.71(-0.77,-0.65) | -0.89(-0.94,-0.84) |
| Free cholesterol in IDL (mmol/l) | -2.16E-02 (-2.36E-02,-1.96E-02) | -1.96E-02 (-2.15E-02,-1.77E-02) | -0.58(-0.64,-0.53) | -0.53(-0.58,-0.48) |
| Triglycerides in IDL (mmol/l) | -1.99E-02 (-2.09E-02,-1.88E-02) | -1.62E-02 (-1.73E-02,-1.51E-02) | -0.96(-10.01,-0.9) | -0.78(-0.83,-0.73) |
| Concentration of large LDL particles (mol/l) | 2.21E-08 (2.04E-08,2.39E-08) | 2.17E-08 (2.01E-08,2.33E-08) | 0.86(0.79,0.92) | 0.84(0.78,0.9) |
| Total lipids in large LDL (mmol/l) | -2.86E-02 (-4.16E-02,-1.57E-02) | -5.12E-02 (-6.31E-02,-3.93E-02) | -0.13(-0.18,-0.07) | -0.23(-0.28,-0.17) |
| Phospholipids in large LDL (mmol/l) | 4.26E-04 (-2.24E-03,3.09E-03) | -3.47E-03 (-5.94E-03,-1.01E-03) | 0.01(-0.05,0.07) | -0.08(-0.13,-0.02) |
| Total cholesterol in large LDL (mmol/l) | -6.51E-03 (-1.61E-02,3.06E-03) | -3.35E-02 (-4.22E-02,-2.49E-02) | -0.04(-0.1,0.02) | -0.2(-0.26,-0.15) |
| Cholesterol esters in large LDL (mmol/l) | 6.79E-04 (-6.64E-03,8.00E-03) | -2.55E-02 (-3.21E-02,-1.90E-02) | 0.01(-0.05,0.06) | -0.21(-0.26,-0.15) |
| Free cholesterol in large LDL (mmol/l) | -7.20E-03 (-9.50E-03,-4.90E-03) | -8.02E-03 (-1.01E-02,-5.90E-03) | -0.17(-0.23,-0.12) | -0.19(-0.25,-0.14) |
| Triglycerides in large LDL (mmol/l) | -2.24E-02 (-2.34E-02,-2.13E-02) | -1.39E-02 (-1.51E-02,-1.27E-02) | -0.97(-10.01,-0.92) | -0.6(-0.65,-0.55) |
| Concentration of medium LDL particles (mol/l) | 2.30E-08 (2.13E-08,2.46E-08) | 1.99E-08 (1.84E-08,2.14E-08) | 10.01(0.93,10.08) | 0.87(0.81,0.94) |
| Total lipids in medium LDL (mmol/l) | 1.63E-02 (7.76E-03,2.48E-02) | -1.21E-02 (-1.99E-02,-4.39E-03) | 0.12(0.06,0.18) | -0.09(-0.14,-0.03) |
| Phospholipids in medium LDL (mmol/l) | -1.31E-03 (-2.94E-03,3.21E-04) | -4.99E-03 (-6.49E-03,-3.49E-03) | -0.05(-0.11,0.01) | -0.19(-0.25,-0.13) |
| Total cholesterol in medium LDL (mmol/l) | 2.11E-02 (1.46E-02,2.76E-02) | -7.76E-03 (-1.36E-02,-1.92E-03) | 0.2(0.14,0.26) | -0.07(-0.13,-0.02) |
| Cholesterol esters in medium LDL (mmol/l) | 2.64E-02 (2.12E-02,3.17E-02) | 1.68E-03 (-3.04E-03,6.41E-03) | 0.31(0.25,0.37) | 0.02(-0.04,0.07) |
| Free cholesterol in medium LDL (mmol/l) | -5.38E-03 (-6.67E-03,-4.10E-03) | -9.43E-03 (-1.06E-02,-8.27E-03) | -0.28(-0.35,-0.21) | -0.49(-0.55,-0.43) |
| Triglycerides in medium LDL (mmol/l) | -5.39E-03 (-6.04E-03,-4.75E-03) | 2.65E-04 (-4.62E-04,9.91E-04) | -0.35(-0.39,-0.31) | 0.02(-0.03,0.06) |
| Concentration of small LDL particles (mol/l) | 1.60E-08 (1.40E-08,1.79E-08) | 1.29E-08 (1.12E-08,1.47E-08) | 0.64(0.56,0.72) | 0.52(0.45,0.59) |
| Total lipids in small LDL (mmol/l) | -6.26E-03 (-1.19E-02,-6.28E-04) | -2.33E-02 (-2.85E-02,-1.81E-02) | -0.07(-0.13,-0.01) | -0.26(-0.32,-0.2) |
| Phospholipids in small LDL (mmol/l) | -7.73E-03 (-8.97E-03,-6.49E-03) | -8.17E-03 (-9.36E-03,-6.98E-03) | -0.42(-0.49,-0.35) | -0.45(-0.51,-0.38) |
| Total cholesterol in small LDL (mmol/l) | 2.74E-03 (-1.41E-03,6.88E-03) | -1.38E-02 (-1.76E-02,-1.00E-02) | 0.04(-0.02,0.11) | -0.21(-0.27,-0.15) |
| Cholesterol esters in small LDL (mmol/l) | 1.11E-02 (7.80E-03,1.45E-02) | -2.83E-03 (-5.88E-03,2.26E-04) | 0.2(0.14,0.26) | -0.05(-0.11,0) |
| Free cholesterol in small LDL (mmol/l) | -8.42E-03 (-9.32E-03,-7.52E-03) | -1.10E-02 (-1.18E-02,-1.01E-02) | -0.7(-0.78,-0.63) | -0.92(-0.98,-0.85) |
| Triglycerides in small LDL (mmol/l) | -3.06E-03 (-3.49E-03,-2.62E-03) | -1.68E-03 (-2.14E-03,-1.22E-03) | -0.33(-0.37,-0.28) | -0.18(-0.23,-0.13) |
| Concentration of very large HDL particles (mol/l) | -2.24E-07 (-2.30E-07,-2.17E-07) | -5.89E-08 (-6.64E-08,-5.13E-08) | -10.94(-2,-10.88) | -0.51(-0.58,-0.45) |
| Total lipids in very large HDL (mmol/l) | -3.56E-01 (-3.63E-01,-3.49E-01) | -1.89E-01 (-1.97E-01,-1.81E-01) | -20.57(-20.62,-20.52) | -10.37(-10.42,-10.31) |
| Phospholipids in very large HDL (mmol/l) | -1.21E-01 (-1.25E-01,-1.17E-01) | -1.96E-02 (-2.40E-02,-1.53E-02) | -10.61(-10.66,-10.56) | -0.26(-0.32,-0.2) |
| Total cholesterol in very large HDL (mmol/l) | -2.32E-01 (-2.36E-01,-2.29E-01) | -1.66E-01 (-1.70E-01,-1.62E-01) | -30.54(-30.59,-30.49) | -20.53(-20.59,-20.47) |
| Cholesterol esters in very large HDL (mmol/l) | -1.80E-01 (-1.83E-01,-1.78E-01) | -1.36E-01 (-1.39E-01,-1.33E-01) | -30.81(-30.86,-30.75) | -20.88(-20.93,-20.82) |
| Free cholesterol in very large HDL (mmol/l) | -5.09E-02 (-5.19E-02,-4.99E-02) | -2.90E-02 (-3.01E-02,-2.80E-02) | -20.68(-20.74,-20.63) | -10.53(-10.59,-10.47) |
| Triglycerides in very large HDL (mmol/l) | -9.31E-03 (-9.61E-03,-9.01E-03) | -7.41E-03 (-7.70E-03,-7.12E-03) | -10.8(-10.86,-10.75) | -10.44(-10.49,-10.38) |
| Concentration of large HDL particles (mol/l) | -2.15E-07 (-2.33E-07,-1.98E-07) | 2.50E-07 (2.31E-07,2.69E-07) | -0.94(-10.02,-0.86) | 10.09(10.01,10.17) |
| Total lipids in large HDL (mmol/l) | -2.35E-01 (-2.47E-01,-2.23E-01) | 6.63E-02 (5.39E-02,7.88E-02) | -10.35(-10.42,-10.28) | 0.38(0.31,0.45) |
| Phospholipids in large HDL (mmol/l) | -8.91E-02 (-9.43E-02,-8.39E-02) | 4.60E-02 (4.04E-02,5.16E-02) | -10.16(-10.23,-10.09) | 0.6(0.53,0.67) |
| Total cholesterol in large HDL (mmol/l) | -1.42E-01 (-1.48E-01,-1.36E-01) | 1.59E-02 (9.30E-03,2.25E-02) | -10.47(-10.53,-10.4) | 0.16(0.1,0.23) |
| Cholesterol esters in large HDL (mmol/l) | -1.10E-01 (-1.15E-01,-1.05E-01) | 1.08E-02 (5.75E-03,1.59E-02) | -10.45(-10.52,-10.39) | 0.14(0.08,0.21) |
| Free cholesterol in large HDL (mmol/l) | -3.16E-02 (-3.31E-02,-3.02E-02) | 5.09E-03 (3.55E-03,6.63E-03) | -10.52(-10.59,-10.44) | 0.24(0.17,0.32) |
| Triglycerides in large HDL (mmol/l) | -3.93E-03 (-4.32E-03,-3.54E-03) | 4.60E-03 (4.10E-03,5.11E-03) | -10.13(-10.24,-10.02) | 10.32(10.18,10.47) |
| Concentration of medium HDL particles (mol/l) | 3.76E-07 (3.58E-07,3.94E-07) | 6.80E-07 (6.60E-07,7.00E-07) | 20.23(20.13,20.34) | 40.04(30.92,40.16) |
| Total lipids in medium HDL (mmol/l) | 7.71E-04 (-6.79E-03,8.33E-03) | 1.40E-01 (1.32E-01,1.49E-01) | 0.01(-0.08,0.1) | 10.64(10.54,10.74) |
| Phospholipids in medium HDL (mmol/l) | -2.09E-02 (-2.43E-02,-1.74E-02) | 4.81E-02 (4.41E-02,5.22E-02) | -0.45(-0.52,-0.37) | 10.03(0.94,10.12) |
| Total cholesterol in medium HDL (mmol/l) | 1.80E-02 (1.36E-02,2.25E-02) | 8.35E-02 (7.89E-02,8.82E-02) | 0.44(0.34,0.55) | 20.06(10.94,20.17) |
| Cholesterol esters in medium HDL (mmol/l) | 1.41E-02 (1.06E-02,1.76E-02) | 6.48E-02 (6.11E-02,6.85E-02) | 0.42(0.32,0.53) | 10.93(10.82,20.04) |
| Free cholesterol in medium HDL (mmol/l) | 3.93E-03 (2.99E-03,4.87E-03) | 1.88E-02 (1.77E-02,1.98E-02) | 0.53(0.4,0.65) | 20.51(20.37,20.64) |
| Triglycerides in medium HDL (mmol/l) | 9.68E-03 (9.12E-03,1.02E-02) | 1.11E-02 (1.06E-02,1.17E-02) | 10.22(10.15,10.3) | 10.41(10.34,10.48) |

| eTable 4: Mean absolute difference in traits between 7y and 25y in males and females | | | | |
|---|---|---|---|---|
| | Original units | | SD units | |
| | Mean absolute difference between 7y and 25y (95% CI) | | Mean absolute difference between 7y and 25y (95% CI) | |
| | Males | Females | Males | Females |
| Concentration of small HDL particles (mol/l) | 6.77E-07 (6.49E-07,7.04E-07) | 8.92E-07 (8.60E-07,9.24E-07) | 20.68(20.57,20.79) | 30.53(30.4,30.66) |
| Total lipids in small HDL (mmol/l) | 5.14E-02 (4.48E-02,5.80E-02) | 1.02E-01 (9.47E-02,1.10E-01) | 0.73(0.63,0.82) | 10.44(10.34,10.55) |
| Phospholipids in small HDL (mmol/l) | -1.43E-02 (-1.75E-02,-1.11E-02) | 1.35E-02 (9.84E-03,1.71E-02) | -0.32(-0.39,-0.25) | 0.3(0.22,0.38) |
| Total cholesterol in small HDL (mmol/l) | 6.17E-02 (5.74E-02,6.61E-02) | 8.95E-02 (8.48E-02,9.42E-02) | 10.39(10.29,10.49) | 20.01(10.91,20.12) |
| Cholesterol esters in small HDL (mmol/l) | 7.68E-02 (7.29E-02,8.07E-02) | 9.84E-02 (9.43E-02,1.02E-01) | 10.93(10.83,20.02) | 20.47(20.36,20.57) |
| Free cholesterol in small HDL (mmol/l) | -1.69E-02 (-1.76E-02,-1.62E-02) | -1.09E-02 (-1.17E-02,-1.01E-02) | -10.77(-10.84,-10.69) | -10.14(-10.22,-10.06) |
| Triglycerides in small HDL (mmol/l) | 7.00E-04 (1.63E-04,1.24E-03) | -1.94E-03 (-2.42E-03,-1.47E-03) | 0.08(0.02,0.14) | -0.23(-0.28,-0.17) |
| Mean diameter for VLDL particles (mm) | 2.84E-01 (2.12E-01,3.57E-01) | -5.88E-01 (-6.48E-01,-5.27E-01) | 0.22(0.16,0.27) | -0.45(-0.5,-0.41) |
| Mean diameter for LDL particles (mm) | -5.89E-02 (-6.73E-02,-5.05E-02) | -2.31E-02 (-3.13E-02,-1.49E-02) | -0.57(-0.65,-0.48) | -0.22(-0.3,-0.14) |
| Mean diameter for HDL particles (mm) | -2.80E-01 (-2.90E-01,-2.70E-01) | -5.72E-02 (-6.65E-02,-4.79E-02) | -10.84(-10.91,-10.78) | -0.38(-0.44,-0.32) |
| Serum total cholesterol (mmol/l) | -4.74E-01 (-5.11E-01,-4.38E-01) | -3.59E-01 (-3.94E-01,-3.24E-01) | -0.79(-0.85,-0.73) | -0.6(-0.66,-0.54) |
| Total cholesterol in VLDL (mmol/l) | -1.27E-01 (-1.37E-01,-1.17E-01) | -2.30E-01 (-2.38E-01,-2.22E-01) | -0.83(-0.89,-0.76) | -10.49(-10.54,-10.44) |
| Remnant cholesterol (non-HDL, non-LDL -cholesterol) (mmol/l) | -2.09E-01 (-2.24E-01,-1.94E-01) | -3.25E-01 (-3.38E-01,-3.13E-01) | -0.87(-0.93,-0.81) | -10.36(-10.41,-10.31) |
| Total cholesterol in LDL (mmol/l) | 1.73E-02 (-2.74E-03,3.74E-02) | -5.51E-02 (-7.32E-02,-3.71E-02) | 0.05(-0.01,0.11) | -0.17(-0.22,-0.11) |
| Total cholesterol in HDL (mmol/l) | -2.85E-01 (-2.99E-01,-2.71E-01) | 2.09E-02 (6.63E-03,3.51E-02) | -10.37(-10.44,-10.31) | 0.1(0.03,0.17) |
| Total cholesterol in HDL2 (mmol/l) | -1.30E-01 (-1.41E-01,-1.18E-01) | 1.48E-01 (1.36E-01,1.61E-01) | -0.94(-10.02,-0.85) | 10.07(0.99,10.16) |
| Total cholesterol in HDL3 (mmol/l) | -1.54E-01 (-1.57E-01,-1.51E-01) | -1.23E-01 (-1.26E-01,-1.20E-01) | -20.07(-20.11,-20.03) | -10.65(-10.7,-10.61) |
| Esterified cholesterol (mmol/l) | -3.52E-01 (-3.78E-01,-3.27E-01) | -2.62E-01 (-2.87E-01,-2.37E-01) | -0.82(-0.88,-0.76) | -0.61(-0.67,-0.55) |
| Free cholesterol (mmol/l) | -1.39E-01 (-1.50E-01,-1.29E-01) | -9.78E-02 (-1.08E-01,-8.75E-02) | -0.77(-0.83,-0.71) | -0.54(-0.6,-0.48) |
| Serum total triglycerides (mmol/l) | -4.75E-02 (-7.13E-02,-2.37E-02) | -2.30E-01 (-2.49E-01,-2.11E-01) | -0.12(-0.19,-0.06) | -0.61(-0.65,-0.56) |
| Triglycerides in VLDL (mmol/l) | 7.82E-03 (-1.31E-02,2.87E-02) | -2.06E-01 (-2.22E-01,-1.90E-01) | 0.02(-0.04,0.09) | -0.62(-0.67,-0.57) |
| Triglycerides in LDL (mmol/l) | -3.06E-02 (-3.27E-02,-2.85E-02) | -1.52E-02 (-1.75E-02,-1.29E-02) | -0.65(-0.69,-0.6) | -0.32(-0.37,-0.27) |
| Triglycerides in HDL (mmol/l) | -2.98E-03 (-4.35E-03,-1.61E-03) | 6.17E-03 (4.76E-03,7.57E-03) | -0.14(-0.21,-0.08) | 0.3(0.23,0.37) |
| Diacylglycerol (mmol/l)* | -1.97E-03 (-2.57E-03,-1.37E-03) | -2.40E-03 (-3.11E-03,-1.68E-03) | -0.18(-0.24,-0.13) | -0.22(-0.29,-0.16) |
| Total phosphoglycerides (mmol/l) | -3.76E-01 (-3.91E-01,-3.62E-01) | -1.84E-01 (-2.00E-01,-1.67E-01) | -10.39(-10.44,-10.33) | -0.68(-0.74,-0.61) |
| Phosphatidylcholine and other cholines (mmol/l) | -3.62E-01 (-3.79E-01,-3.45E-01) | -2.10E-01 (-2.27E-01,-1.93E-01) | -10.32(-10.38,-10.26) | -0.77(-0.83,-0.7) |
| Total cholines (mmol/l) | -3.85E-01 (-4.02E-01,-3.68E-01) | -1.75E-01 (-1.94E-01,-1.56E-01) | -10.29(-10.35,-10.23) | -0.59(-0.65,-0.52) |
| Apolipoprotein A-I (g/l) | -1.22E-01 (-1.30E-01,-1.13E-01) | 2.63E-02 (1.75E-02,3.52E-02) | -10.09(-10.16,-10.01) | 0.23(0.16,0.31) |
| Apolipoprotein B (g/l) | -1.72E-02 (-2.50E-02,-9.40E-03) | -8.39E-02 (-9.04E-02,-7.73E-02) | -0.14(-0.2,-0.08) | -0.68(-0.73,-0.63) |
| Total fatty acids (mmol/l) | -1.80E+00 (-1.90E+00,-1.70E+00) | -1.71E+00 (-1.80E+00,-1.61E+00) | -10.13(-10.19,-10.07) | -10.07(-10.13,-10.01) |
| Fatty acid length* | 1.57E-01 (1.33E-01,1.81E-01) | 1.05E-01 (7.44E-02,1.35E-01) | 0.44(0.38,0.51) | 0.3(0.21,0.38) |
| Estimated degree of unsaturation* | 1.72E-02 (1.30E-02,2.15E-02) | 2.86E-02 (2.33E-02,3.39E-02) | 0.28(0.21,0.36) | 0.47(0.39,0.56) |
| 22:6, docosahexaenoic acid (mmol/l) | -7.50E-03 (-9.04E-03,-5.95E-03) | 6.54E-03 (4.87E-03,8.21E-03) | -0.28(-0.34,-0.22) | 0.24(0.18,0.31) |
| 18:2, linoleic acid (mmol/l) | -5.42E-01 (-5.68E-01,-5.15E-01) | -5.18E-01 (-5.43E-01,-4.93E-01) | -10.23(-10.29,-10.16) | -10.17(-10.23,-10.12) |
| Conjugated linoleic acid (mmol/l)* | -7.74E-03 (-8.71E-03,-6.78E-03) | -8.89E-03 (-1.03E-02,-7.46E-03) | -0.44(-0.49,-0.38) | -0.5(-0.58,-0.42) |
| Omega-3 fatty acids (mmol/l) | -3.63E-02 (-4.05E-02,-3.22E-02) | -4.03E-02 (-4.43E-02,-3.63E-02) | -0.53(-0.59,-0.47) | -0.59(-0.65,-0.53) |
| Omega-6 fatty acids (mmol/l) | -6.34E-01 (-6.63E-01,-6.04E-01) | -5.44E-01 (-5.73E-01,-5.16E-01) | -10.29(-10.35,-10.23) | -10.11(-10.16,-10.05) |
| Polyunsaturated fatty acids (mmol/l) | -6.70E-01 (-7.02E-01,-6.38E-01) | -5.85E-01 (-6.16E-01,-5.53E-01) | -10.25(-10.31,-10.19) | -10.09(-10.15,-10.03) |
| Monounsaturated fatty acids; 16:1, 18:1 (mmol/l) | -2.16E-01 (-2.51E-01,-1.80E-01) | -2.61E-01 (-2.93E-01,-2.28E-01) | -0.39(-0.45,-0.32) | -0.47(-0.52,-0.41) |
| Saturated fatty acids (mmol/l) | -9.22E-01 (-9.61E-01,-8.82E-01) | -8.76E-01 (-9.13E-01,-8.38E-01) | -10.38(-10.43,-10.32) | -10.31(-10.36,-10.25) |
| Glucose (mmol/l) | -1.68E-01 (-2.02E-01,-1.34E-01) | -2.82E-01 (-3.08E-01,-2.57E-01) | -0.33(-0.39,-0.26) | -0.55(-0.6,-0.5) |
| Lactate (mmol/l) | -4.06E-01 (-4.33E-01,-3.79E-01) | -4.73E-01 (-5.01E-01,-4.44E-01) | -0.84(-0.9,-0.79) | -0.98(-10.04,-0.92) |
| Citrate (mmol/l) | 2.81E-02 (2.66E-02,2.96E-02) | 2.32E-02 (2.18E-02,2.45E-02) | 10.12(10.06,10.18) | 0.92(0.87,0.98) |
| Alanine (mmol/l) | 5.32E-02 (4.95E-02,5.69E-02) | 3.97E-02 (3.64E-02,4.30E-02) | 0.83(0.77,0.89) | 0.62(0.57,0.67) |
| Glutamine (mmol/l) | -5.55E-02 (-5.97E-02,-5.12E-02) | -1.45E-01 (-1.48E-01,-1.41E-01) | -0.96(-10.03,-0.88) | -20.49(-20.56,-20.43) |
| Histidine (mmol/l) | -2.10E-02 (-2.16E-02,-2.04E-02) | -2.66E-02 (-2.72E-02,-2.60E-02) | -10.47(-10.51,-10.42) | -10.86(-10.9,-10.81) |

**eTable 4: Mean absolute difference in traits between 7y and 25y in males and females**

| | Original units | | SD units | |
|---|---|---|---|---|
| | Mean absolute difference between 7y and 25y (95% CI) | | Mean absolute difference between 7y and 25y (95% CI) | |
| | Males | Females | Males | Females |
| Isoleucine (mmol/l) | 3.01E-03 (2.08E-03,3.94E-03) | -8.71E-03 (-9.52E-03,-7.90E-03) | 0.17(0.12,0.22) | -0.48(-0.53,-0.44) |
| Leucine (mmol/l) | 9.37E-03 (8.52E-03,1.02E-02) | -2.61E-03 (-3.33E-03,-1.89E-03) | 0.62(0.57,0.68) | -0.17(-0.22,-0.13) |
| Valine (mmol/l) | 1.53E-02 (1.34E-02,1.73E-02) | -1.17E-02 (-1.34E-02,-9.99E-03) | 0.44(0.38,0.49) | -0.33(-0.38,-0.29) |
| Phenylalanine (mmol/l) | 6.10E-03 (5.65E-03,6.55E-03) | 4.39E-03 (3.98E-03,4.80E-03) | 0.71(0.66,0.76) | 0.51(0.46,0.56) |
| Tyrosine (mmol/l) | -2.07E-02 (-2.15E-02,-2.00E-02) | -2.63E-02 (-2.70E-02,-2.55E-02) | -10.31(-10.35,-10.26) | -10.66(-10.71,-10.61) |
| Acetate (mmol/l) | -6.45E-03 (-8.73E-03,-4.17E-03) | -1.30E-02 (-1.43E-02,-1.17E-02) | -0.27(-0.36,-0.17) | -0.54(-0.59,-0.49) |
| Acetoacetate (mmol/l) | -1.91E-02 (-2.10E-02,-1.72E-02) | -2.30E-02 (-2.50E-02,-2.10E-02) | -0.4(-0.44,-0.36) | -0.49(-0.53,-0.44) |
| 3-hydroxybutyrate (mmol/l) | 5.34E-02 (4.84E-02,5.84E-02) | 5.75E-02 (5.17E-02,6.33E-02) | 0.55(0.5,0.6) | 0.59(0.53,0.65) |
| Creatinine (mmol/l) | 2.92E-02 (2.87E-02,2.96E-02) | 1.68E-02 (1.65E-02,1.71E-02) | 40.77(40.7,40.84) | 20.75(20.7,20.81) |
| Albumin (mmol/l) | 4.28E-03 (3.99E-03,4.57E-03) | 3.09E-04 (4.88E-05,5.70E-04) | 10.18(10.1,10.27) | 0.09(0.01,0.16) |
| Glycoprotein acetyls, mainly a1-acid glycoprotein (mmol/l) | 6.33E-03 (-3.30E-03,1.60E-02) | -8.46E-03 (-1.67E-02,-1.73E-04) | 0.05(-0.02,0.12) | -0.06(-0.12,0) |

*These metabolites (diacylglycerol, fatty acid chain length, estimated degree of saturation and conjugated linoleic acid) were not measured at 25y; all models include data only up to aged 18y and values in this table for these traits are at 7y and 18y respectively. HDL: high-density lipoprotein; IDL: intermediate-density lipoprotein; LDL: low-density lipoprotein; VLDL: very-low-density lipoprotein.

| eTable 5: Mean rate of change in concentration per year in males and females | | | | |
|---|---|---|---|---|
| | Males | | Females | |
| | Mean change in concentration per year from 7 to 15y/ 18y(95%CI)* | Mean change in concentration per year from 15y/18y to 25y (95%CI)* | Mean change in concentration per year from 7 to 15y/ 18y(95%CI)* | Mean change in concentration per year from 15y/18y to 25y (95%CI)* |
| Concentration of chylomicrons and extremely large VLDL particles (mol/l) | -3.48E-12 (-4.11E-12,-2.85E-12) | -1.80E-13 (-7.41E-13,3.81E-13) | -6.18E-12 (-6.76E-12,-5.59E-12) | -2.58E-12 (-2.93E-12,-2.22E-12) |
| Total lipids in chylomicrons and extremely large VLDL (mmol/l) | -7.84E-04 (-9.25E-04,-6.43E-04) | -4.32E-04 (-5.55E-04,-3.10E-04) | -1.35E-03 (-1.49E-03,-1.22E-03) | -9.43E-04 (-1.02E-03,-8.65E-04) |
| Phospholipids in chylomicrons and extremely large VLDL (mmol/l) | -1.01E-04 (-1.18E-04,-8.35E-05) | -6.28E-05 (-7.81E-05,-4.76E-05) | -1.65E-04 (-1.81E-04,-1.49E-04) | -1.25E-04 (-1.35E-04,-1.16E-04) |
| Total cholesterol in chylomicrons and extremely large VLDL (mmol/l) | -8.79E-05 (-1.05E-04,-7.06E-05) | -9.34E-05 (-1.21E-04,-6.54E-05) | -1.69E-04 (-1.85E-04,-1.53E-04) | -1.68E-04 (-1.87E-04,-1.50E-04) |
| Cholesterol esters in chylomicrons and extremely large VLDL (mmol/l) | -3.52E-05 (-4.42E-05,-2.63E-05) | -3.71E-05 (-5.21E-05,-2.21E-05) | -7.15E-05 (-7.97E-05,-6.33E-05) | -8.00E-05 (-9.02E-05,-6.97E-05) |
| Free cholesterol in chylomicrons and extremely large VLDL (mmol/l) | -6.13E-05 (-7.28E-05,-4.98E-05) | -4.70E-05 (-5.70E-05,-3.70E-05) | -1.03E-04 (-1.14E-04,-9.27E-05) | -8.69E-05 (-9.33E-05,-8.05E-05) |
| Triglycerides in chylomicrons and extremely large VLDL (mmol/l) | -5.68E-04 (-6.70E-04,-4.65E-04) | -3.05E-04 (-3.92E-04,-2.18E-04) | -1.00E-03 (-1.10E-03,-9.07E-04) | -6.64E-04 (-7.19E-04,-6.08E-04) |
| Concentration of very large VLDL particles (mol/l) | -1.82E-11 (-2.17E-11,-1.47E-11) | 1.20E-12 (-2.10E-12,4.50E-12) | -3.37E-11 (-3.69E-11,-3.05E-11) | -1.22E-11 (-1.43E-11,-1.02E-11) |
| Total lipids in very large VLDL (mmol/l) | -1.83E-03 (-2.17E-03,-1.49E-03) | -7.20E-05 (-3.93E-04,2.49E-04) | -3.26E-03 (-3.57E-03,-2.94E-03) | -1.41E-03 (-1.61E-03,-1.21E-03) |
| Phospholipids in very large VLDL (mmol/l) | -3.47E-04 (-4.03E-04,-2.90E-04) | 1.69E-05 (-3.64E-05,7.01E-05) | -5.48E-04 (-6.01E-04,-4.96E-04) | -2.12E-04 (-2.45E-04,-1.79E-04) |
| Total cholesterol in very large VLDL (mmol/l) | -3.16E-04 (-3.82E-04,-2.50E-04) | -1.97E-04 (-2.58E-04,-1.35E-04) | -5.91E-04 (-6.52E-04,-5.30E-04) | -4.64E-04 (-5.03E-04,-4.25E-04) |
| Cholesterol esters in very large VLDL (mmol/l) | -1.03E-04 (-1.29E-04,-7.68E-05) | -1.53E-04 (-1.97E-04,-1.10E-04) | -2.70E-04 (-2.94E-04,-2.46E-04) | -2.59E-04 (-2.87E-04,-2.31E-04) |
| Free cholesterol in very large VLDL (mmol/l) | -1.81E-04 (-2.13E-04,-1.48E-04) | -9.48E-05 (-1.24E-04,-6.57E-05) | -2.96E-04 (-3.26E-04,-2.65E-04) | -2.23E-04 (-2.41E-04,-2.04E-04) |
| Triglycerides in very large VLDL (mmol/l) | -1.16E-03 (-1.38E-03,-9.44E-04) | 1.08E-04 (-9.96E-05,3.15E-04) | -2.12E-03 (-2.32E-03,-1.92E-03) | -7.36E-04 (-8.66E-04,-6.06E-04) |
| Concentration of large VLDL particles (mol/l) | -9.61E-11 (-1.13E-10,-7.95E-11) | 1.07E-10 (8.79E-11,1.25E-10) | -1.62E-10 (-1.77E-10,-1.47E-10) | 1.72E-12 (-1.01E-11,1.35E-11) |
| Total lipids in large VLDL (mmol/l) | -5.64E-03 (-6.63E-03,-4.65E-03) | 5.10E-03 (4.01E-03,6.19E-03) | -9.56E-03 (-1.05E-02,-8.66E-03) | -1.02E-03 (-1.71E-03,-3.25E-04) |
| Phospholipids in large VLDL (mmol/l) | -1.14E-03 (-1.31E-03,-9.59E-04) | 1.16E-03 (9.63E-04,1.36E-03) | -1.77E-03 (-1.93E-03,-1.61E-03) | 5.08E-05 (-7.57E-05,1.77E-04) |
| Total cholesterol in large VLDL (mmol/l) | -1.40E-03 (-1.62E-03,-1.17E-03) | 8.61E-04 (6.18E-04,1.10E-03) | -2.14E-03 (-2.35E-03,-1.94E-03) | -5.93E-04 (-7.48E-04,-4.39E-04) |
| Cholesterol esters in large VLDL (mmol/l) | -6.43E-04 (-7.48E-04,-5.38E-04) | 5.77E-04 (4.59E-04,6.95E-04) | -9.71E-04 (-1.07E-03,-8.74E-04) | -2.36E-04 (-3.13E-04,-1.59E-04) |
| Free cholesterol in large VLDL (mmol/l) | -7.51E-04 (-8.71E-04,-6.32E-04) | 2.82E-04 (1.57E-04,4.08E-04) | -1.17E-03 (-1.28E-03,-1.06E-03) | -3.57E-04 (-4.36E-04,-2.78E-04) |
| Triglycerides in large VLDL (mmol/l) | -3.11E-03 (-3.70E-03,-2.51E-03) | 3.08E-03 (2.43E-03,3.73E-03) | -5.65E-03 (-6.19E-03,-5.11E-03) | -4.76E-04 (-8.90E-04,-6.28E-05) |
| Concentration of medium VLDL particles (mol/l) | -1.98E-10 (-2.34E-10,-1.61E-10) | 4.05E-10 (3.58E-10,4.52E-10) | -3.58E-10 (-3.91E-10,-3.24E-10) | 5.89E-11 (2.86E-11,8.91E-11) |
| Total lipids in medium VLDL (mmol/l) | -6.58E-03 (-7.83E-03,-5.33E-03) | 1.12E-02 (9.64E-03,1.27E-02) | -1.17E-02 (-1.28E-02,-1.05E-02) | -5.47E-04 (-1.56E-03,4.70E-04) |
| Phospholipids in medium VLDL (mmol/l) | -1.58E-03 (-1.81E-03,-1.34E-03) | 2.29E-03 (2.00E-03,2.59E-03) | -2.37E-03 (-2.59E-03,-2.14E-03) | -4.23E-05 (-2.39E-04,1.55E-04) |
| Total cholesterol in medium VLDL (mmol/l) | -2.62E-03 (-2.94E-03,-2.30E-03) | 3.13E-03 (2.73E-03,3.52E-03) | -3.21E-03 (-3.51E-03,-2.91E-03) | 2.97E-05 (-2.42E-04,3.01E-04) |
| Cholesterol esters in medium VLDL (mmol/l) | -1.57E-03 (-1.74E-03,-1.40E-03) | 2.09E-03 (1.88E-03,2.30E-03) | -1.70E-03 (-1.87E-03,-1.54E-03) | 4.65E-04 (3.15E-04,6.14E-04) |
| Free cholesterol in medium VLDL (mmol/l) | -1.04E-03 (-1.20E-03,-8.85E-04) | 1.03E-03 (8.39E-04,1.23E-03) | -1.50E-03 (-1.65E-03,-1.36E-03) | -4.41E-04 (-5.69E-04,-3.12E-04) |
| Triglycerides in medium VLDL (mmol/l) | -2.37E-03 (-3.08E-03,-1.67E-03) | 5.75E-03 (4.88E-03,6.62E-03) | -6.07E-03 (-6.71E-03,-5.43E-03) | -5.43E-04 (-1.10E-03,1.79E-05) |
| Concentration of small VLDL particles (mol/l) | -2.93E-10 (-3.28E-10,-2.58E-10) | 2.21E-10 (1.74E-10,2.68E-10) | -3.78E-10 (-4.13E-10,-3.43E-10) | -2.09E-10 (-2.43E-10,-1.74E-10) |
| Total lipids in small VLDL (mmol/l) | -3.67E-03 (-4.20E-03,-3.13E-03) | -2.89E-03 (-4.08E-03,-1.69E-03) | -6.28E-03 (-6.83E-03,-5.73E-03) | -1.35E-02 (-1.44E-02,-1.26E-02) |
| Phospholipids in small VLDL (mmol/l) | -1.38E-03 (-1.51E-03,-1.24E-03) | -1.87E-04 (-3.66E-04,-8.70E-06) | -1.46E-03 (-1.61E-03,-1.32E-03) | -1.79E-03 (-1.94E-03,-1.65E-03) |
| Total cholesterol in small VLDL (mmol/l) | -1.44E-03 (-1.62E-03,-1.25E-03) | -5.10E-03 (-5.52E-03,-4.68E-03) | -2.22E-03 (-2.42E-03,-2.01E-03) | -8.90E-03 (-9.26E-03,-8.55E-03) |
| Cholesterol esters in small VLDL (mmol/l) | -6.73E-04 (-7.98E-04,-5.48E-04) | -4.25E-03 (-4.53E-03,-3.97E-03) | -1.28E-03 (-1.42E-03,-1.14E-03) | -6.56E-03 (-6.81E-03,-6.32E-03) |
| Free cholesterol in small VLDL (mmol/l) | -7.66E-04 (-8.38E-04,-6.95E-04) | -8.52E-04 (-1.01E-03,-6.93E-04) | -9.33E-04 (-1.01E-03,-8.57E-04) | -2.34E-03 (-2.47E-03,-2.21E-03) |
| Triglycerides in small VLDL (mmol/l) | -2.16E-03 (-2.53E-03,-1.80E-03) | 2.13E-03 (1.67E-03,2.58E-03) | -3.32E-03 (-3.67E-03,-2.97E-03) | -1.89E-03 (-2.20E-03,-1.57E-03) |
| Concentration of very small VLDL particles (mol/l) | -4.17E-10 (-4.42E-10,-3.92E-10) | 1.48E-10 (1.08E-10,1.89E-10) | -2.95E-10 (-3.24E-10,-2.66E-10) | -9.82E-11 (-1.34E-10,-6.21E-11) |
| Total lipids in very small VLDL (mmol/l) | -3.75E-03 (-4.05E-03,-3.45E-03) | -1.06E-02 (-1.13E-02,-9.92E-03) | -3.78E-03 (-4.13E-03,-3.44E-03) | -1.48E-02 (-1.54E-02,-1.41E-02) |
| Phospholipids in very small VLDL (mmol/l) | -2.10E-03 (-2.22E-03,-1.97E-03) | 5.32E-04 (3.61E-04,7.04E-04) | -1.21E-03 (-1.35E-03,-1.06E-03) | -5.70E-04 (-7.29E-04,-4.10E-04) |
| Total cholesterol in very small VLDL (mmol/l) | -1.17E-03 (-1.37E-03,-9.79E-04) | -1.11E-02 (-1.15E-02,-1.07E-02) | -1.95E-03 (-2.16E-03,-1.75E-03) | -1.19E-02 (-1.23E-02,-1.16E-02) |
| Cholesterol esters in very small VLDL (mmol/l) | -4.68E-04 (-6.04E-04,-3.33E-04) | -8.95E-03 (-9.21E-03,-8.68E-03) | -1.40E-03 (-1.55E-03,-1.25E-03) | -8.89E-03 (-9.14E-03,-8.64E-03) |
| Free cholesterol in very small VLDL (mmol/l) | -7.13E-04 (-7.79E-04,-6.46E-04) | -2.13E-03 (-2.25E-03,-2.00E-03) | -5.56E-04 (-6.28E-04,-4.84E-04) | -3.02E-03 (-3.13E-03,-2.90E-03) |
| Triglycerides in very small VLDL (mmol/l) | -9.96E-04 (-1.09E-03,-9.04E-04) | -5.76E-04 (-7.61E-04,-3.90E-04) | -8.71E-04 (-9.73E-04,-7.69E-04) | -2.12E-03 (-2.29E-03,-1.96E-03) |
| Concentration of IDL particles (mol/l) | -1.10E-09 (-1.16E-09,-1.05E-09) | 2.43E-09 (2.28E-09,2.57E-09) | -6.93E-10 (-7.56E-10,-6.29E-10) | 1.77E-09 (1.63E-09,1.91E-09) |
| Total lipids in IDL (mmol/l) | -1.33E-02 (-1.40E-02,-1.26E-02) | 4.04E-03 (2.48E-03,5.60E-03) | -8.22E-03 (-9.02E-03,-7.42E-03) | -6.77E-03 (-8.27E-03,-5.27E-03) |

| eTable 5: Mean rate of change in concentration per year in males and females | | | | |
|---|---|---|---|---|
| | Males | | Females | |
| | **Mean change in concentration per year from 7 to 15y/ 18y(95%CI)*** | **Mean change in concentration per year from 15y/18y to 25y (95%CI)*** | **Mean change in concentration per year from 7 to 15y/ 18y(95%CI)*** | **Mean change in concentration per year from 15y/18y to 25y (95%CI)*** |
| Phospholipids in IDL (mmol/l) | -3.56E-03 (-3.74E-03,-3.37E-03) | 3.08E-03 (2.69E-03,3.48E-03) | -1.86E-03 (-2.08E-03,-1.64E-03) | 1.97E-04 (-1.87E-04,5.81E-04) |
| Total cholesterol in IDL (mmol/l) | -8.18E-03 (-8.65E-03,-7.71E-03) | 1.36E-03 (2.93E-04,2.43E-03) | -5.82E-03 (-6.35E-03,-5.30E-03) | -5.21E-03 (-6.22E-03,-4.20E-03) |
| Cholesterol esters in IDL (mmol/l) | -5.44E-03 (-5.78E-03,-5.10E-03) | -6.70E-05 (-8.45E-04,7.11E-04) | -4.35E-03 (-4.72E-03,-3.98E-03) | -4.65E-03 (-5.37E-03,-3.92E-03) |
| Free cholesterol in IDL (mmol/l) | -2.75E-03 (-2.89E-03,-2.60E-03) | 1.44E-03 (1.13E-03,1.74E-03) | -1.47E-03 (-1.64E-03,-1.31E-03) | -5.66E-04 (-8.60E-04,-2.72E-04) |
| Triglycerides in IDL (mmol/l) | -1.59E-03 (-1.69E-03,-1.50E-03) | -3.91E-04 (-5.57E-04,-2.26E-04) | -5.13E-04 (-6.26E-04,-3.99E-04) | -1.76E-03 (-1.94E-03,-1.57E-03) |
| Concentration of large LDL particles (mol/l) | -1.98E-09 (-2.08E-09,-1.88E-09) | 7.32E-09 (7.04E-09,7.60E-09) | -1.01E-09 (-1.13E-09,-8.88E-10) | 5.46E-09 (5.21E-09,5.71E-09) |
| Total lipids in large LDL (mmol/l) | -1.80E-02 (-1.89E-02,-1.72E-02) | 2.83E-02 (2.63E-02,3.03E-02) | -9.53E-03 (-1.06E-02,-8.49E-03) | 8.93E-03 (7.04E-03,1.08E-02) |
| Phospholipids in large LDL (mmol/l) | -3.44E-03 (-3.62E-03,-3.26E-03) | 6.37E-03 (5.95E-03,6.79E-03) | -1.78E-03 (-1.99E-03,-1.57E-03) | 2.68E-03 (2.29E-03,3.07E-03) |
| Total cholesterol in large LDL (mmol/l) | -1.27E-02 (-1.34E-02,-1.21E-02) | 2.23E-02 (2.08E-02,2.38E-02) | -7.23E-03 (-7.97E-03,-6.48E-03) | 7.66E-03 (6.27E-03,9.04E-03) |
| Cholesterol esters in large LDL (mmol/l) | -9.62E-03 (-1.01E-02,-9.13E-03) | 1.77E-02 (1.66E-02,1.89E-02) | -5.63E-03 (-6.19E-03,-5.07E-03) | 6.07E-03 (5.02E-03,7.12E-03) |
| Free cholesterol in large LDL (mmol/l) | -3.13E-03 (-3.29E-03,-2.96E-03) | 4.53E-03 (4.18E-03,4.89E-03) | -1.59E-03 (-1.78E-03,-1.41E-03) | 1.59E-03 (1.25E-03,1.92E-03) |
| Triglycerides in large LDL (mmol/l) | -1.87E-03 (-1.97E-03,-1.77E-03) | -2.99E-04 (-4.57E-04,-1.40E-04) | -5.02E-04 (-6.25E-04,-3.79E-04) | -1.39E-03 (-1.58E-03,-1.21E-03) |
| Concentration of medium LDL particles (mol/l) | -1.68E-09 (-1.78E-09,-1.59E-09) | 6.92E-09 (6.66E-09,7.18E-09) | -8.88E-10 (-9.97E-10,-7.80E-10) | 4.94E-09 (4.71E-09,5.17E-09) |
| Total lipids in medium LDL (mmol/l) | -1.11E-02 (-1.16E-02,-1.05E-02) | 2.30E-02 (2.16E-02,2.43E-02) | -6.03E-03 (-6.68E-03,-5.37E-03) | 9.03E-03 (7.79E-03,1.03E-02) |
| Phospholipids in medium LDL (mmol/l) | -3.39E-03 (-3.53E-03,-3.24E-03) | 2.86E-03 (2.66E-03,3.07E-03) | -1.98E-03 (-2.14E-03,-1.82E-03) | 1.20E-03 (1.02E-03,1.39E-03) |
| Total cholesterol in medium LDL (mmol/l) | -7.78E-03 (-8.19E-03,-7.36E-03) | 1.78E-02 (1.67E-02,1.88E-02) | -4.61E-03 (-5.08E-03,-4.13E-03) | 7.15E-03 (6.22E-03,8.08E-03) |
| Cholesterol esters in medium LDL (mmol/l) | -6.13E-03 (-6.48E-03,-5.79E-03) | 1.56E-02 (1.48E-02,1.65E-02) | -3.60E-03 (-3.99E-03,-3.20E-03) | 6.88E-03 (6.13E-03,7.62E-03) |
| Free cholesterol in medium LDL (mmol/l) | -1.65E-03 (-1.72E-03,-1.57E-03) | 2.12E-03 (1.92E-03,2.33E-03) | -1.00E-03 (-1.09E-03,-9.18E-04) | 2.70E-04 (8.05E-05,4.60E-04) |
| Triglycerides in medium LDL (mmol/l) | -1.01E-03 (-1.08E-03,-9.46E-04) | 9.56E-04 (8.66E-04,1.05E-03) | -1.26E-04 (-2.04E-04,-4.81E-05) | 2.75E-04 (1.72E-04,3.78E-04) |
| Concentration of small LDL particles (mol/l) | -2.15E-09 (-2.25E-09,-2.05E-09) | 6.61E-09 (6.29E-09,6.92E-09) | -1.30E-09 (-1.42E-09,-1.18E-09) | 4.54E-09 (4.26E-09,4.82E-09) |
| Total lipids in small LDL (mmol/l) | -7.41E-03 (-7.77E-03,-7.06E-03) | 1.25E-02 (1.17E-02,1.34E-02) | -4.45E-03 (-4.86E-03,-4.03E-03) | 4.28E-03 (3.45E-03,5.10E-03) |
| Phospholipids in small LDL (mmol/l) | -2.55E-03 (-2.65E-03,-2.45E-03) | 1.41E-03 (1.25E-03,1.57E-03) | -1.63E-03 (-1.74E-03,-1.52E-03) | 5.42E-04 (3.96E-04,6.88E-04) |
| Total cholesterol in small LDL (mmol/l) | -4.96E-03 (-5.22E-03,-4.70E-03) | 9.55E-03 (8.90E-03,1.02E-02) | -3.04E-03 (-3.34E-03,-2.74E-03) | 3.27E-03 (2.67E-03,3.86E-03) |
| Cholesterol esters in small LDL (mmol/l) | -3.92E-03 (-4.14E-03,-3.70E-03) | 9.04E-03 (8.52E-03,9.56E-03) | -2.31E-03 (-2.57E-03,-2.05E-03) | 3.76E-03 (3.29E-03,4.23E-03) |
| Free cholesterol in small LDL (mmol/l) | -1.05E-03 (-1.09E-03,-9.98E-04) | 5.14E-04 (3.69E-04,6.58E-04) | -7.20E-04 (-7.74E-04,-6.67E-04) | -5.06E-04 (-6.41E-04,-3.72E-04) |
| Triglycerides in small LDL (mmol/l) | -8.23E-04 (-8.79E-04,-7.68E-04) | 3.92E-04 (3.44E-04,4.41E-04) | -3.99E-04 (-4.59E-04,-3.39E-04) | 1.68E-04 (1.21E-04,2.14E-04) |
| Concentration of very large HDL particles (mol/l) | -8.69E-09 (-9.30E-09,-8.07E-09) | -1.71E-08 (-1.79E-08,-1.63E-08) | -9.70E-10 (-1.65E-09,-2.95E-10) | -5.68E-09 (-6.59E-09,-4.77E-09) |
| Total lipids in very large HDL (mmol/l) | -9.16E-03 (-9.91E-03,-8.41E-03) | -3.14E-02 (-3.23E-02,-3.06E-02) | -4.20E-04 (-1.24E-03,3.96E-04) | -2.06E-02 (-2.16E-02,-1.97E-02) |
| Phospholipids in very large HDL (mmol/l) | -5.18E-03 (-5.57E-03,-4.79E-03) | -8.79E-03 (-9.25E-03,-8.34E-03) | 5.53E-04 (1.28E-04,9.77E-04) | -2.67E-03 (-3.19E-03,-2.15E-03) |
| Total cholesterol in very large HDL (mmol/l) | -6.97E-03 (-7.26E-03,-6.67E-03) | -2.59E-02 (-2.65E-02,-2.54E-02) | -3.79E-03 (-4.11E-03,-3.48E-03) | -2.07E-02 (-2.13E-02,-2.01E-02) |
| Cholesterol esters in very large HDL (mmol/l) | -5.07E-03 (-5.29E-03,-4.85E-03) | -2.08E-02 (-2.12E-02,-2.03E-02) | -3.05E-03 (-3.28E-03,-2.81E-03) | -1.71E-02 (-1.76E-02,-1.67E-02) |
| Free cholesterol in very large HDL (mmol/l) | -1.25E-03 (-1.35E-03,-1.15E-03) | -4.54E-03 (-4.65E-03,-4.43E-03) | -1.81E-04 (-2.92E-04,-7.01E-05) | -3.07E-03 (-3.19E-03,-2.94E-03) |
| Triglycerides in very large HDL (mmol/l) | -3.76E-04 (-4.08E-04,-3.44E-04) | -7.01E-04 (-7.34E-04,-6.67E-04) | -2.01E-04 (-2.34E-04,-1.69E-04) | -6.44E-04 (-6.77E-04,-6.12E-04) |
| Concentration of large HDL particles (mol/l) | -2.10E-08 (-2.19E-08,-2.01E-08) | 2.66E-09 (5.96E-11,5.26E-09) | 1.59E-09 (5.56E-10,2.62E-09) | 3.88E-08 (3.58E-08,4.18E-08) |
| Total lipids in large HDL (mmol/l) | -1.54E-02 (-1.61E-02,-1.47E-02) | -1.09E-02 (-1.26E-02,-9.22E-03) | 1.13E-03 (3.56E-04,1.90E-03) | 8.99E-03 (7.06E-03,1.09E-02) |
| Phospholipids in large HDL (mmol/l) | -7.07E-03 (-7.37E-03,-6.77E-03) | -1.88E-03 (-2.65E-03,-1.12E-03) | 7.68E-04 (4.20E-04,1.12E-03) | 6.25E-03 (5.38E-03,7.13E-03) |
| Total cholesterol in large HDL (mmol/l) | -7.97E-03 (-8.34E-03,-7.59E-03) | -9.03E-03 (-9.94E-03,-8.13E-03) | 4.95E-04 (7.30E-05,9.17E-04) | 1.74E-03 (7.20E-04,2.76E-03) |
| Cholesterol esters in large HDL (mmol/l) | -6.23E-03 (-6.52E-03,-5.93E-03) | -6.95E-03 (-7.64E-03,-6.26E-03) | 4.74E-04 (1.41E-04,8.07E-04) | 9.32E-04 (1.49E-04,1.71E-03) |
| Free cholesterol in large HDL (mmol/l) | -1.74E-03 (-1.82E-03,-1.66E-03) | -2.08E-03 (-2.30E-03,-1.86E-03) | 2.04E-05 (-6.94E-05,1.10E-04) | 8.11E-04 (5.71E-04,1.05E-03) |
| Triglycerides in large HDL (mmol/l) | -3.73E-04 (-3.89E-04,-3.57E-04) | 2.85E-05 (-3.51E-05,9.21E-05) | -1.38E-04 (-1.55E-04,-1.20E-04) | 1.02E-03 (9.38E-04,1.10E-03) |
| Concentration of medium HDL particles (mol/l) | -1.43E-08 (-1.51E-08,-1.34E-08) | 8.88E-08 (8.59E-08,9.17E-08) | 5.23E-09 (4.16E-09,6.29E-09) | 1.04E-07 (1.01E-07,1.07E-07) |
| Total lipids in medium HDL (mmol/l) | -1.09E-02 (-1.15E-02,-1.04E-02) | 9.80E-03 (8.86E-03,1.07E-02) | -7.53E-04 (-1.42E-03,-9.00E-05) | 1.63E-02 (1.52E-02,1.73E-02) |
| Phospholipids in medium HDL (mmol/l) | -5.56E-03 (-5.86E-03,-5.26E-03) | 2.63E-03 (2.21E-03,3.04E-03) | 2.76E-04 (-8.66E-05,6.38E-04) | 5.10E-03 (4.62E-03,5.59E-03) |
| Total cholesterol in medium HDL (mmol/l) | -3.00E-03 (-3.21E-03,-2.79E-03) | 8.50E-03 (7.79E-03,9.22E-03) | 1.13E-03 (8.83E-04,1.37E-03) | 1.19E-02 (1.11E-02,1.26E-02) |
| Cholesterol esters in medium HDL (mmol/l) | -2.29E-03 (-2.47E-03,-2.12E-03) | 6.55E-03 (5.99E-03,7.12E-03) | 1.05E-03 (8.50E-04,1.25E-03) | 8.87E-03 (8.26E-03,9.48E-03) |

**eTable 5: Mean rate of change in concentration per year in males and females**

| | Males | | Females | |
|---|---|---|---|---|
| | **Mean change in concentration per year from 7 to 15y/ 18y(95%CI)*** | **Mean change in concentration per year from 15y/18y to 25y (95%CI)*** | **Mean change in concentration per year from 7 to 15y/ 18y(95%CI)*** | **Mean change in concentration per year from 15y/18y to 25y (95%CI)*** |
| Free cholesterol in medium HDL (mmol/l) | -7.09E-04 (-7.47E-04,-6.71E-04) | 1.95E-03 (1.80E-03,2.11E-03) | 7.82E-05 (3.33E-05,1.23E-04) | 2.98E-03 (2.82E-03,3.15E-03) |
| Triglycerides in medium HDL (mmol/l) | -5.78E-04 (-6.28E-04,-5.28E-04) | 1.59E-03 (1.52E-03,1.66E-03) | -3.63E-04 (-4.16E-04,-3.11E-04) | 1.56E-03 (1.50E-03,1.63E-03) |
| Concentration of small HDL particles (mol/l) | -3.34E-08 (-3.53E-08,-3.15E-08) | 1.05E-07 (1.01E-07,1.08E-07) | -1.66E-08 (-1.87E-08,-1.44E-08) | 1.14E-07 (1.10E-07,1.18E-07) |
| Total lipids in small HDL (mmol/l) | -8.08E-03 (-8.61E-03,-7.56E-03) | 1.28E-02 (1.19E-02,1.37E-02) | 3.58E-04 (-2.85E-04,1.00E-03) | 1.05E-02 (9.48E-03,1.15E-02) |
| Phospholipids in small HDL (mmol/l) | -2.08E-03 (-2.39E-03,-1.77E-03) | 2.58E-04 (-1.29E-04,6.45E-04) | 5.88E-06 (-3.48E-04,3.60E-04) | 1.49E-03 (1.04E-03,1.94E-03) |
| Total cholesterol in small HDL (mmol/l) | -5.40E-03 (-5.70E-03,-5.09E-03) | 1.17E-02 (1.11E-02,1.22E-02) | 6.93E-04 (3.54E-04,1.03E-03) | 9.33E-03 (8.73E-03,9.92E-03) |
| Cholesterol esters in small HDL (mmol/l) | -4.66E-03 (-4.93E-03,-4.39E-03) | 1.27E-02 (1.22E-02,1.32E-02) | 5.09E-04 (2.21E-04,7.97E-04) | 1.05E-02 (9.96E-03,1.10E-02) |
| Free cholesterol in small HDL (mmol/l) | -7.00E-04 (-7.50E-04,-6.50E-04) | -1.53E-03 (-1.65E-03,-1.42E-03) | 9.54E-05 (3.21E-05,1.59E-04) | -1.99E-03 (-2.12E-03,-1.85E-03) |
| Triglycerides in small HDL (mmol/l) | -4.05E-04 (-4.60E-04,-3.50E-04) | 4.38E-04 (3.76E-04,5.00E-04) | -2.64E-04 (-3.22E-04,-2.07E-04) | 1.89E-05 (-3.56E-05,7.33E-05) |
| Mean diameter for VLDL particles (mm) | -1.92E-02 (-2.79E-02,-1.06E-02) | 4.87E-02 (4.05E-02,5.69E-02) | -7.13E-02 (-7.90E-02,-6.37E-02) | -1.88E-03 (-8.08E-03,4.32E-03) |
| Mean diameter for LDL particles (mm) | 1.53E-02 (1.45E-02,1.61E-02) | -2.01E-02 (-2.12E-02,-1.90E-02) | 9.00E-03 (8.26E-03,9.74E-03) | -1.06E-02 (-1.16E-02,-9.52E-03) |
| Mean diameter for HDL particles (mm) | -1.01E-02 (-1.10E-02,-9.31E-03) | -2.21E-02 (-2.32E-02,-2.09E-02) | 5.30E-04 (-3.42E-04,1.40E-03) | -6.83E-03 (-7.95E-03,-5.71E-03) |
| Serum total cholesterol (mmol/l) | -5.89E-02 (-6.12E-02,-5.65E-02) | 2.88E-02 (2.31E-02,3.45E-02) | -2.90E-02 (-3.18E-02,-2.63E-02) | -6.59E-03 (-1.22E-02,-9.76E-04) |
| Total cholesterol in VLDL (mmol/l) | -5.24E-03 (-5.95E-03,-4.54E-03) | -1.16E-02 (-1.31E-02,-1.01E-02) | -9.10E-03 (-9.81E-03,-8.40E-03) | -2.17E-02 (-2.28E-02,-2.05E-02) |
| Remnant cholesterol (non-HDL, non-LDL -cholesterol) (mmol/l) | -1.34E-02 (-1.44E-02,-1.24E-02) | -1.02E-02 (-1.26E-02,-7.89E-03) | -1.49E-02 (-1.60E-02,-1.38E-02) | -2.69E-02 (-2.89E-02,-2.49E-02) |
| Total cholesterol in LDL (mmol/l) | -2.55E-02 (-2.68E-02,-2.42E-02) | 4.96E-02 (4.64E-02,5.27E-02) | -1.49E-02 (-1.64E-02,-1.34E-02) | 1.81E-02 (1.52E-02,2.10E-02) |
| Total cholesterol in HDL (mmol/l) | -2.00E-02 (-2.08E-02,-1.92E-02) | -1.09E-02 (-1.29E-02,-8.83E-03) | 6.93E-04 (-2.76E-04,1.66E-03) | 2.20E-03 (-2.14E-05,4.43E-03) |
| Total cholesterol in HDL2 (mmol/l) | -1.15E-02 (-1.21E-02,-1.10E-02) | -4.88E-04 (-2.31E-03,1.33E-03) | 1.68E-03 (1.05E-03,2.32E-03) | 2.17E-02 (1.97E-02,2.36E-02) |
| Total cholesterol in HDL3 (mmol/l) | -8.59E-03 (-8.89E-03,-8.30E-03) | -9.92E-03 (-1.04E-02,-9.48E-03) | -1.31E-03 (-1.67E-03,-9.47E-04) | -1.81E-02 (-1.87E-02,-1.76E-02) |
| Esterified cholesterol (mmol/l) | -5.48E-02 (-5.71E-02,-5.25E-02) | 9.56E-03 (6.42E-03,1.27E-02) | -2.90E-02 (-3.15E-02,-2.64E-02) | -3.32E-03 (-6.29E-03,-3.42E-04) |
| Free cholesterol (mmol/l) | -1.63E-02 (-1.70E-02,-1.56E-02) | 6.63E-03 (4.94E-03,8.31E-03) | -7.04E-03 (-7.89E-03,-6.20E-03) | -3.39E-03 (-5.08E-03,-1.70E-03) |
| Serum total triglycerides (mmol/l) | -1.84E-02 (-2.08E-02,-1.60E-02) | 1.11E-02 (8.35E-03,1.38E-02) | -2.20E-02 (-2.43E-02,-1.98E-02) | -5.99E-03 (-7.88E-03,-4.10E-03) |
| Triglycerides in VLDL (mmol/l) | -1.06E-02 (-1.27E-02,-8.54E-03) | 1.03E-02 (7.92E-03,1.27E-02) | -1.91E-02 (-2.10E-02,-1.72E-02) | -5.93E-03 (-7.48E-03,-4.37E-03) |
| Triglycerides in LDL (mmol/l) | -3.48E-03 (-3.69E-03,-3.28E-03) | 1.29E-03 (9.87E-04,1.60E-03) | -8.64E-04 (-1.11E-03,-6.19E-04) | -9.47E-04 (-1.29E-03,-6.02E-04) |
| Triglycerides in HDL (mmol/l) | -1.77E-03 (-1.90E-03,-1.64E-03) | 1.24E-03 (1.08E-03,1.40E-03) | -1.12E-03 (-1.26E-03,-9.79E-04) | 1.68E-03 (1.51E-03,1.84E-03) |
| Diacylglycerol (mmol/l)* | -3.33E-04 (-4.11E-04,-2.56E-04) | 2.32E-04 (6.66E-06,4.56E-04) | -4.10E-04 (-4.93E-04,-3.27E-04) | 4.18E-04 (1.70E-04,6.66E-04) |
| Total phosphoglycerides (mmol/l) | -4.28E-02 (-4.43E-02,-4.13E-02) | -3.76E-03 (-5.47E-03,-2.05E-03) | -2.07E-02 (-2.25E-02,-1.88E-02) | -2.05E-03 (-3.99E-03,-1.04E-04) |
| Phosphatidylcholine and other cholines (mmol/l) | -2.65E-02 (-2.77E-02,-2.53E-02) | -1.17E-02 (-1.43E-02,-9.09E-03) | -5.41E-03 (-6.88E-03,-3.93E-03) | -2.51E-02 (-2.78E-02,-2.23E-02) |
| Total cholines (mmol/l) | -4.19E-02 (-4.36E-02,-4.02E-02) | -5.52E-03 (-7.60E-03,-3.45E-03) | -1.83E-02 (-2.02E-02,-1.63E-02) | -3.20E-03 (-5.43E-03,-9.79E-04) |
| Apolipoprotein A-I (g/l) | -1.33E-02 (-1.38E-02,-1.29E-02) | 4.18E-03 (2.91E-03,5.44E-03) | -2.12E-03 (-2.65E-03,-1.58E-03) | 8.27E-03 (6.85E-03,9.68E-03) |
| Apolipoprotein B (g/l) | -1.01E-02 (-1.08E-02,-9.34E-03) | 7.02E-03 (6.09E-03,7.96E-03) | -9.85E-03 (-1.06E-02,-9.12E-03) | -5.59E-04 (-1.31E-03,1.94E-04) |
| Total fatty acids (mmol/l) | -2.09E-01 (-2.19E-01,-2.00E-01) | -1.36E-02 (-2.53E-02,-1.99E-03) | -1.35E-01 (-1.46E-01,-1.25E-01) | -6.94E-02 (-8.00E-02,-5.88E-02) |
| Fatty acid length* | 3.08E-02 (2.72E-02,3.43E-02) | -2.97E-02 (-4.08E-02,-1.86E-02) | 2.07E-02 (1.74E-02,2.41E-02) | -1.80E-03 (-1.19E-02,8.33E-03) |
| Estimated degree of unsaturation* | 4.04E-03 (3.45E-03,4.62E-03) | -5.02E-03 (-6.99E-03,-3.06E-03) | 5.45E-03 (4.87E-03,6.02E-03) | -4.94E-03 (-6.66E-03,-3.22E-03) |
| 22:6, docosahexaenoic acid (mmol/l) | -3.04E-03 (-3.21E-03,-2.87E-03) | 1.87E-03 (1.67E-03,2.06E-03) | -1.23E-03 (-1.43E-03,-1.02E-03) | 1.82E-03 (1.61E-03,2.03E-03) |
| 18:2, linoleic acid (mmol/l) | -6.69E-02 (-6.93E-02,-6.44E-02) | -7.25E-04 (-3.92E-03,2.47E-03) | -4.15E-02 (-4.41E-02,-3.89E-02) | -2.07E-02 (-2.35E-02,-1.79E-02) |
| Conjugated linoleic acid (mmol/l)* | -8.57E-04 (-1.02E-03,-6.90E-04) | -2.96E-04 (-7.78E-04,1.85E-04) | -1.03E-03 (-1.20E-03,-8.63E-04) | -5.71E-05 (-5.17E-04,4.03E-04) |
| Omega-3 fatty acids (mmol/l) | -7.65E-03 (-8.11E-03,-7.20E-03) | 2.77E-03 (2.26E-03,3.27E-03) | -4.66E-03 (-5.14E-03,-4.18E-03) | -3.36E-04 (-8.13E-04,1.41E-04) |
| Omega-6 fatty acids (mmol/l) | -7.41E-02 (-7.68E-02,-7.14E-02) | -4.53E-03 (-8.04E-03,-1.03E-03) | -4.32E-02 (-4.61E-02,-4.02E-02) | -2.21E-02 (-2.53E-02,-1.88E-02) |
| Polyunsaturated fatty acids (mmol/l) | -8.18E-02 (-8.47E-02,-7.88E-02) | -1.77E-03 (-5.63E-03,2.09E-03) | -4.79E-02 (-5.11E-02,-4.47E-02) | -2.24E-02 (-2.60E-02,-1.88E-02) |
| Monounsaturated fatty acids; 16:1, 18:1 (mmol/l) | -1.73E-02 (-2.00E-02,-1.45E-02) | -4.25E-03 (-9.70E-03,1.19E-03) | -6.00E-03 (-8.91E-03,-3.09E-03) | -3.24E-02 (-3.73E-02,-2.76E-02) |
| Saturated fatty acids (mmol/l) | -1.04E-01 (-1.08E-01,-1.00E-01) | -9.71E-03 (-1.43E-02,-5.09E-03) | -7.91E-02 (-8.35E-02,-7.47E-02) | -2.70E-02 (-3.12E-02,-2.28E-02) |
| Glucose (mmol/l) | 2.00E-02 (1.66E-02,2.34E-02) | -3.64E-02 (-3.97E-02,-3.31E-02) | 1.16E-02 (8.45E-03,1.48E-02) | -4.17E-02 (-4.40E-02,-3.94E-02) |
| Lactate (mmol/l) | -2.29E-02 (-2.68E-02,-1.91E-02) | -2.47E-02 (-2.84E-02,-2.10E-02) | -2.65E-02 (-3.06E-02,-2.24E-02) | -2.90E-02 (-3.25E-02,-2.54E-02) |
| Citrate (mmol/l) | -2.49E-03 (-2.62E-03,-2.36E-03) | 9.25E-03 (9.01E-03,9.48E-03) | -3.34E-03 (-3.46E-03,-3.22E-03) | 9.99E-03 (9.78E-03,1.02E-02) |

**eTable 5: Mean rate of change in concentration per year in males and females**

| | Males | | Females | |
|---|---|---|---|---|
| | **Mean change in concentration per year from 7 to 15y/ 18y(95%CI)*** | **Mean change in concentration per year from 15y/18y to 25y (95%CI)*** | **Mean change in concentration per year from 7 to 15y/ 18y(95%CI)*** | **Mean change in concentration per year from 15y/18y to 25y (95%CI)*** |
| Alanine (mmol/l) | -4.59E-03 (-4.91E-03,-4.27E-03) | 1.73E-02 (1.68E-02,1.78E-02) | -5.20E-03 (-5.52E-03,-4.88E-03) | 1.62E-02 (1.57E-02,1.66E-02) |
| Glutamine (mmol/l) | 3.92E-03 (3.63E-03,4.21E-03) | -1.64E-02 (-1.71E-02,-1.58E-02) | -3.80E-03 (-4.12E-03,-3.48E-03) | -1.71E-02 (-1.77E-02,-1.65E-02) |
| Histidine (mmol/l) | -1.34E-04 (-2.01E-04,-6.67E-05) | -3.26E-03 (-3.36E-03,-3.15E-03) | -7.73E-04 (-8.44E-04,-7.02E-04) | -3.01E-03 (-3.11E-03,-2.92E-03) |
| Isoleucine (mmol/l) | -1.16E-04 (-2.02E-04,-2.98E-05) | 7.14E-04 (5.88E-04,8.39E-04) | -1.13E-03 (-1.21E-03,-1.05E-03) | 6.18E-04 (5.32E-04,7.04E-04) |
| Leucine (mmol/l) | 1.97E-04 (1.27E-04,2.68E-04) | 1.20E-03 (1.08E-03,1.32E-03) | -8.51E-04 (-9.18E-04,-7.83E-04) | 1.12E-03 (1.04E-03,1.21E-03) |
| Valine (mmol/l) | 1.99E-03 (1.76E-03,2.22E-03) | -6.33E-05 (-2.67E-04,1.41E-04) | -6.33E-04 (-8.48E-04,-4.17E-04) | -7.36E-04 (-9.00E-04,-5.72E-04) |
| Phenylalanine (mmol/l) | -5.33E-04 (-5.73E-04,-4.94E-04) | 1.99E-03 (1.93E-03,2.06E-03) | -7.01E-04 (-7.40E-04,-6.62E-04) | 2.02E-03 (1.97E-03,2.07E-03) |
| Tyrosine (mmol/l) | -1.07E-03 (-1.16E-03,-9.73E-04) | -1.35E-03 (-1.42E-03,-1.29E-03) | -1.52E-03 (-1.62E-03,-1.43E-03) | -1.57E-03 (-1.63E-03,-1.50E-03) |
| Acetate (mmol/l) | -2.12E-03 (-2.28E-03,-1.97E-03) | 1.17E-03 (8.98E-04,1.44E-03) | -2.47E-03 (-2.62E-03,-2.33E-03) | 7.48E-04 (6.27E-04,8.69E-04) |
| Acetoacetate (mmol/l) | -1.11E-03 (-1.23E-03,-9.95E-04) | | -2.22E-03 (-2.46E-03,-1.99E-03) | |
| 3-hydroxybutyrate (mmol/l) | 1.17E-03 (6.35E-04,1.71E-03) | 6.75E-03 (5.89E-03,7.61E-03) | 2.90E-03 (2.24E-03,3.57E-03) | 4.26E-03 (3.26E-03,5.26E-03) |
| Creatinine (mmol/l) | 3.34E-03 (3.30E-03,3.38E-03) | -1.27E-03 (-1.34E-03,-1.19E-03) | 2.26E-03 (2.23E-03,2.30E-03) | -1.35E-03 (-1.40E-03,-1.29E-03) |
| Albumin (mmol/l) | 1.01E-04 (8.04E-05,1.21E-04) | 5.28E-04 (4.80E-04,5.76E-04) | -1.92E-04 (-2.14E-04,-1.71E-04) | 4.04E-04 (3.61E-04,4.48E-04) |
| Glycoprotein acetyls, mainly a1-acid glycoprotein (mmol/l) | -3.32E-03 (-4.01E-03,-2.63E-03) | 7.13E-03 (5.68E-03,8.58E-03) | -5.11E-04 (-1.22E-03,2.00E-04) | -4.73E-04 (-1.68E-03,7.35E-04) |

*These metabolites (diacylglycerol, fatty acid chain length, estimated degree of saturation and conjugated linoleic acid) were not measured at 25y; all models include data only up to aged 18y and values in this table for these traits are at 7y and 18y respectively. HDL: high-density lipoprotein; IDL: intermediate-density lipoprotein; LDL: low-density lipoprotein; VLDL: very-low-density lipoprotein.

**eTable 6: Mean absolute difference in traits in SD units between 7y and 25y in males and females comparing i)findings from multilevel models implemented here to ii)linear regression of traits at 7y and 25y**

| | 7y | | 25y | |
|---|---|---|---|---|
| | Mean absolute sex difference in SD units (95% CI) | | Mean absolute sex difference in SD units (95% CI) | |
| | MLM | Regression | MLM | Regression |
| Concentration of chylomicrons and extremely large VLDL particles (mol/l) | 0.16(0.11,0.21) | 0.14(0.09,0.2) | -0.39(-0.45,-0.32) | -0.39(-0.46,-0.31) |
| Total lipids in chylomicrons and extremely large VLDL (mmol/l) | 0.15(0.09,0.2) | 0.13(0.08,0.19) | -0.39(-0.46,-0.32) | -0.38(-0.46,-0.31) |
| Phospholipids in chylomicrons and extremely large VLDL (mmol/l) | 0.15(0.09,0.2) | 0.13(0.08,0.19) | -0.36(-0.43,-0.29) | -0.35(-0.43,-0.28) |
| Total cholesterol in chylomicrons and extremely large VLDL (mmol/l) | 0.16(0.11,0.22) | 0.14(0.09,0.2) | -0.3(-0.37,-0.23) | -0.31(-0.39,-0.24) |
| Cholesterol esters in chylomicrons and extremely large VLDL (mmol/l) | 0.16(0.11,0.22) | 0.15(0.1,0.2) | -0.26(-0.33,-0.19) | -0.28(-0.35,-0.2) |
| Free cholesterol in chylomicrons and extremely large VLDL (mmol/l) | 0.15(0.09,0.2) | 0.13(0.08,0.19) | -0.35(-0.42,-0.28) | -0.35(-0.42,-0.27) |
| Triglycerides in chylomicrons and extremely large VLDL (mmol/l) | 0.15(0.09,0.2) | 0.13(0.08,0.18) | -0.41(-0.48,-0.34) | -0.4(-0.48,-0.33) |
| Concentration of very large VLDL particles (mol/l) | 0.14(0.08,0.19) | 0.12(0.07,0.17) | -0.4(-0.47,-0.33) | -0.4(-0.47,-0.32) |
| Total lipids in very large VLDL (mmol/l) | 0.14(0.08,0.19) | 0.12(0.07,0.17) | -0.4(-0.46,-0.33) | -0.39(-0.47,-0.32) |
| Phospholipids in very large VLDL (mmol/l) | 0.14(0.09,0.2) | 0.13(0.08,0.18) | -0.35(-0.42,-0.28) | -0.36(-0.43,-0.28) |
| Total cholesterol in very large VLDL (mmol/l) | 0.17(0.11,0.22) | 0.15(0.1,0.2) | -0.38(-0.45,-0.31) | -0.37(-0.45,-0.29) |
| Cholesterol esters in very large VLDL (mmol/l) | 0.18(0.12,0.23) | 0.15(0.1,0.2) | -0.38(-0.44,-0.31) | -0.39(-0.46,-0.31) |
| Free cholesterol in very large VLDL (mmol/l) | 0.16(0.11,0.21) | 0.15(0.09,0.2) | -0.35(-0.42,-0.28) | -0.35(-0.43,-0.27) |
| Triglycerides in very large VLDL (mmol/l) | 0.12(0.07,0.18) | 0.11(0.06,0.16) | -0.41(-0.48,-0.34) | -0.41(-0.48,-0.33) |
| Concentration of large VLDL particles (mol/l) | 0.14(0.08,0.19) | 0.12(0.07,0.17) | -0.43(-0.5,-0.37) | -0.45(-0.52,-0.37) |
| Total lipids in large VLDL (mmol/l) | 0.14(0.09,0.2) | 0.12(0.07,0.18) | -0.43(-0.5,-0.37) | -0.44(-0.52,-0.37) |
| Phospholipids in large VLDL (mmol/l) | 0.15(0.09,0.2) | 0.13(0.08,0.18) | -0.4(-0.47,-0.34) | -0.41(-0.49,-0.34) |
| Total cholesterol in large VLDL (mmol/l) | 0.16(0.11,0.22) | 0.15(0.1,0.2) | -0.4(-0.47,-0.34) | -0.41(-0.49,-0.34) |
| Cholesterol esters in large VLDL (mmol/l) | 0.18(0.13,0.24) | 0.17(0.12,0.22) | -0.42(-0.49,-0.36) | -0.44(-0.51,-0.36) |
| Free cholesterol in large VLDL (mmol/l) | 0.15(0.09,0.2) | 0.13(0.08,0.18) | -0.38(-0.45,-0.31) | -0.39(-0.46,-0.31) |
| Triglycerides in large VLDL (mmol/l) | 0.13(0.08,0.18) | 0.11(0.06,0.16) | -0.45(-0.52,-0.38) | -0.46(-0.53,-0.38) |
| Concentration of medium VLDL particles (mol/l) | 0.18(0.13,0.23) | 0.16(0.11,0.21) | -0.49(-0.56,-0.43) | -0.52(-0.59,-0.44) |
| Total lipids in medium VLDL (mmol/l) | 0.19(0.14,0.24) | 0.17(0.12,0.22) | -0.49(-0.55,-0.42) | -0.51(-0.58,-0.43) |
| Phospholipids in medium VLDL (mmol/l) | 0.2(0.15,0.26) | 0.19(0.13,0.24) | -0.46(-0.52,-0.39) | -0.49(-0.56,-0.41) |
| Total cholesterol in medium VLDL (mmol/l) | 0.23(0.18,0.29) | 0.22(0.17,0.28) | -0.36(-0.43,-0.3) | -0.39(-0.46,-0.31) |
| Cholesterol esters in medium VLDL (mmol/l) | 0.25(0.2,0.31) | 0.25(0.19,0.3) | -0.28(-0.35,-0.22) | -0.31(-0.39,-0.24) |
| Free cholesterol in medium VLDL (mmol/l) | 0.2(0.14,0.25) | 0.18(0.13,0.23) | -0.43(-0.5,-0.37) | -0.46(-0.53,-0.38) |
| Triglycerides in medium VLDL (mmol/l) | 0.16(0.11,0.21) | 0.14(0.09,0.19) | -0.55(-0.61,-0.48) | -0.56(-0.63,-0.49) |
| Concentration of small VLDL particles (mol/l) | 0.25(0.2,0.31) | 0.24(0.19,0.29) | -0.42(-0.48,-0.35) | -0.45(-0.52,-0.38) |
| Total lipids in small VLDL (mmol/l) | 0.29(0.24,0.35) | 0.28(0.23,0.33) | -0.41(-0.47,-0.35) | -0.43(-0.5,-0.36) |

**eTable 6: Mean absolute difference in traits in SD units between 7y and 25y in males and females comparing i)findings from multilevel models implemented here to ii)linear regression of traits at 7y and 25y**

| | 7y | | 25y | |
|---|---|---|---|---|
| | Mean absolute sex difference in SD units (95% CI) | | Mean absolute sex difference in SD units (95% CI) | |
| | MLM | Regression | MLM | Regression |
| Phospholipids in small VLDL (mmol/l) | 0.27(0.22,0.32) | 0.26(0.21,0.31) | -0.31(-0.37,-0.25) | -0.34(-0.41,-0.27) |
| Total cholesterol in small VLDL (mmol/l) | 0.37(0.32,0.42) | 0.36(0.3,0.41) | -0.3(-0.36,-0.23) | -0.31(-0.38,-0.24) |
| Cholesterol esters in small VLDL (mmol/l) | 0.39(0.34,0.44) | 0.37(0.32,0.42) | -0.27(-0.33,-0.21) | -0.28(-0.35,-0.21) |
| Free cholesterol in small VLDL (mmol/l) | 0.3(0.24,0.35) | 0.29(0.24,0.34) | -0.32(-0.38,-0.25) | -0.34(-0.41,-0.27) |
| Triglycerides in small VLDL (mmol/l) | 0.21(0.15,0.26) | 0.19(0.14,0.24) | -0.49(-0.56,-0.43) | -0.53(-0.6,-0.46) |
| Concentration of very small VLDL particles (mol/l) | 0.39(0.34,0.44) | 0.39(0.34,0.44) | 0.08(0.01,0.14) | 0.03(-0.04,0.1) |
| Total lipids in very small VLDL (mmol/l) | 0.39(0.34,0.44) | 0.39(0.34,0.44) | 0.03(-0.03,0.09) | 0.04(-0.03,0.11) |
| Phospholipids in very small VLDL (mmol/l) | 0.36(0.31,0.41) | 0.37(0.32,0.42) | 0.2(0.14,0.26) | 0.15(0.08,0.22) |
| Total cholesterol in very small VLDL (mmol/l) | 0.32(0.27,0.37) | 0.31(0.25,0.36) | 0.02(-0.05,0.08) | 0.04(-0.03,0.12) |
| Cholesterol esters in very small VLDL (mmol/l) | 0.35(0.3,0.4) | 0.32(0.27,0.37) | 0.02(-0.04,0.09) | 0.06(-0.01,0.13) |
| Free cholesterol in very small VLDL (mmol/l) | 0.23(0.18,0.28) | 0.24(0.19,0.29) | 0(-0.06,0.07) | 0.01(-0.06,0.08) |
| Triglycerides in very small VLDL (mmol/l) | 0.28(0.22,0.33) | 0.29(0.24,0.34) | -0.11(-0.18,-0.05) | -0.14(-0.22,-0.07) |
| Concentration of IDL particles (mol/l) | 0.3(0.25,0.36) | 0.32(0.27,0.37) | 0.23(0.17,0.29) | 0.21(0.14,0.28) |
| Total lipids in IDL (mmol/l) | 0.32(0.27,0.37) | 0.33(0.28,0.39) | 0.22(0.16,0.28) | 0.2(0.13,0.27) |
| Phospholipids in IDL (mmol/l) | 0.27(0.22,0.32) | 0.29(0.24,0.34) | 0.26(0.2,0.32) | 0.24(0.17,0.31) |
| Total cholesterol in IDL (mmol/l) | 0.32(0.27,0.37) | 0.32(0.27,0.38) | 0.17(0.11,0.23) | 0.15(0.08,0.23) |
| Cholesterol esters in IDL (mmol/l) | 0.33(0.28,0.38) | 0.33(0.28,0.38) | 0.12(0.06,0.18) | 0.11(0.04,0.18) |
| Free cholesterol in IDL (mmol/l) | 0.26(0.21,0.31) | 0.28(0.23,0.33) | 0.28(0.22,0.34) | 0.26(0.19,0.33) |
| Triglycerides in IDL (mmol/l) | 0.25(0.2,0.3) | 0.29(0.24,0.35) | 0.43(0.37,0.49) | 0.4(0.34,0.47) |
| Concentration of large LDL particles (mol/l) | 0.26(0.21,0.31) | 0.28(0.23,0.33) | 0.17(0.1,0.23) | 0.13(0.06,0.2) |
| Total lipids in large LDL (mmol/l) | 0.28(0.23,0.34) | 0.31(0.25,0.36) | 0.15(0.09,0.21) | 0.12(0.04,0.19) |
| Phospholipids in large LDL (mmol/l) | 0.29(0.24,0.34) | 0.31(0.26,0.36) | 0.16(0.1,0.23) | 0.13(0.06,0.2) |
| Total cholesterol in large LDL (mmol/l) | 0.28(0.23,0.34) | 0.3(0.25,0.35) | 0.1(0.04,0.16) | 0.06(-0.01,0.13) |
| Cholesterol esters in large LDL (mmol/l) | 0.29(0.24,0.34) | 0.31(0.26,0.36) | 0.06(0,0.12) | 0.02(-0.05,0.09) |
| Free cholesterol in large LDL (mmol/l) | 0.26(0.21,0.31) | 0.28(0.23,0.33) | 0.21(0.15,0.27) | 0.17(0.1,0.24) |
| Triglycerides in large LDL (mmol/l) | 0.18(0.12,0.23) | 0.23(0.18,0.28) | 0.63(0.57,0.69) | 0.6(0.54,0.67) |
| Concentration of medium LDL particles (mol/l) | 0.24(0.19,0.3) | 0.27(0.22,0.32) | 0.07(0.01,0.13) | 0.03(-0.04,0.1) |
| Total lipids in medium LDL (mmol/l) | 0.27(0.22,0.33) | 0.3(0.24,0.35) | 0.06(-0.01,0.12) | 0.01(-0.06,0.08) |
| Phospholipids in medium LDL (mmol/l) | 0.3(0.25,0.36) | 0.32(0.27,0.37) | 0.12(0.06,0.18) | 0.04(-0.03,0.11) |
| Total cholesterol in medium LDL (mmol/l) | 0.27(0.22,0.32) | 0.29(0.24,0.34) | 0(-0.07,0.06) | -0.05(-0.12,0.02) |
| Cholesterol esters in medium LDL (mmol/l) | 0.27(0.22,0.32) | 0.29(0.23,0.34) | -0.02(-0.08,0.05) | -0.06(-0.14,0.01) |

**eTable 6: Mean absolute difference in traits in SD units between 7y and 25y in males and females comparing i)findings from multilevel models implemented here to ii)linear regression of traits at 7y and 25y**

| | 7y | | 25y | |
|---|---|---|---|---|
| | Mean absolute sex difference in SD units (95% CI) | | Mean absolute sex difference in SD units (95% CI) | |
| | MLM | Regression | MLM | Regression |
| Free cholesterol in medium LDL (mmol/l) | 0.27(0.22,0.32) | 0.29(0.24,0.34) | 0.04(-0.02,0.1) | 0.01(-0.06,0.08) |
| Triglycerides in medium LDL (mmol/l) | 0.11(0.06,0.17) | 0.17(0.12,0.22) | 0.7(0.64,0.75) | 0.65(0.59,0.72) |
| Concentration of small LDL particles (mol/l) | 0.22(0.17,0.27) | 0.24(0.19,0.29) | 0.06(0,0.12) | 0.02(-0.05,0.09) |
| Total lipids in small LDL (mmol/l) | 0.26(0.2,0.31) | 0.28(0.22,0.33) | 0.05(-0.01,0.11) | 0.01(-0.06,0.08) |
| Phospholipids in small LDL (mmol/l) | 0.25(0.19,0.3) | 0.26(0.21,0.32) | 0.15(0.09,0.21) | 0.08(0.01,0.15) |
| Total cholesterol in small LDL (mmol/l) | 0.26(0.2,0.31) | 0.27(0.22,0.33) | 0(-0.06,0.06) | -0.04(-0.11,0.03) |
| Cholesterol esters in small LDL (mmol/l) | 0.25(0.2,0.3) | 0.27(0.22,0.32) | 0(-0.06,0.06) | -0.05(-0.12,0.02) |
| Free cholesterol in small LDL (mmol/l) | 0.23(0.18,0.28) | 0.25(0.19,0.3) | 0.01(-0.05,0.07) | 0(-0.07,0.07) |
| Triglycerides in small LDL (mmol/l) | 0.17(0.12,0.22) | 0.19(0.14,0.24) | 0.42(0.35,0.48) | 0.33(0.26,0.4) |
| Concentration of very large HDL particles (mol/l) | -0.1(-0.15,-0.05) | -0.05(-0.1,0) | 0.82(0.77,0.87) | 0.85(0.79,0.91) |
| Total lipids in very large HDL (mmol/l) | -0.11(-0.17,-0.06) | -0.07(-0.12,-0.02) | 0.8(0.75,0.85) | 0.85(0.79,0.91) |
| Phospholipids in very large HDL (mmol/l) | -0.11(-0.16,-0.06) | -0.06(-0.11,0) | 0.87(0.81,0.92) | 0.88(0.81,0.94) |
| Total cholesterol in very large HDL (mmol/l) | -0.13(-0.18,-0.07) | -0.09(-0.14,-0.04) | 0.72(0.67,0.78) | 0.79(0.73,0.85) |
| Cholesterol esters in very large HDL (mmol/l) | -0.13(-0.18,-0.08) | -0.1(-0.15,-0.05) | 0.68(0.62,0.74) | 0.76(0.7,0.82) |
| Free cholesterol in very large HDL (mmol/l) | -0.11(-0.16,-0.05) | -0.07(-0.12,-0.01) | 0.8(0.75,0.85) | 0.86(0.8,0.92) |
| Triglycerides in very large HDL (mmol/l) | 0.11(0.05,0.16) | 0.13(0.08,0.18) | 0.42(0.36,0.48) | 0.44(0.37,0.51) |
| Concentration of large HDL particles (mol/l) | -0.19(-0.24,-0.14) | -0.12(-0.17,-0.07) | 0.86(0.8,0.91) | 0.89(0.82,0.95) |
| Total lipids in large HDL (mmol/l) | -0.18(-0.24,-0.13) | -0.12(-0.17,-0.06) | 0.85(0.8,0.91) | 0.88(0.82,0.94) |
| Phospholipids in large HDL (mmol/l) | -0.17(-0.23,-0.12) | -0.1(-0.16,-0.05) | 0.86(0.81,0.92) | 0.89(0.83,0.95) |
| Total cholesterol in large HDL (mmol/l) | -0.2(-0.25,-0.14) | -0.13(-0.19,-0.08) | 0.83(0.78,0.89) | 0.87(0.81,0.93) |
| Cholesterol esters in large HDL (mmol/l) | -0.2(-0.25,-0.15) | -0.14(-0.19,-0.09) | 0.83(0.78,0.89) | 0.87(0.8,0.93) |
| Free cholesterol in large HDL (mmol/l) | -0.17(-0.22,-0.12) | -0.11(-0.17,-0.06) | 0.83(0.78,0.89) | 0.87(0.81,0.93) |
| Triglycerides in large HDL (mmol/l) | 0.07(0.02,0.13) | 0.12(0.07,0.17) | 0.76(0.71,0.81) | 0.8(0.74,0.86) |
| Concentration of medium HDL particles (mol/l) | -0.18(-0.23,-0.12) | -0.1(-0.15,-0.05) | 0.62(0.56,0.68) | 0.61(0.55,0.68) |
| Total lipids in medium HDL (mmol/l) | -0.17(-0.23,-0.12) | -0.11(-0.16,-0.06) | 0.64(0.59,0.7) | 0.61(0.54,0.67) |
| Phospholipids in medium HDL (mmol/l) | -0.17(-0.23,-0.12) | -0.11(-0.16,-0.05) | 0.7(0.64,0.75) | 0.66(0.6,0.73) |
| Total cholesterol in medium HDL (mmol/l) | -0.2(-0.26,-0.15) | -0.14(-0.19,-0.09) | 0.54(0.48,0.6) | 0.54(0.48,0.61) |
| Cholesterol esters in medium HDL (mmol/l) | -0.22(-0.27,-0.16) | -0.16(-0.21,-0.11) | 0.52(0.46,0.58) | 0.53(0.46,0.59) |
| Free cholesterol in medium HDL (mmol/l) | -0.12(-0.18,-0.07) | -0.06(-0.11,0) | 0.6(0.54,0.66) | 0.61(0.54,0.67) |
| Triglycerides in medium HDL (mmol/l) | 0.16(0.11,0.21) | 0.17(0.12,0.23) | 0.24(0.18,0.31) | 0.19(0.12,0.26) |
| Concentration of small HDL particles (mol/l) | -0.11(-0.17,-0.06) | -0.08(-0.13,-0.03) | 0.26(0.2,0.32) | 0.16(0.1,0.23) |

**eTable 6: Mean absolute difference in traits in SD units between 7y and 25y in males and females comparing i)findings from multilevel models implemented here to ii)linear regression of traits at 7y and 25y**

| | 7y | | 25y | |
|---|---|---|---|---|
| | Mean absolute sex difference in SD units (95% CI) | | Mean absolute sex difference in SD units (95% CI) | |
| | MLM | Regression | MLM | Regression |
| Total lipids in small HDL (mmol/l) | -0.14(-0.19,-0.08) | -0.08(-0.13,-0.02) | 0.25(0.19,0.31) | 0.17(0.1,0.23) |
| Phospholipids in small HDL (mmol/l) | -0.17(-0.22,-0.11) | -0.15(-0.2,-0.09) | 0.27(0.21,0.33) | 0.27(0.2,0.33) |
| Total cholesterol in small HDL (mmol/l) | -0.07(-0.13,-0.02) | -0.01(-0.06,0.05) | 0.24(0.18,0.3) | 0.09(0.02,0.16) |
| Cholesterol esters in small HDL (mmol/l) | -0.02(-0.08,0.04) | 0.04(-0.01,0.09) | 0.23(0.17,0.29) | 0.06(-0.01,0.13) |
| Free cholesterol in small HDL (mmol/l) | -0.27(-0.32,-0.21) | -0.2(-0.26,-0.15) | 0.22(0.16,0.28) | 0.22(0.16,0.29) |
| Triglycerides in small HDL (mmol/l) | 0.16(0.1,0.21) | 0.16(0.11,0.21) | -0.14(-0.2,-0.07) | -0.21(-0.28,-0.14) |
| Mean diameter for VLDL particles (mm) | 0.1(0.04,0.15) | 0.07(0.02,0.13) | -0.64(-0.71,-0.58) | -0.65(-0.72,-0.58) |
| Mean diameter for LDL particles (mm) | 0.09(0.04,0.15) | 0.06(0.01,0.12) | 0.25(0.19,0.31) | 0.36(0.29,0.42) |
| Mean diameter for HDL particles (mm) | -0.12(-0.17,-0.06) | -0.07(-0.12,-0.01) | 0.84(0.78,0.89) | 0.86(0.8,0.92) |
| Serum total cholesterol (mmol/l) | 0.24(0.18,0.29) | 0.27(0.21,0.32) | 0.32(0.26,0.38) | 0.31(0.24,0.37) |
| Total cholesterol in VLDL (mmol/l) | 0.33(0.28,0.38) | 0.31(0.26,0.36) | -0.29(-0.35,-0.22) | -0.29(-0.37,-0.22) |
| Remnant cholesterol (non-HDL, non-LDL -cholesterol) (mmol/l) | 0.37(0.32,0.42) | 0.36(0.31,0.41) | -0.09(-0.15,-0.03) | -0.1(-0.17,-0.03) |
| Total cholesterol in LDL (mmol/l) | 0.28(0.22,0.33) | 0.29(0.24,0.35) | 0.05(-0.01,0.11) | 0(-0.07,0.08) |
| Total cholesterol in HDL (mmol/l) | -0.19(-0.24,-0.14) | -0.12(-0.17,-0.07) | 0.78(0.73,0.84) | 0.81(0.75,0.87) |
| Total cholesterol in HDL2 (mmol/l) | -0.21(-0.27,-0.16) | -0.15(-0.2,-0.1) | 0.8(0.75,0.85) | 0.83(0.76,0.89) |
| Total cholesterol in HDL3 (mmol/l) | -0.12(-0.18,-0.07) | -0.06(-0.11,0) | 0.61(0.55,0.68) | 0.51(0.44,0.57) |
| Esterified cholesterol (mmol/l) | 0.23(0.18,0.28) | 0.25(0.2,0.31) | 0.33(0.27,0.39) | 0.28(0.21,0.35) |
| Free cholesterol (mmol/l) | 0.25(0.19,0.3) | 0.28(0.22,0.33) | 0.37(0.31,0.43) | 0.35(0.29,0.42) |
| Serum total triglycerides (mmol/l) | 0.19(0.14,0.24) | 0.18(0.13,0.23) | -0.29(-0.36,-0.22) | -0.32(-0.4,-0.25) |
| Triglycerides in VLDL (mmol/l) | 0.17(0.11,0.22) | 0.15(0.1,0.2) | -0.47(-0.54,-0.41) | -0.49(-0.56,-0.42) |
| Triglycerides in LDL (mmol/l) | 0.16(0.1,0.21) | 0.2(0.15,0.26) | 0.61(0.55,0.67) | 0.57(0.51,0.64) |
| Triglycerides in HDL (mmol/l) | 0.17(0.11,0.22) | 0.19(0.13,0.24) | 0.45(0.38,0.51) | 0.42(0.36,0.49) |
| Diacylglycerol (mmol/l)* | 0.09(0.04,0.15) | 0.08(0.03,0.13) | 0.1(0.03,0.18) | 0.11(0.04,0.18) |
| Total phosphoglycerides (mmol/l) | 0.08(0.03,0.14) | 0.12(0.07,0.18) | 0.65(0.6,0.71) | 0.63(0.57,0.7) |
| Phosphatidylcholine and other cholines (mmol/l) | 0.05(-0.01,0.1) | 0.1(0.05,0.16) | 0.47(0.41,0.53) | 0.46(0.4,0.53) |
| Total cholines (mmol/l) | 0.1(0.05,0.15) | 0.14(0.09,0.19) | 0.61(0.56,0.67) | 0.57(0.51,0.63) |
| Apolipoprotein A-I (g/l) | -0.07(-0.13,-0.02) | -0.01(-0.06,0.04) | 0.68(0.62,0.73) | 0.69(0.62,0.75) |
| Apolipoprotein B (g/l) | 0.36(0.31,0.41) | 0.35(0.3,0.4) | -0.15(-0.21,-0.08) | -0.19(-0.26,-0.12) |
| Total fatty acids (mmol/l) | 0.23(0.18,0.29) | 0.25(0.2,0.31) | 0.24(0.18,0.3) | 0.2(0.13,0.27) |
| Fatty acid length* | 0.01(-0.04,0.07) | -0.01(-0.06,0.04) | 0.02(-0.05,0.1) | 0.01(-0.06,0.08) |
| Estimated degree of unsaturation* | -0.07(-0.12,-0.01) | -0.05(-0.1,0.01) | 0.12(0.04,0.19) | 0.12(0.05,0.19) |

**eTable 6: Mean absolute difference in traits in SD units between 7y and 25y in males and females comparing i)findings from multilevel models implemented here to ii)linear regression of traits at 7y and 25y**

| | 7y | | 25y | |
|---|---|---|---|---|
| | Mean absolute sex difference in SD units (95% CI) | | Mean absolute sex difference in SD units (95% CI) | |
| | MLM | Regression | MLM | Regression |
| 22:6, docosahexaenoic acid (mmol/l) | 0.19(0.14,0.24) | 0.23(0.18,0.28) | 0.59(0.53,0.65) | 0.54(0.48,0.61) |
| 18:2, linoleic acid (mmol/l) | 0.23(0.18,0.28) | 0.26(0.21,0.31) | 0.23(0.17,0.29) | 0.17(0.1,0.24) |
| Conjugated linoleic acid (mmol/l)* | 0.12(0.06,0.17) | 0.12(0.07,0.17) | 0.1(0.03,0.18) | 0.1(0.03,0.17) |
| Omega-3 fatty acids (mmol/l) | 0.17(0.11,0.22) | 0.19(0.14,0.24) | 0.09(0.03,0.16) | 0.06(-0.01,0.13) |
| Omega-6 fatty acids (mmol/l) | 0.22(0.17,0.27) | 0.25(0.2,0.31) | 0.32(0.26,0.38) | 0.28(0.21,0.35) |
| Polyunsaturated fatty acids (mmol/l) | 0.22(0.17,0.28) | 0.26(0.2,0.31) | 0.3(0.24,0.37) | 0.26(0.19,0.33) |
| Monounsaturated fatty acids; 16:1, 18:1 (mmol/l) | 0.24(0.19,0.29) | 0.25(0.2,0.31) | 0.14(0.07,0.2) | 0.13(0.05,0.2) |
| Saturated fatty acids (mmol/l) | 0.17(0.11,0.22) | 0.19(0.14,0.24) | 0.22(0.16,0.29) | 0.19(0.12,0.26) |
| Glucose (mmol/l) | -0.15(-0.21,-0.09) | -0.15(-0.21,-0.1) | -0.42(-0.49,-0.35) | -0.41(-0.48,-0.33) |
| Lactate (mmol/l) | 0.12(0.07,0.18) | 0.12(0.07,0.17) | -0.02(-0.08,0.05) | 0.01(-0.05,0.08) |
| Citrate (mmol/l) | 0.12(0.07,0.18) | 0.12(0.07,0.18) | -0.08(-0.14,-0.01) | -0.1(-0.17,-0.03) |
| Alanine (mmol/l) | 0.07(0.01,0.12) | 0.03(-0.02,0.09) | -0.16(-0.23,-0.1) | -0.19(-0.26,-0.12) |
| Glutamine (mmol/l) | 0.44(0.39,0.49) | 0.35(0.3,0.4) | -0.83(-0.89,-0.77) | -0.8(-0.87,-0.74) |
| Histidine (mmol/l) | 0.17(0.12,0.21) | 0.14(0.09,0.18) | -0.4(-0.47,-0.33) | -0.33(-0.4,-0.26) |
| Isoleucine (mmol/l) | 0.1(0.05,0.16) | 0.06(0.01,0.11) | -0.83(-0.9,-0.77) | -0.83(-0.9,-0.77) |
| Leucine (mmol/l) | -0.01(-0.07,0.04) | -0.06(-0.11,0) | -0.92(-0.98,-0.86) | -0.93(-0.99,-0.86) |
| Valine (mmol/l) | 0.06(0,0.12) | 0.02(-0.03,0.08) | -0.85(-0.91,-0.79) | -0.83(-0.89,-0.76) |
| Phenylalanine (mmol/l) | -0.03(-0.09,0.02) | -0.05(-0.1,0.01) | -0.32(-0.38,-0.25) | -0.35(-0.41,-0.28) |
| Tyrosine (mmol/l) | 0.01(-0.05,0.07) | -0.01(-0.06,0.05) | -0.55(-0.62,-0.49) | -0.52(-0.58,-0.45) |
| Acetate (mmol/l) | 0.04(-0.02,0.1) | 0.03(-0.02,0.08) | -0.16(-0.22,-0.1) | -0.17(-0.24,-0.1) |
| Acetoacetate (mmol/l) | 0.04(-0.01,0.09) | 0.08(0.03,0.13) | -0.2(-0.31,-0.08) | -0.11(-0.18,-0.04) |
| 3-hydroxybutyrate (mmol/l) | 0.12(0.07,0.18) | 0.14(0.09,0.2) | 0.18(0.12,0.25) | 0.16(0.1,0.22) |
| Creatinine (mmol/l) | 0.1(0.05,0.15) | -0.02(-0.07,0.03) | -10.25(-10.3,-10.2) | -10.24(-10.3,-10.18) |
| Albumin (mmol/l) | 0.3(0.24,0.35) | 0.23(0.18,0.28) | -0.52(-0.58,-0.45) | -0.53(-0.6,-0.46) |
| Glycoprotein acetyls, mainly a1-acid glycoprotein (mmol/l) | 0.23(0.18,0.29) | 0.24(0.19,0.29) | 0.1(0.04,0.17) | 0.08(0.01,0.15) |

*These metabolites (diacylglycerol, fatty acid chain length, estimated degree of saturation and conjugated linoleic acid) were not measured at 25y; all models include data only up to aged 18y and values in this table for these traits are at 7y and 18y respectively. HDL: high-density lipoprotein; IDL: intermediate-density lipoprotein; LDL: low-density lipoprotein; MLM; multilevel model; VLDL: very-low-density lipoprotein.

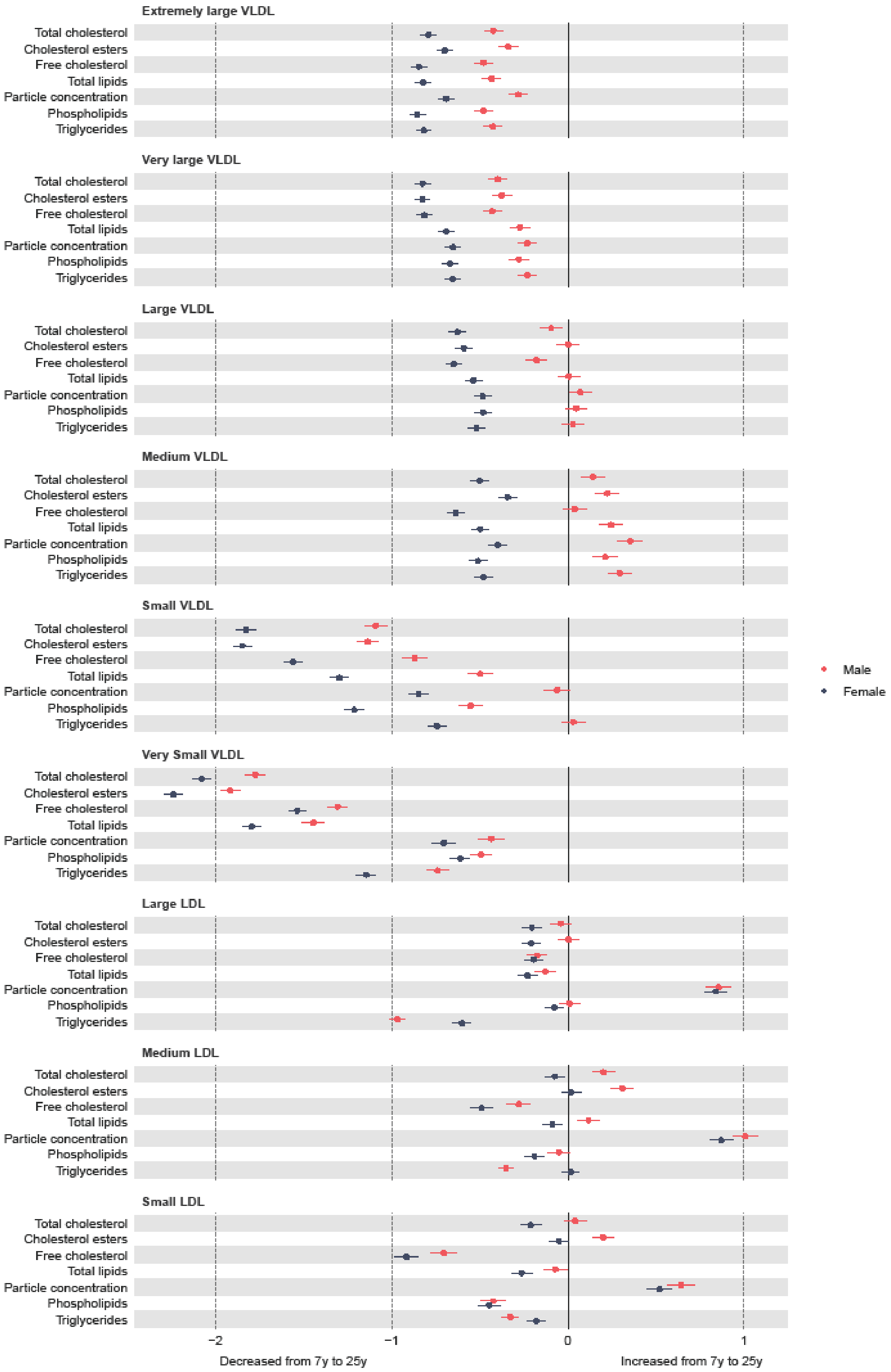


**Figure 1 Mean sex-specific change in VLDL and LDL lipid concentrations in SD units from 7y to 25y, estimated from multilevel models. Legend:** LDL, low-density lipoprotein; VLDL, very-low-density lipoprotein.

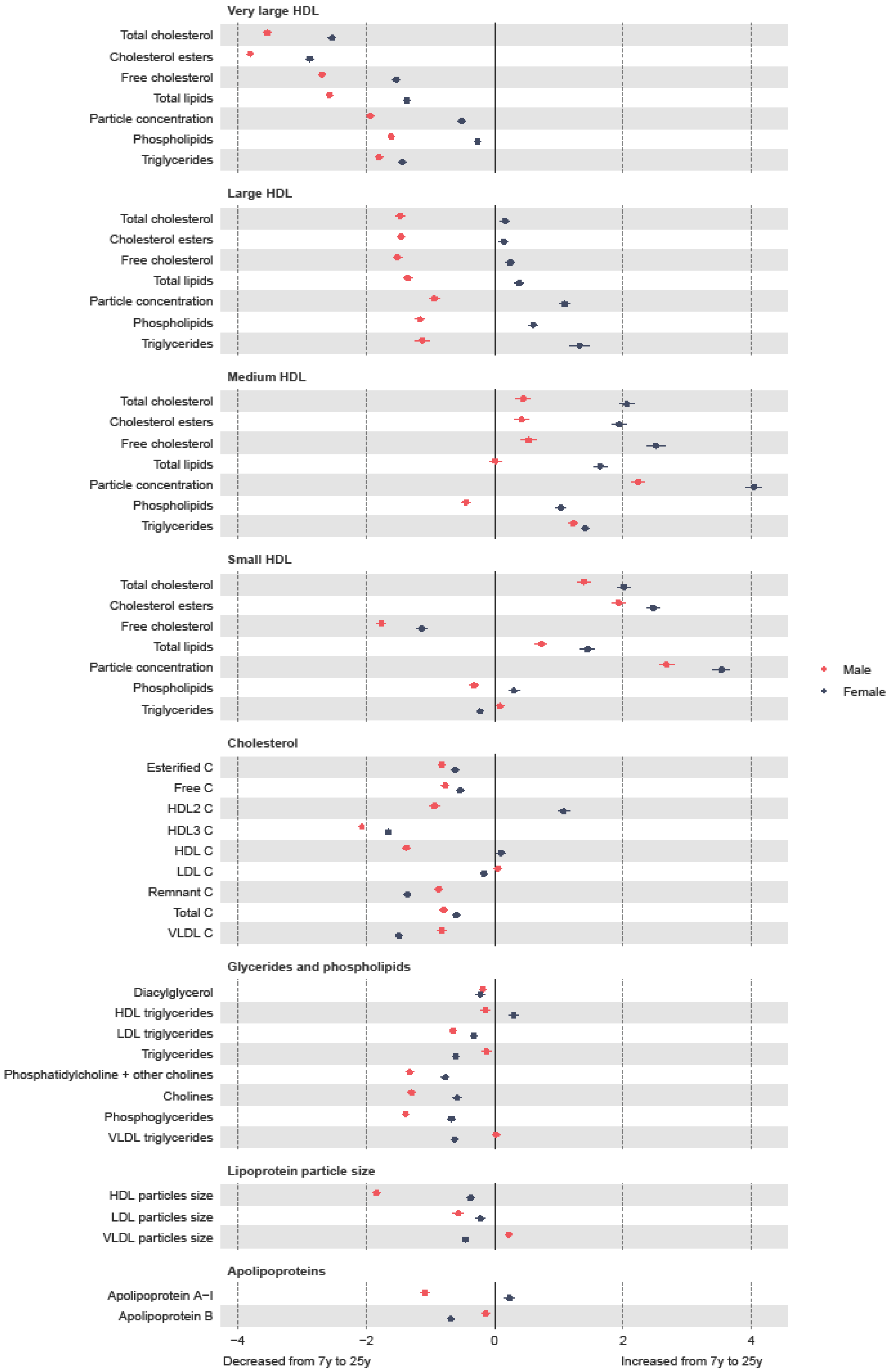


**Figure 2 Mean sex-specific change in lipid concentrations in SD units from 7y to 25y, estimated from multilevel models. Legend:** HDL, high-density lipoprotein; LDL, low-density lipoprotein; VLDL, very-low-density lipoprotein. Note that diacylglycerol is only measured up to 18y.

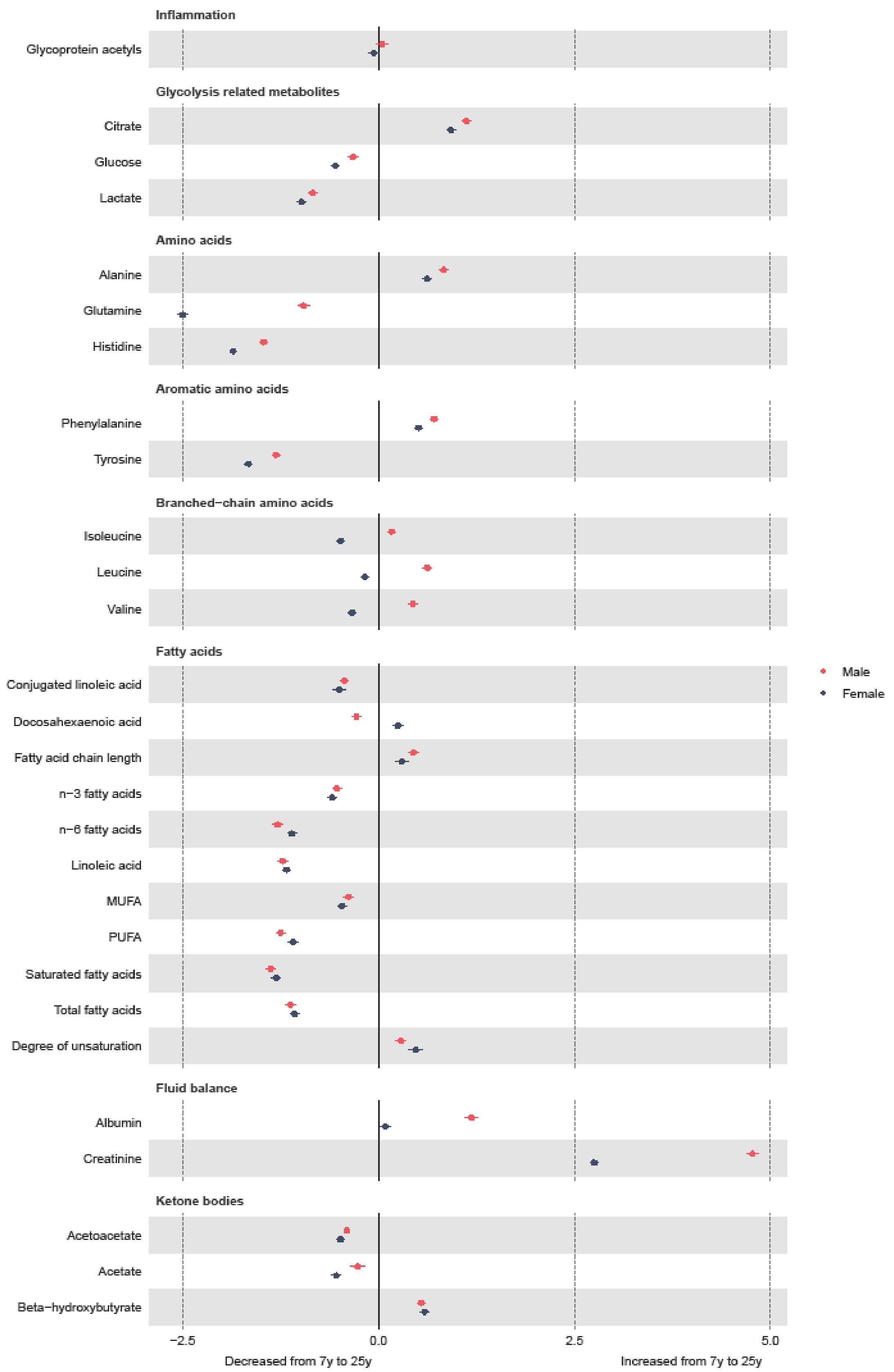


**Figure 3 Mean sex-specific change other trait concentrations in SD units from 7y to 25y, estimated from multilevel models. Legend:** MUFA, monounsaturated fatty acids; PUFA, polyunsaturated fatty acids. Note that conjugated linoleic acid, fatty acid chain length and estimated degree of unsaturation are only measured up to 18y.

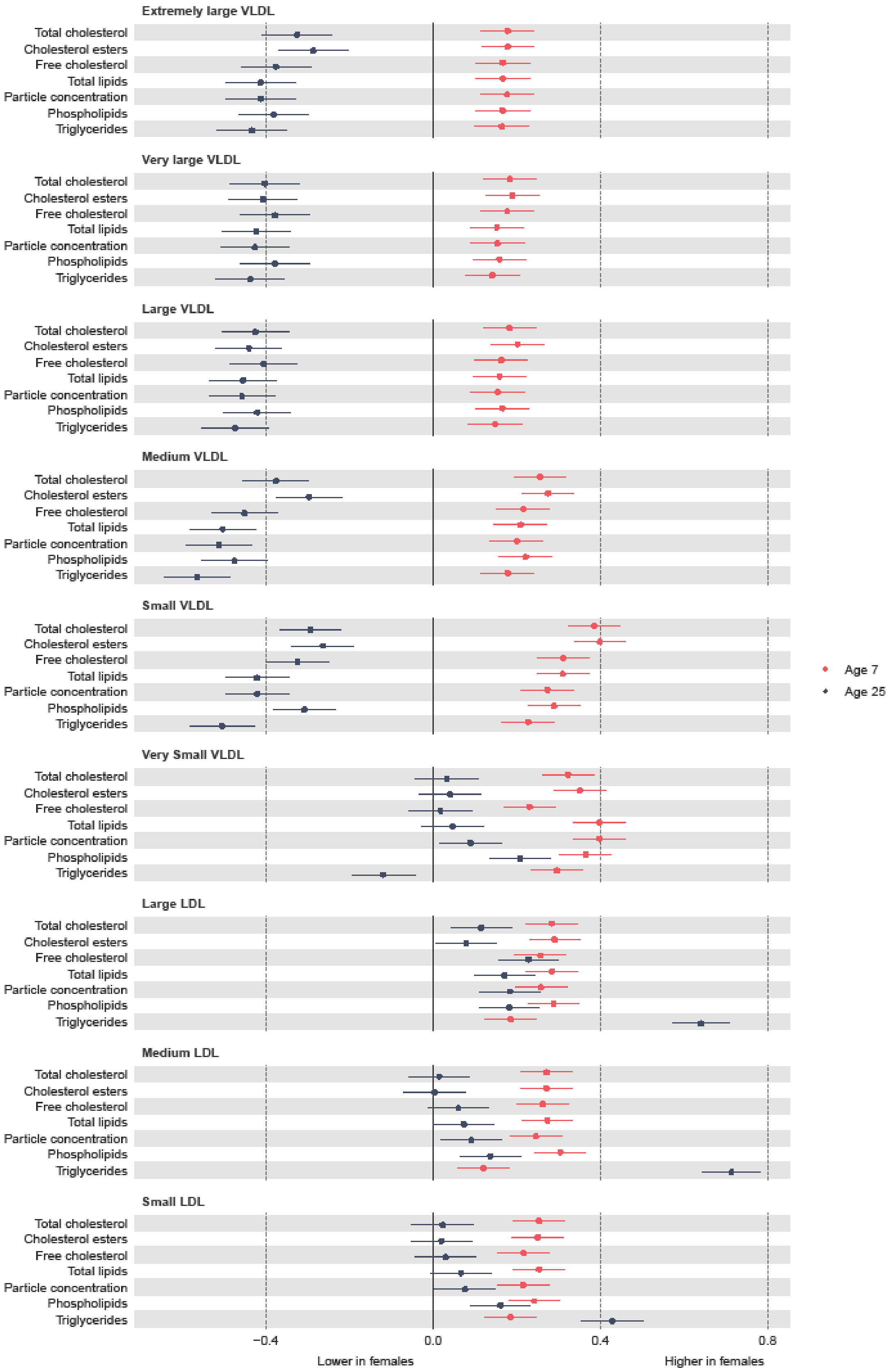


**Figure 4 Mean sex difference in VLDL and LDL lipoprotein concentrations in SD units at 7y and 25y, estimated from multilevel models weighted by the probability of inclusion in analysis. Legend:** LDL, low-density lipoprotein; VLDL, very-low-density lipoprotein.

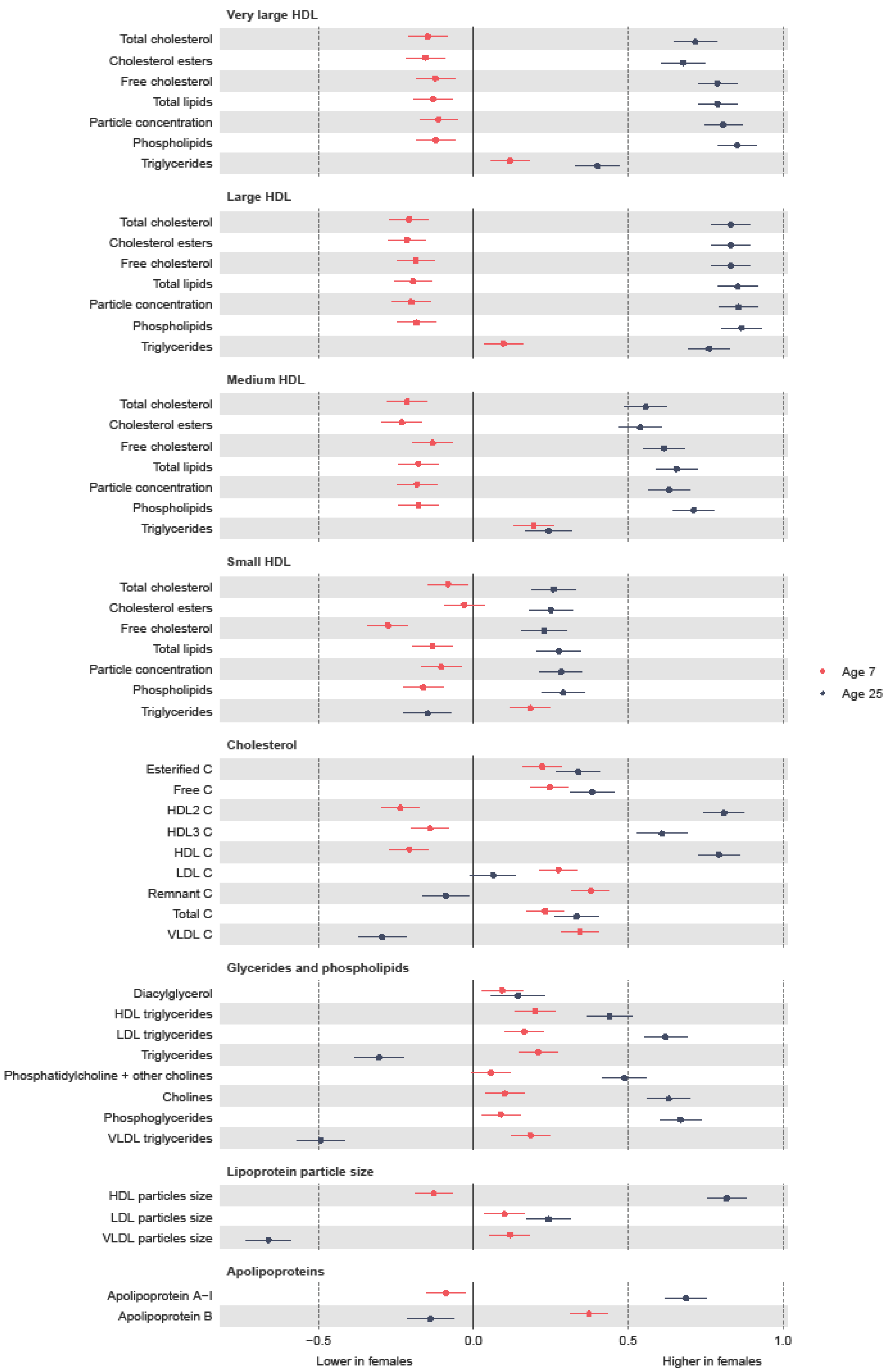


**Figure 5 Mean sex difference in lipid concentrations in SD units at 7y and 25y, estimated from multilevel models weighted by the probability of inclusion in analysis. Legend:** HDL, high-density lipoprotein; LDL, low-density lipoprotein; VLDL, very-low-density lipoprotein. Note that diacylglycerol is only measured up to 18y.

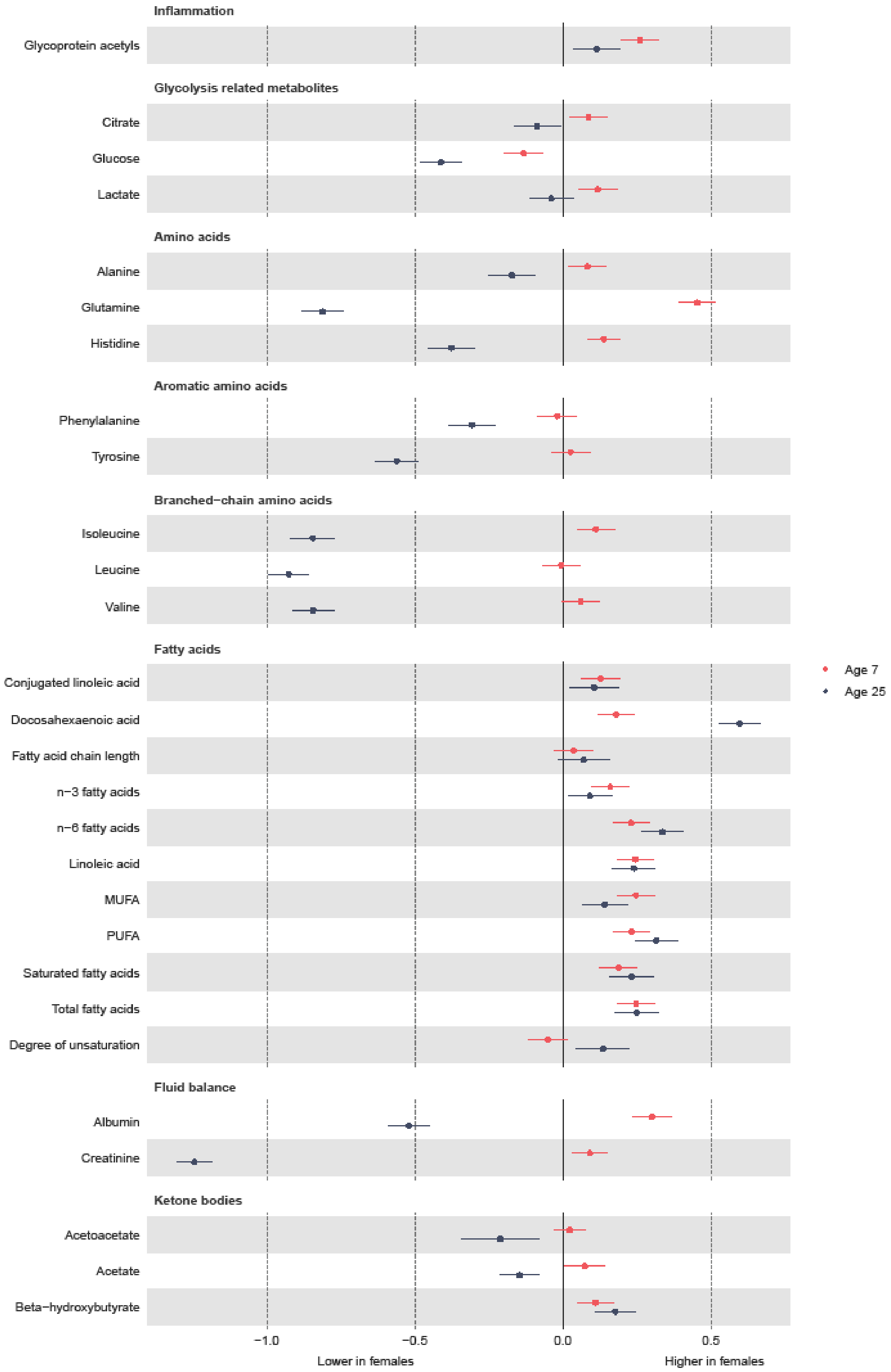


**Figure 6 Mean sex difference in other trait concentrations in SD units at 7y and 25y, estimated from multilevel models weighted by the probability of inclusion in analysis. Legend:** MUFA, monounsaturated fatty acids; PUFA, polyunsaturated fatty acids. Note that conjugated linoleic acid, fatty acid chain length and estimated degree of unsaturation are only measured up to 18y.

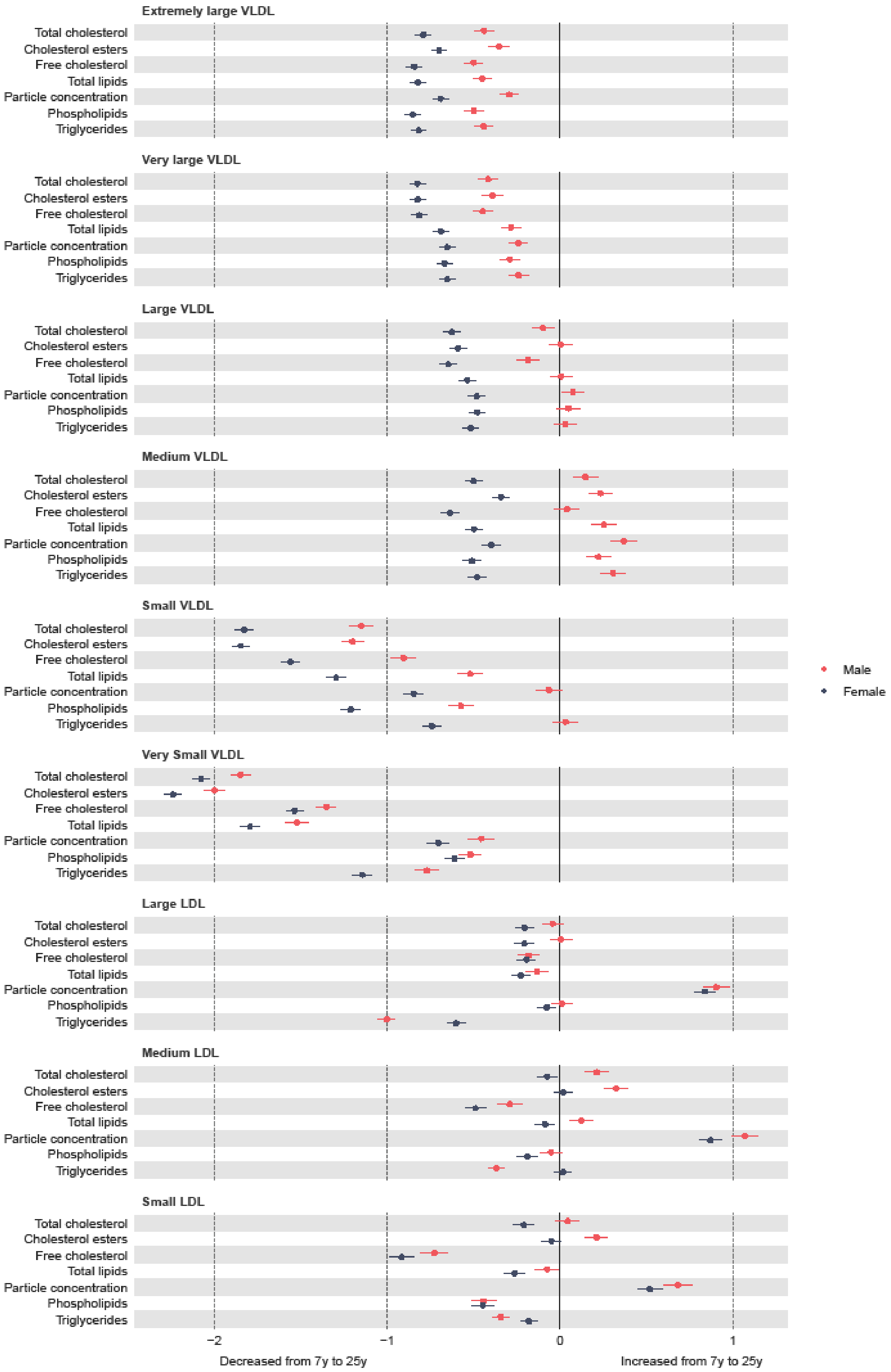


**Figure 7 Mean sex-specific change in VLDL and LDL lipoprotein concentrations in SD units (standardised using sex-specific SDs) from 7y to 25y, estimated from multilevel models. Legend:** LDL, low-density lipoprotein; VLDL, very-low-density lipoprotein.

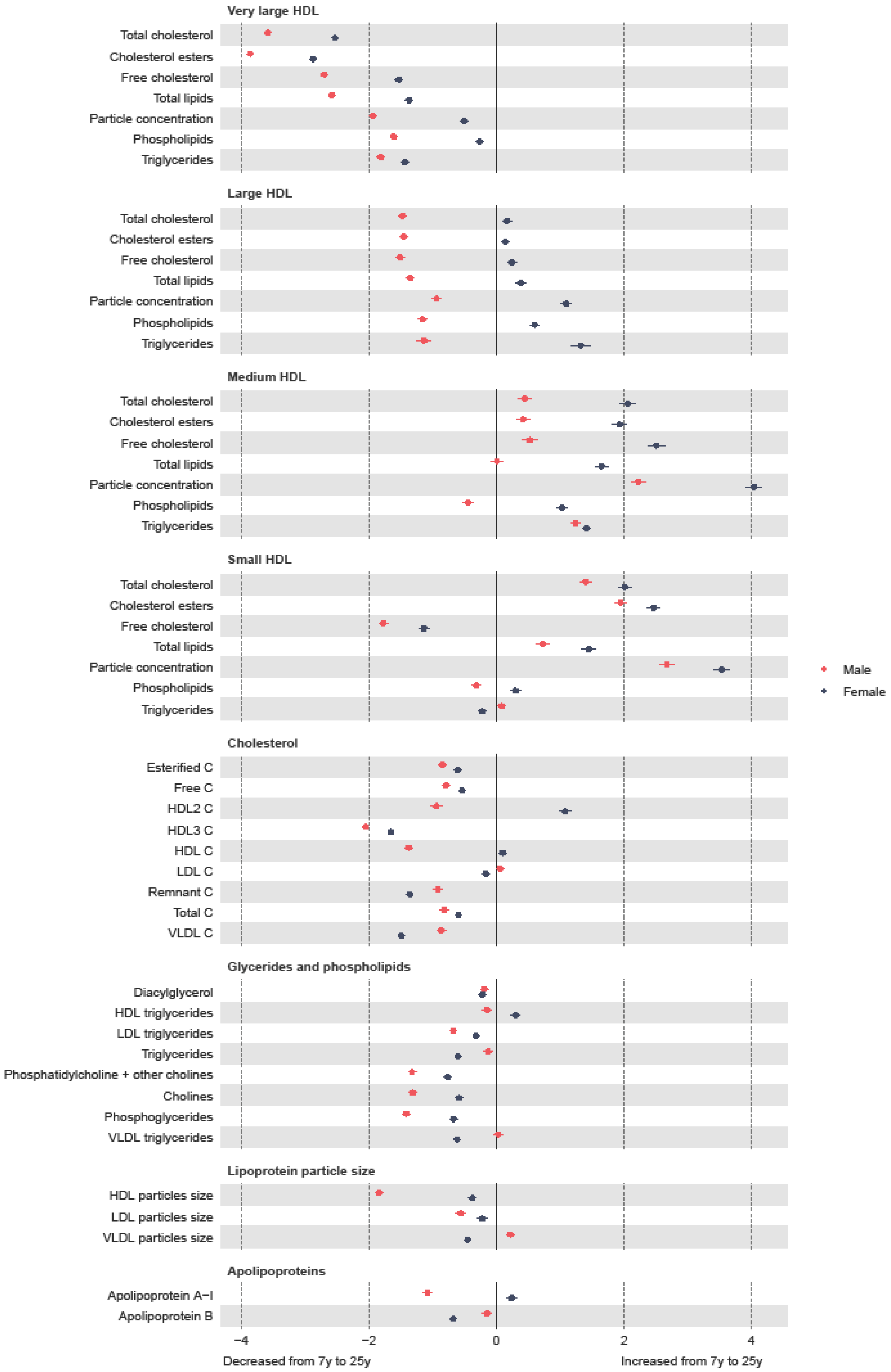


**Figure 8 Mean sex-specific change in lipid concentrations in SD units (standardised using sex-specific SDs) from 7y to 25y, estimated from multilevel models. Legend:** HDL, high-density lipoprotein; LDL, low-density lipoprotein; VLDL, very-low-density lipoprotein. Note that diacylglycerol is only measured up to 18y.

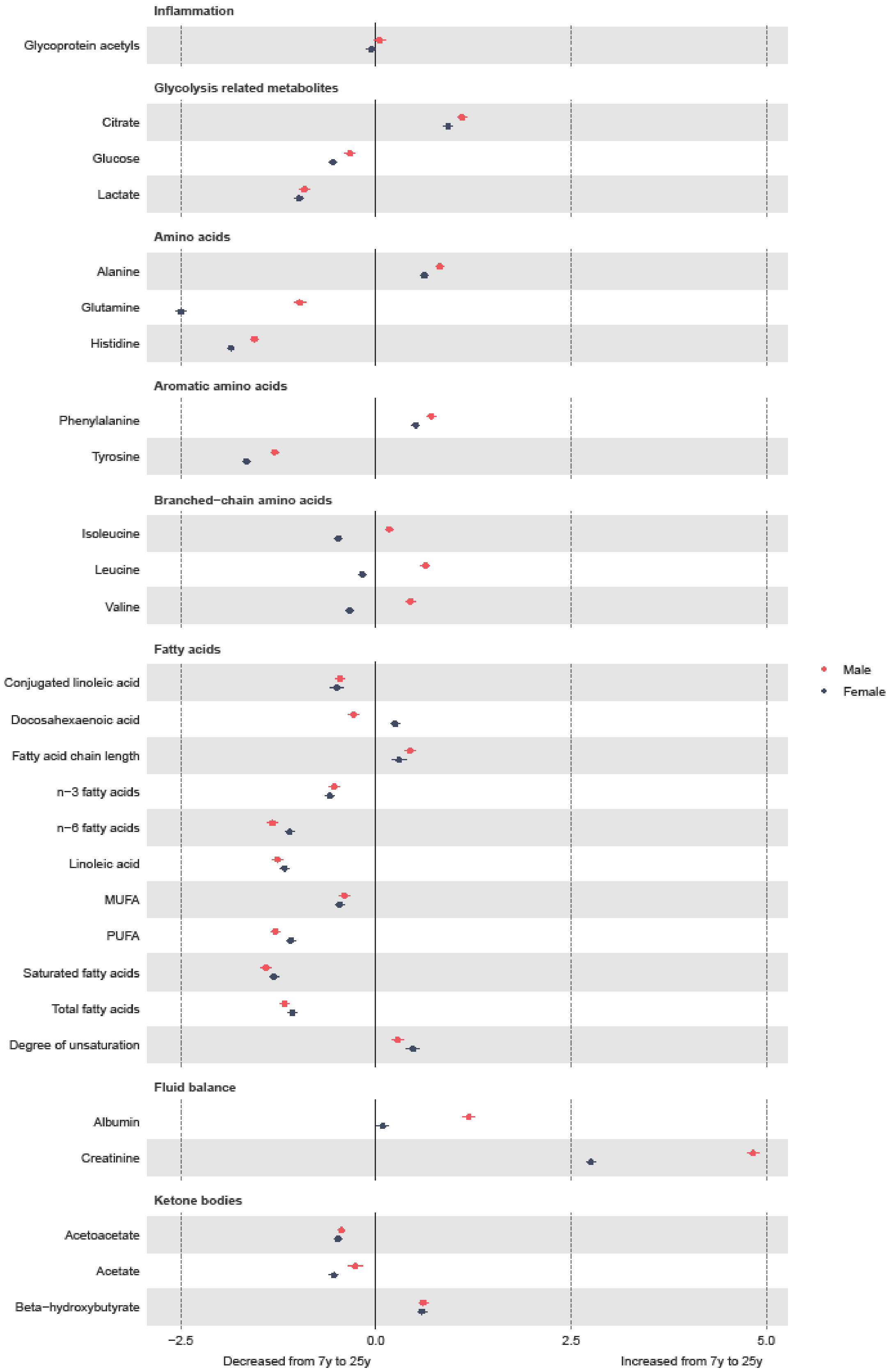


**Figure 9 Mean sex-specific change in other trait concentrations in SD units (standardised using sex-specific SDs) from 7y to 25y, estimated from multilevel models. Legend:** MUFA, monounsaturated fatty acids; PUFA, polyunsaturated fatty acids. Note that conjugated linoleic acid, fatty acid chain length and estimated degree of unsaturation are only measured up to 18y.
